# Unified classification and risk-stratification in Acute Myeloid Leukemia

**DOI:** 10.1101/2022.03.09.22271087

**Authors:** Yanis Tazi, Juan E. Arango-Ossa, Yangyu Zhou, Elsa Bernard, Ian Thomas, Amanda Gilkes, Sylvie Freeman, Yoann Pradat, Sean J Johnson, Robert Hills, Richard Dillon, Max F Levine, Daniel Leongamornlert, Adam Butler, Arnold Ganser, Lars Bullinger, Konstanze Döhner, Oliver Ottmann, Richard Adams, Hartmut Döhner, Peter J Campbell, Alan K Burnett, Michael Dennis, Nigel H Russell, Sean M. Devlin, Brian J P Huntly, Elli Papaemmanuil

## Abstract

Clinical recommendations for AML classification and risk-stratification remain heavily reliant on cytogenetic findings at diagnosis, which are present in <50% of patients. Using comprehensive molecular profiling data from 3,653 patients we characterize and validate 16 molecular classes describing 100% of AML patients. Each class represents diverse biological AML subgroups, and is associated with distinct clinical presentation, likelihood of response to induction chemotherapy, risk of relapse and death over time. Secondary AML-2, emerges as the second largest class (24%), associates with high-risk disease, poor prognosis irrespective of flow MRD negativity, and derives significant benefit from transplantation. Guided by class membership we derive a 3-tier risk-stratification score that re-stratifies 26% of patients as compared to standard of care. This results in a unified framework for disease classification and risk-stratification in AML that relies on information from cytogenetics and 32 genes. Last, we develop an open-access patient-tailored clinical decision support tool.

## Introduction

Acute Myeloid Leukemias (AML) are overlapping hematological neoplasms associated with rapid onset, progressive and frequently chemoresistant disease^1,2^. Intensive chemotherapy and combination regimens have recently shown improvement in patient response^3-4^, however, the risk of relapse-related mortality remains high^5^. At diagnosis, classification and risk-stratification are critical for treatment decisions^2-4^. Decisions on type of consolidation chemotherapy, timing of hematopoietic stem cell transplantation (HSCT) or eligibility for clinical trials^3^, are evaluated on each patients’ *a priori* likelihood of attaining complete remission, the prospective persistence of measurable residual disease^6^ (MRD), and the predicted likelihood of relapse or death^2^.

As prospective sequencing is becoming routine during AML diagnosis, there is a need to understand the clinical relevance of molecular biomarkers in the context of established endpoints (i.e. MRD, CR, relapse). Translation of such findings into clinical practice warrants the development of evidence-based and dynamic clinical decision support tools that consider molecular and clinical biomarkers to inform optimal diagnosis and treatment decisions and improve patient outcomes^7^.

To this end, gene mutations are being gradually incorporated into classification and risk-stratification guidelines for AML patient management^1,2^. However, with the exception of *NPM1, CEBPA* and provisionally *RUNX1*, the WHO^2016^ classification is primarily reliant on cytogenetic findings^1,2^. Here, we incorporate data from 2,113 representative AML patients enrolled in three UK-NCRI trials^8,9^. We study the relationships between genetic alterations, clinical presentation, treatment response and outcome to develop a framework that unifies diagnostic classification to risk stratification that results in significant improvement in predictive accuracy. Results were validated in an independent cohort of 1,540 AML patients^10^.

## Results

### Study Participants

Study participants included 2,113 AML adult patients enrolled in UK-NCRI trials^3,8,9^ (training), which uniquely recruit up to 80% of UK patients fit for either intensive or non-intensive treatment and are therefore representative of the “real-world” patient population rather than studies limited by strict trial entry criteria. The majority (83%, n=1,755) were intensively treated^8,11,12^ (median age = 56). Data from 1,540 AML patients from the AML-SG^8^ (median age = 50) with comparable molecular annotation were used as a validation cohort (S.Table 1-2, S.Figure 1). Informed consent was obtained for all patients. Molecular assessment of UK-NCRI cohort included karyotypes^8,9^, copy number alterations (CNA) and putative oncogenic mutations across the entire gene body of 128 genes implicated in myeloid neoplasia pathogenesis at diagnosis (S.Table 3-5).

### Genomic landscape of AML

Mapping of recurrent cytogenetic abnormalities and gene mutations characterized 8,460 driver events in 98% of the UK-NCRI cohort (S.Table 4-5). Genotype and clinical relationships for 70 recurrent cytogenetic abnormalities and 84 genes were consistent with prior studies (S.Figure 2-4; https://www.aml-risk-model.com/gene-panel). Detailed genotype and clinical relationships to include patterns of co-mutation, clinical and outcome correlates were evaluated for each of 70 recurrent (>1%) cytogenetic abnormalities and 84 genes with established role in AML pathogenesis (S.Figure 4; https://www.aml-risk-model.com/supplementary).

### Molecular Classification in AML

Utilizing the WHO^2016^ guidelines for AML classification, 49.6% (n=1049) of UK-NCRI patients mapped to established WHO^2016^ classes. Each class ranged in size from 0.4% to 31.5% (S.Figure 5). Clustering analysis on the basis of cytogenetic and gene mutation findings identified 14 non-overlapping clusters classifying 92% (n=1943) of patients (Extended Figure 1-2). These validate established WHO^2016^ entities, resolve provisional subgroups^2,13,14^ and characterize novel entities that describe 33.3% of AML patients (S.Figure 6). Each class is associated with distinct demographic and clinical parameters and in unison, explain the heterogeneity observed at diagnosis across age, peripheral blood and blast counts amongst AML patients (S.Table 6).

Classes defined by cytogenetic alterations included entities defined by translocations and patients with complex karyotype (CK, ≥3 unbalanced abnormalities) (n=217, 10.3%) with frequent involvement of *TP53* mutations (n=141, 65%)^14^. Consistent with prior studies, patients with CK were generally older (median diagnostic age=62) and associated with adverse outcomes^10^. With the exception of mutations in *TP53*, which was mutated in 65% of CK cases, there was a paucity of other acquired mutations in this group. Unlike in MDS^15^, the allelic state of *TP53* (mono allelic or multi-hit) provided no further prognostic information in AML (Extended Figure 3). A novel cytogenetic subgroup was defined by the presence of ≥1 trisomies (n=237, 11.2%), frequently involving +8, +11, +13, +21 and +22 but no deletions. This group had infrequent involvement of *TP53* (4%), and was associated with more favorable disease, even when ≥3 trisomies were present (Figure 1A-B, Extended Figure 4).

**Figure 1:**
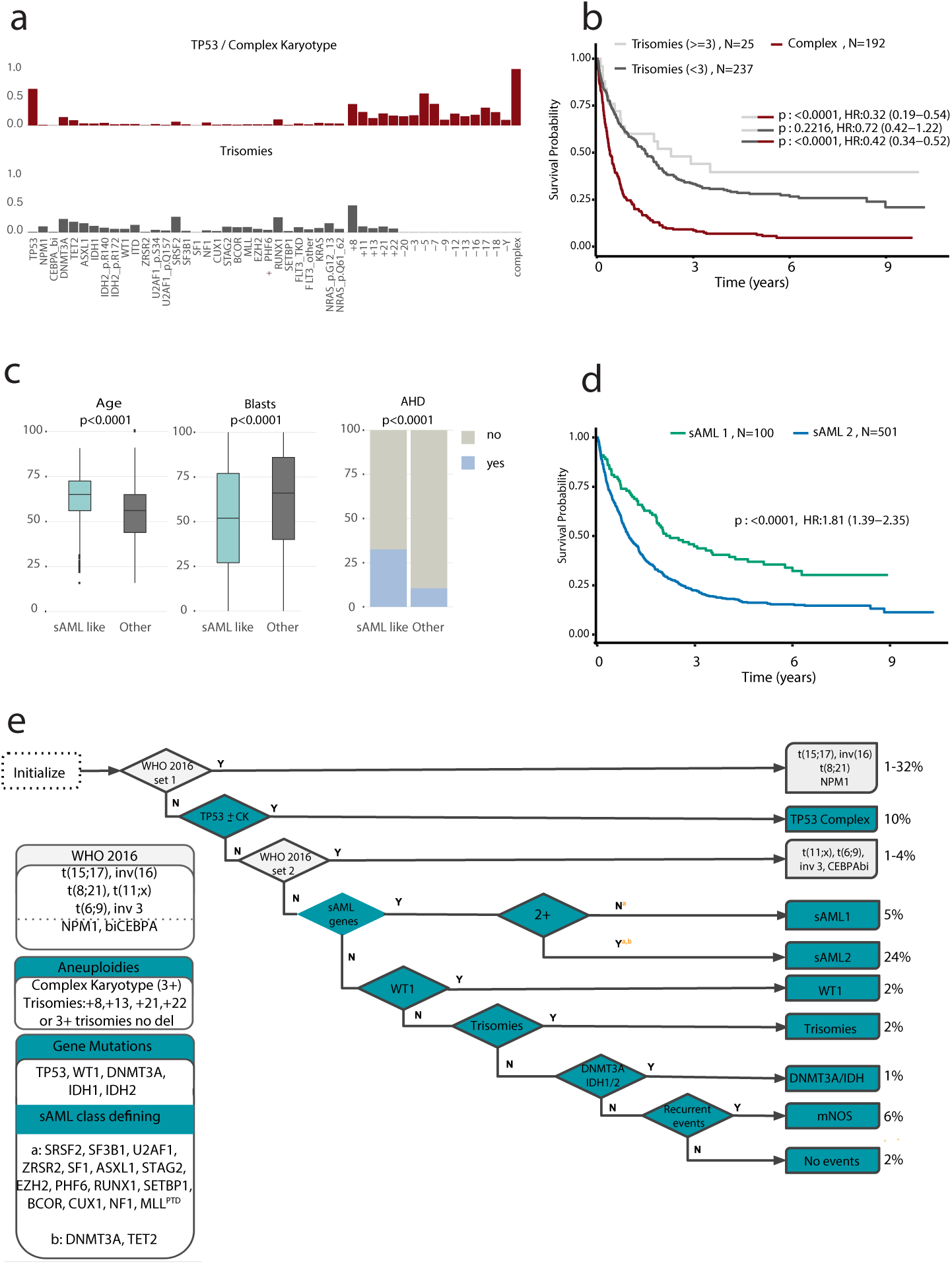
Molecular classification in AML. (**a**)Repartition of 2 patterns of chromosomal aneuploidies to include TP53 and complex and trisomies. The y axis represents the fraction of patients carrying each driver event (on the x-axis) for each of the 2 subgroups (training, n= 2,113). (**b**)Kaplan-Meier overall survival curves for overall survival curves for patients with trisomies (<3)(grey), trisomies(>=3)(lightgrey) and complex karyotype (burgundy) in the training cohort (n= 2,113). Log-rank tests compared the survival distributions between complex and MDS related cytogenetics and between complex and trisomies not complex subgroups. (**c**)Comparison of age (years), bone marrow blasts (%) and AHD (antecedent hematologic disorder) distributions for sAML like subgroups to other AML in AML NCRI cohort. P values on the boxplots used either a Wilcoxon rank-sum test or a Fisher’s exact test. (**d**)Kaplan-Meier overall survival curves for the secondary AML like classes (sAML1 and sAML2) in the training cohort (n=2,113). (**e**)Hierarchical classification schema. Hierarchy rules for AML class assignment, biomarkers for hierarchy implementation and class range proportions. sAML2 comes before biCEPBA in the hierarchy (S.Appendix for more details). WHO 2016 set1 and WHO 2016 set2 display classifications for more than one group. For those 2 specific boxes, we displayed range values representing the proportions of the smallest class and largest class in that subset. For all other sets, the values represent the proportion of patients in the cohort for that particular class.

Patients with ≤2 aneuploidies (n=233, 11%), enriched for “MDS-related”^16,17^ cytogenetic abnormalities clustered with secondary AML type mutations (sAML)^16^ such as *SRSF2, SF3B1, U2AF1, ZRSR2, ASXL1, EZH2, BCOR*, or *STAG2*, as well as novelly described here, *RUNX1, SETBP1*, and *MLL*^*PTD*^ mutations. This represented the second largest cluster (28.4%, n=601). Patients in this group were older (median diagnostic age 65.5 vs 56, p< 0.0001), with lower blast counts (median 51 vs 65, p< 0.0001) and higher incidence of antecedent hematologic disease (AHD) (32% vs 11.4% p<0.0001) (Figure 1C). Given the enrichment of MDS-related abnormalities and sAML like gene mutations, we name this cluster “sAML” per Lindsley et al^18^. Of prognostic importance, the association with adverse outcomes was specific to patients with ⩾2 mutations (5-year survival rate = 16%), as compared to patients with a single gene mutation in a class-defining gene (5-year survival rate = 37%) (Figure 1D,Extended Figure 5, S.Figure 7, S.Table 6). Thus, we subdivided this cluster into secondary AML Like-1 (sAML1) defined by patients with single mutations (n=100) and secondary AML Like-2 (sAML2) for patients with ⩾2 class defining genes (n=501) (Figure 1D, Extended Figure 5, S.Figure 7). AHD was enriched in sAML2 and associated with even worse outcomes (p<0.0001) (S.Figure 8). The provisional WHO entity defined by *RUNX1* mutations (13.5%) spread across both sAML1 and sAML2 subgroups at similar frequencies and did not confer independent prognosis (S.Figure 9).

In the absence of classifying events (e.g CEBPAbi, t(8;21)), *WT1* mutations defined a novel cluster (n=40, 2%) (S.Figure 10) characterized by few mutations, younger diagnostic age (median=41), high white blood cell (WBC) counts and intermediate risk, unless co-mutated with FLT3^ITD^(S.Figure 10). We further validated the *DNMT3A*/*IDH* class in 1% of patients and demonstrate that this is a heterogeneous group (S.Figure 11)^14^. 6% of patients (n=124) not clustering with any class were labelled as “molecularly Not Otherwise Specified” (mNOS) and 2% had no identifiable mutation (n=46) in our panel (no mutations).

These findings informed a hierarchical classification that explicitly assigns 100% of patients into a molecular class (including mNOS and no-mutations) (Figure 1E, S.Figure 12, https://www.aml-risk-model.com/supplementary). Patients in the novel sAML1, sAML2, *WT1* and trisomy classes demarcated independent prognostic groups relative to ELN^2017^ (Figure 2A, S.Figure 13).

**Figure 2:**
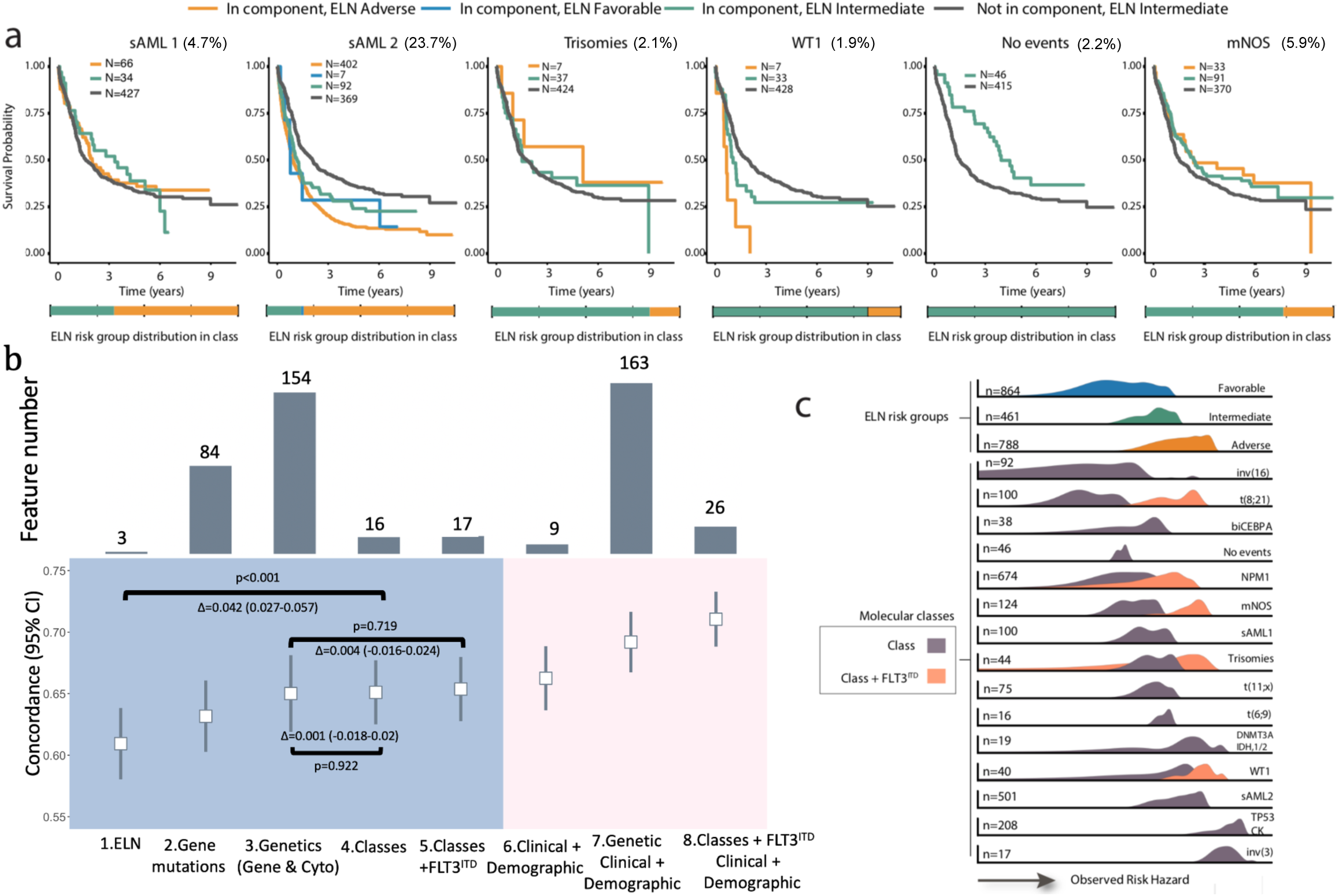
Prognostic relevance of molecular subgroups. (**a**)Kaplan-Meier overall survival curves for the sAML 1, sAML 2, trisomies, WT1, no event and mNOS subgroups, separated by ELN^2017^ scores. A bar plot representing ELN^2017^ repartition for each class is included in the lower panel. (**b**) Estimates of the concordance index (C-index) derived from Cox regression with a ridge penalty that consider 1. ELN^2017^ strata, 2. Gene mutations, 3. molecular classes, 4. molecular classes + FLT3^ITD^, 5. Genetic data (gene mutations and cytogenetics), 6. Clinical and demographic, 7. Genetic, clinical and demographic and 8. Classes, FLT3^ITD^, clinical and demographic features using internal 5 fold cross-validation for penalty selection. Top panel includes bar plots representing the number of features/categories considered in each model (i.e. 3 for ELN). **(c)**Density plots representing the scaled observed hazard (0-1) for the ELN^2017^ risk categories and the proposed molecular classes. In purple we show the density of risk for each class, in orange we present the subset of cases in class that also have FLT3^ITD^. We omitted the density plot for class t(15;17) due to small numbers. The hazard is depicted for overall survival.

This is important as in the absence of risk stratifying biomarkers a significant proportion of patients in these newly defined groups were considered as intermediate risk AML (eg. 18% of patients in the AML2 class) (Figure 2A). Patients with no-mutations had favorable outcomes and were distinct from intermediate-risk mNOS. This demonstrates that given a comprehensive workup, negative findings also provide relevant prognostic information (Figure 1E, 2A, S.Figure 13). Proposed class associations were validated in AMLSG (S.Figure 14). As expected, non-intensively treated patients in the NCRI cohort were enriched in the TP53-CK and sAML2 groups. Nonetheless, the associations with adverse outcomes remained, as in the intensively treated subsets (S.Figure 15).

Notably, similarly to most signaling gene mutations (e.g. *NRAS*), FLT3 mutations are present across classes. Thus, these mutations are not “class defining” and are therefore not considered in the hierarchical classification schema.

### Integration of AML classes into prognostic models for clinical management

Prompted by the strong associations between class and outcomes, we compared prognostic models that considered genetic features to class-based models (S.Table 7-8, Online Methods) or both, using ELN^2017^ as a reference. Monosomal karyotypes^19^ did not provide independent prognostic value^17^ (S.Figure 16). Model comparison demonstrates that a simple model, based on class membership and FLT3^ITD^ status (17 features), captures the same prognostic information as more complex genetic models (154 features)(Figure 2B, S.Figure 17-23, Extended Figure 6). These findings provide a rationale for the development of a risk stratification schema that is based on class membership and FLT3^ITD^ status, thus offering the opportunity to unify classification to risk stratification and importantly link a biological definition of disease ontology to clinical presentation and outcomes.

In agreement with prior findings^7^, inclusion of clinical features (age of diagnosis, gender blast, antecedent hematologic disorder, performance status, white blood cells, hemoglobin and platelet) achieved the highest improvement in model discrimation (Figure 2B, S.Figure 24). Figure 2C exemplifies how the heterogeneity in clinical outcomes (as a function of overall survival hazard) is captured by the proposed classification. Despite differences in age, geography and chronology, feature selection was comparable in the AMLSG cohort, indicating that results are generalizable across AML patients (S.Figure 25-26) and further demonstrating class membership as stable features for prognostic model construction in AML.

### A multi-state model for disease progression

We next studied associations between class membership, treatment response and relapse. Modelling a patient’s journey through treatment, we applied a six-state Markov Model (MM)^20^ that includes the following states: alive (n=2017); alive in CR (n=1460); relapse (n=778); death without CR (n=543); death with CR (n=199) and death following relapse (n=607) (Figure 3A, S.Figure 27). Results were consistent in the intensively treated subset (n=1661) (S.Figure 28).

**Figure 3:**
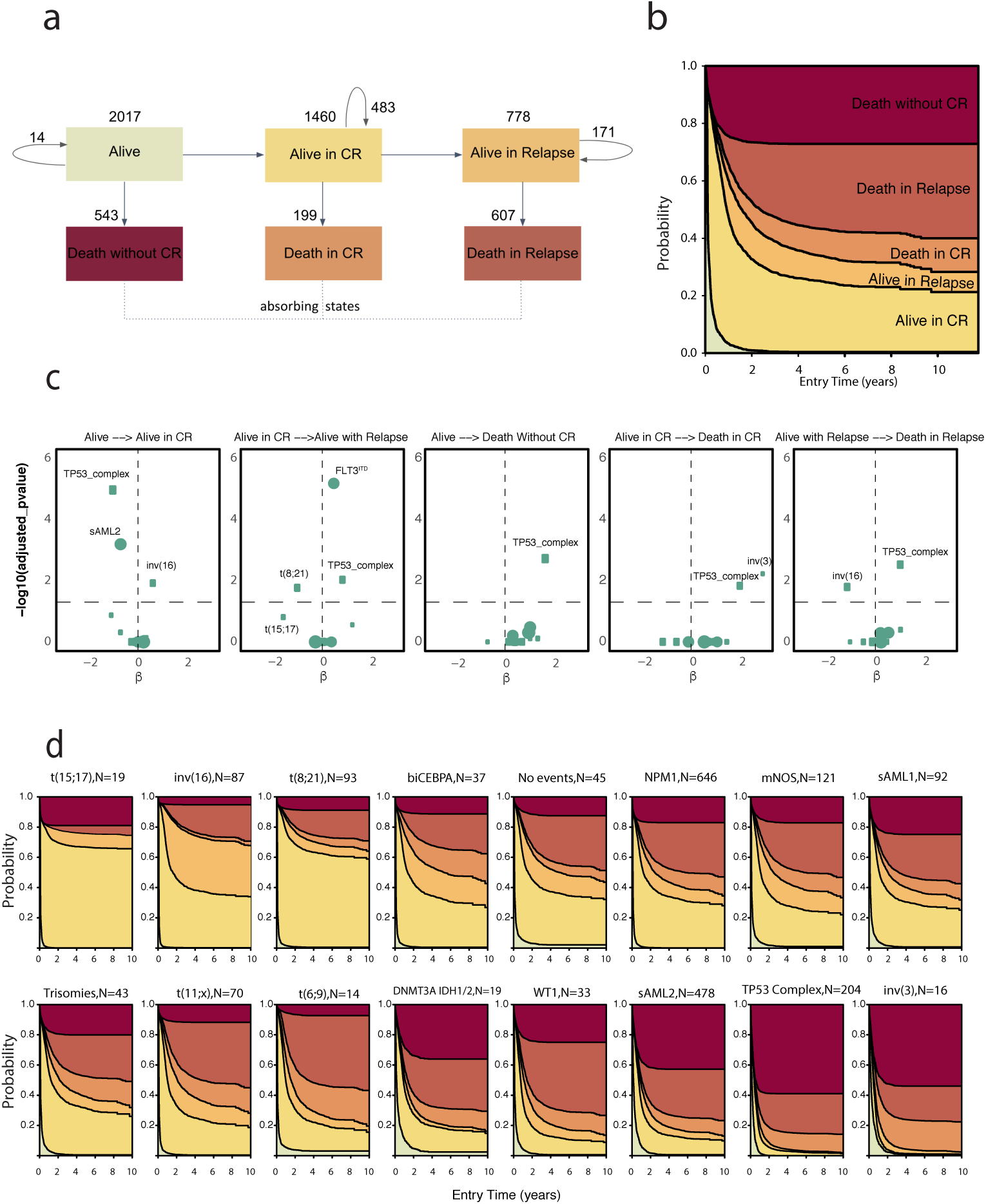
Multi-state model for disease progression in the AML NCRI Cohort (n=2,017). **(a)**Representation of patient transitions (in numbers) across clinical endpoints (alive (meaning received induction chemotherapy); alive in complete remission; alive in relapse; death without complete remission; death in complete remission; death in relapse).The arrows represent the number of transitioning patients. Circle arrows correspond to number of patients that do not transition. **(b)**Stacked transition probabilities (y-axis) across time (x-axis). **(c)**Cox volcano plots depicting the association between state transitions and molecular classes and FLT3^ITD^. The horizontal dotted curve corresponds to the p-value threshold of 0.05 and the vertical one corresponds to β=0 on the x axis. Highlighted predictors have a significant effect or have large β coefficients (p-value greater than the threshold: 0.05 here or p-value close to threshold and |β|>1.5). The size of each point corresponds to the frequency of the event. The reference class in the Cox transition models is no events. **(d)** Stacked transition probabilities for each class (y-axis) across time (x-axis). The bold lines represent the transition states. We omitted n=96 patients from the multi-state model for disease progression (2,113-96=2,017) due to missing timepoints.

This analysis provides the first detailed map of the proportion of patients in each class likely to transition between any two clinical endpoints over time (e.g Alive in CR -> Death in CR, or Alive in CR -> Relapse ->Death in relapse). The resulting survival estimates reflect the cumulative hazard for each of the transitions (Figure 3B-D). This provides further resolution to established associations and provides novel insights. For example, patients with inv(16) or t(8;21) have comparable OS estimates, yet patients with inv(16) are more likely to relapse^21^ (S.Figure 29-30). Notably, upon relapse, inv(16) patients achieve the highest salvage frequencies, as compared to all other AML classes. Patients with no events, considered as intermediate-risk, have similar transitions to the *NPM1* class. We estimate endpoint-specific outcomes for the novel *WT1*, Trisomy and mNOS classes, which together with t(6;9) respond well to induction chemotherapy. However, patients in *WT1* and t(6;9) class have a high likelihood of relapse related mortality (Figure 3B-D). This is particularly the case for the subset of patients with FLT3^ITD^. Indeed, subjects with FLT3^ITD^ had both decreased likelihood of achieving CR, and increased risk of relapse-related mortality across all AML classes, not just *NPM1* (S.Figure 29-30). This is despite the use of escalated doses of daunorubicin in AML17 which has been reported to reduce relapse risk in patients with FLT3^ITD 22^. Furthermore, this model demonstrates that a key differentiator between sAML1 and sAML2 is response to induction chemotherapy, with 43.7% of sAML2 group patients not attaining CR as compared to 26% in sAML1 (Fisher’s Exact test p=0.002). Consistent with prior findings, adverse outcomes in TP53/complex and inv(3) are explained by highly chemoresistant disease and relapse-related mortality^14,23^. These observations were also observed in the AMLSG cohort (S.Figure 31) (S.Table 9).

### Implications for disease surveillance

MRD surveillance assesses initial response and guides treatment decisions^24,25^, such as HSCT. Whilst MRD status is considered an independent predictor of outcome^26,27^, the predictive relevance of MRD has not been determined across classes.

Results from MRD surveillance by flow-cytometry after course 1 were available in 523 UK-NCRI AML17 patients^16^. Of these, 202 were CR MRD^-ve^ and 321 were CR MRD^+ve^ (Figure 4A, S.Figure 32). The MRD^+ve^ rate, by class, ranged from ∼33% to 95% (Figure 4B). As expected^6^, MRD^+ve^ patients had a higher risk of relapse and death (Figure 4A), with some exceptions. 70% (MRD^+ve^=69) of sAML2 patients in CR were MRD+, yet while there was no evidence of a significant difference in relapse or survival rates, there was no difference of effect by group (p=0.3 for interaction) (Figure 4C, S.Figure 33). For sAML1, there was no difference in relapse-incidence between MRD^+ve^ and MRD^-ve^ subjects. A trend towards poorer OS for MRD^+ve^ patients was observed. These results suggest that while achieving MRD-negativity after the first course is associated with favorable outcomes, its utility may not be universal across classes (S.Figure 34-37) and that differences may be explained by the underlying biology associated with the mutations in each class.

**Figure 4:**
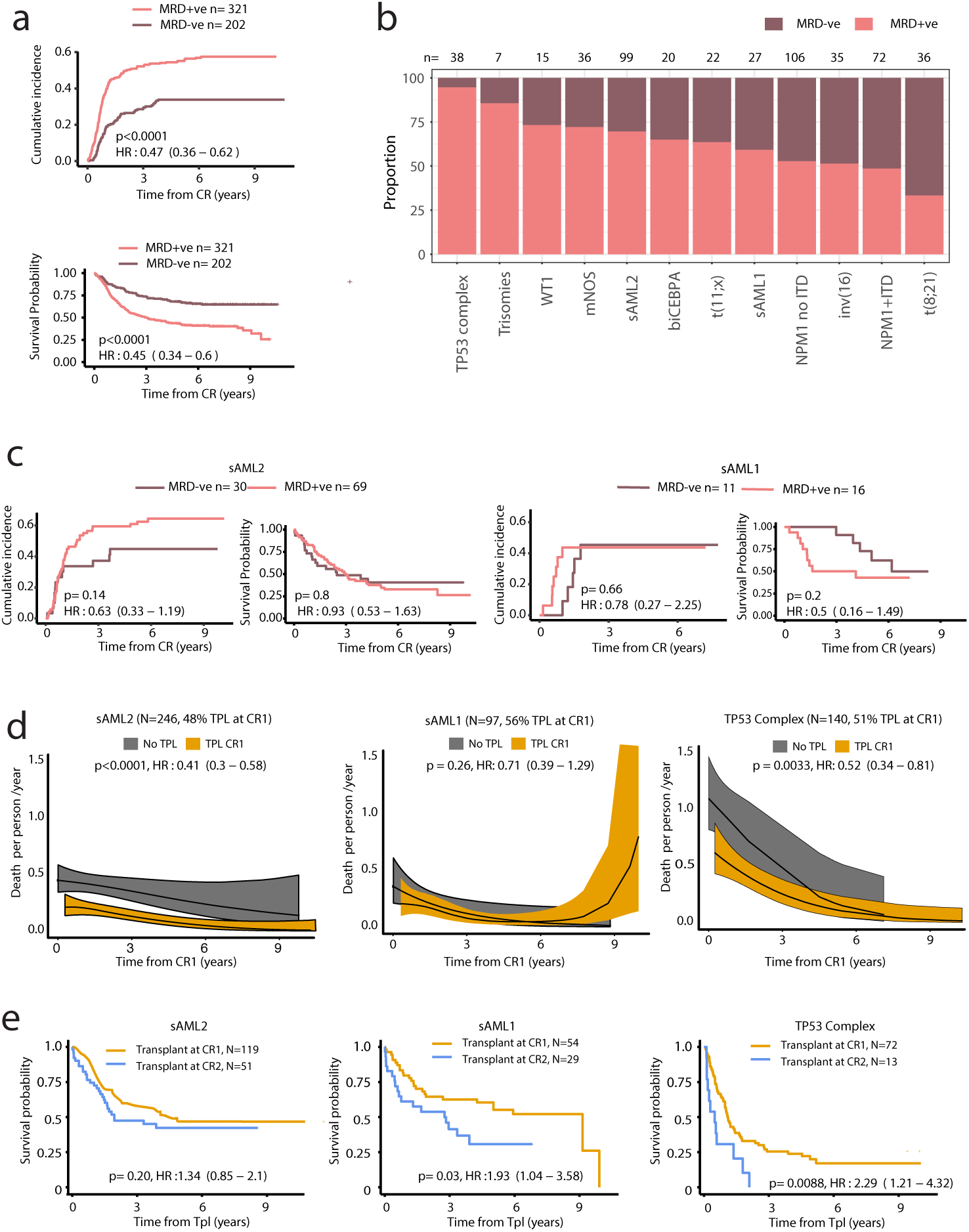
Implications for measurable residual disease surveillance and transplant outcomes. **(a)**Cumulative incidence of relapse and Kaplan-Meier overall survival curves for patients that attained CR in AML17 trial subset, stratified by MRD status post course 1 (n=523). Gray’s test and the logrank test were used to compare the relapse incidence and survival, respectively. **(b)**Barplots indicating proportion of patients in each molecular class with flow MRD+ve (any detectable MRD) or MRD-ve status post course 1. Restricted to the AML17 trial subset (n=523) and to classes with at least 5 patients in the MRD+ve subset. **(c)** Incidence of relapse and OS by MRD status for the sAML 2, sAML1 subgroups. A test for interaction between sAML1 vs sAML2 and MRD (Interaction HR: 1.90 (0.55-6.49), p-value: 0.31) was not significant. The analysis provided in figure 4c is limited to AML17 patients with MRD data available. **(d)** Nonparametric estimated curves of the hazard rate (deaths per person-year;y-axis) across time (x-axis) for the sAML2, sAML1 and TP53 complex subgroups in the combined dataset (UK-NCRI and AMLSG). Curves display the hazard for patients transplanted (TPL) in CR1 to the non-transplanted patients. Tests of association were modeling transplant as a time-dependent covariate adjusted for age and performance status. A test for interaction between sAML1 vs sAML2 and transplant was borderline significant (Interaction HR: 0.57 (0.30-1.08), p-value: 0.08). **(e)**Kaplan-Meier overall survival curves comparing patients who have been transplanted in CR1 to patients transplanted in CR2 for the selected classes. P-values are computed using the log-rank test. The analysis in figure 4d-e is limited to the patients to 2,244 intensively treated patients in the UK-NCRI (n=1,095) and AMLSG (total n=1,149) that achieved CR, 759 patients were transplanted in CR1 and 436 after relapse (Total n=1,195).

### Relevance of AML classes to transplant outcomes

Next, we evaluated HSCT outcomes by AML class. Consolidating data from 2,244 intensively treated patients in the UK-NCRI (n=1,095) and AMLSG (total n=1,149) that achieved CR, 759 patients were transplanted in CR1 and 436 after relapse (Total n=1,195) (S.Table 10-12).

Evaluation of OS with respect to class and HSCT timing (Figure 4D-E) demonstrated that sAML2 patients undergoing HSCT had a reduced risk of death, following adjustment for performance score and age (p<0.0001; Figure 4D). There was no significant survival difference based on HSCT in CR1 or CR2 (p=0.21; Figure 4E). Of note, patients in the TP53-CK also benefited from HSCT. However, in this group, patients transplanted in CR2 had significantly worse survival than those transplanted in CR1 (p=0.009; Figure 4E, Extended Figure 7). Adjusting for age and performance score, HSCT was not associated with a reduced risk of death for patients in the sAML1 group, albeit there was evidence of a benefit for patients transplanted in CR1 vs CR2 (Figure 4D-E, Extended Figure 7).

Prior studies show that adverse risk groups as defined by cytogenetics benefit from transplant^28^. Here our findings extend the definition of adverse risk groups to include the newly defined sAML2 group, which account for 23.7% of patients in the study. However, given inherent selection biases associated with transplant, these results warrant validation in prospective studies.

### Relevance of molecular class in AML risk stratification

Using a panel of 32 genes (S.Table 13), the proposed classification explicitly assigns 92% of patients into one of 14 AML classes and is sufficient to classify remaining patients into the two mNOS or no events subgroups. We demonstrated that class membership and FLT3^ITD^ status, capture the same prognostic information as genetic parameters. We next assessed how a class-based framework might inform future ELN^2017^ revisions.

Using the ELN^2017^ as a foundation, we assigned each class to one of three proposed risk strata (Favorable^p^, Intermediate^p^, Adverse^p^) (Figure 5A). Patients in the no events class were assigned to Favorable^p^ group and “mNOS’’ patients to the Intermediate^p^. sAML1, trisomies, *WT1, DNMT3A/IDH* and t(6;9) were classified as Intermediate^p^-risk and sAML2 as Adverse^p^. Per ELN^2017,^ t(6;9) was considered an adverse-risk group. We show that adverse-risk is specific to the subset of patients (72%) with FLT3^ITD^ (Extended Figure 8). We demonstrate that FLT3^ITD^ is the only gene that delivers independent prognostic information from class membership. ,Indeed, the presence of FLT3^ITD^ was associated with worse outcomes for all intermediate-risk (Figure 5B, S.Figure 38) classes. This association was independently of FLT3^ITD^ ratio^29,30^ (S.Figure 39). Thus, FLT3^ITD^ status was used to upgrade risk for all intermediate-risk patients to adverse-risk.

**Figure 5:**
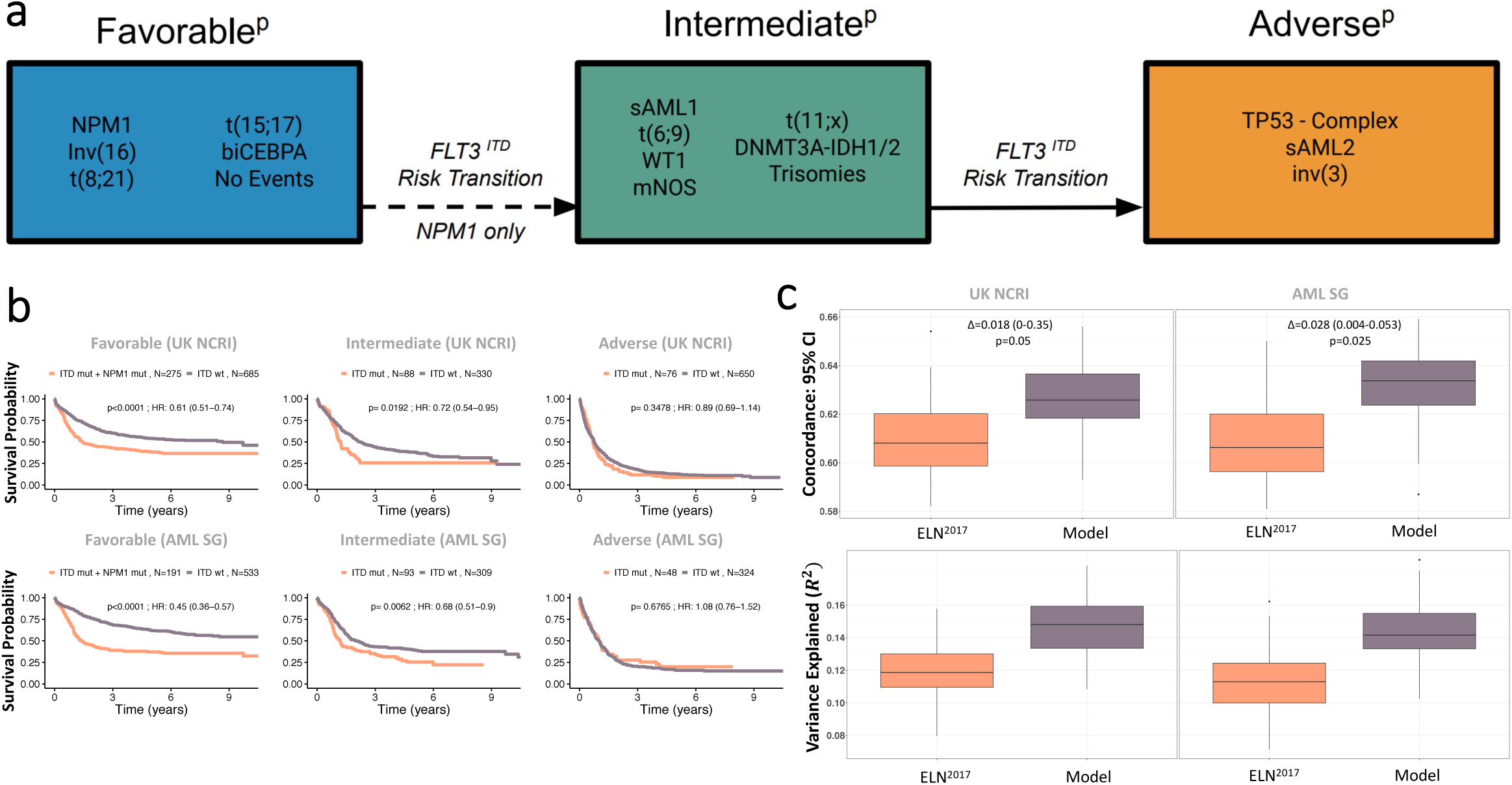
Establishment of a new risk proposal based on the AML classes. **(a)**Class assignment into one of three proposed risk categories (Favorable^P^, Intermediate^P^, Adverse^P^) is based on class membership and FLT3^ITD^ status, whereby the presence of NPM1 and FLT3^ITD^ in the Favorable^P^, the presence of FLT3^ITD^ in the Intermediate^P^ groups shifts one risk category to the Intermediate^P^ and Adverse^P^ respectively. Patients initially classified as favorable with NPM1 and FLT3^ITD^ shift to intermediate. Patients classified as intermediate by class membership with the presence of ITD shift to adverse. The dotted arrow refers to the risk transition for patients with both NPM1 and FLT3^ITD^ mutations from favorable to intermediate. The solid arrow refers to the risk transition for patients with FLT3^ITD^ from intermediate to adverse. **(b)**Kaplan-Meier overall survival curves comparing each of the proposed risk strata (Favorable^P^, Intermediate^P^, Adverse^P^) by the presence of NPM1 and FLT3^ITD^ status for the Favorable^P^ and by FLT3^ITD^ status for the Intermediate^P^ and Adverse^p^ in the training AML NCRI cohort (n=2,113) and the validation AML SG cohort (n=1,540) validate the rationale for the FLT3^ITD^ shift in risk. P values were computed using the log-rank test. **(c)**The estimated improvement in the concordance index (C-index) and pseudo-variance explained (R^2^) for the two classifiers in the training AML NCRI Cohort (n=2,113) and validation AML SG Cohort (n=1,540). 95% confidence intervals were generated by bootstrap resampling for the C-index.

Taken together, this framework re-stratified 25.5% of NCRI and 24.6% of AMLSG patients (Extended Figure 9, S.Figure 40). The redefined risk-strata overlapped with ELN^2017^ trajectories. However, the proposed framework led to an increase in variance explained and a significant improvement in the c-index (p = 0.05 for NCRI; p=0.025 in AMLSG) (Figure 5C, Extended Figure 10, S.Table 8). The relative proportion of transplanted patients did not differ amongst the respective ELN strata (S.Table 12) and results were consistent in the intensively treated subset (n=1755) (S.Figure 41).

### Clinical decision support tools

Appreciating the complexity introduced by the multitude of genetic features considered, we developed a web-based tool that executes the proposed classification and risk stratification hierarchy (Figure 1E, Figure 5A) (Figure 5-6, https://www.aml-risk-model.com/calculator). Using mutations in 32 genes and cytogenetics as input variables, supervised classification assigns each patient into the corresponding AML class and risk group. The model is restricted to the intensively treated subset of the study (n=3201), which represents 90% of the patients in both NCRI and AMLSG cohorts. Graphical representation of end-point specific predictions across time are presented in the form of sediment and barplots. The contributing factor tab displays patient specific covariates that inform each transition estimate alongside the corresponding coefficients. For example, Patient PD25176a, classified as intermediate risk per ELN^2017^ is in their 60s with normal karyotype and mutations in *BCOR* and *SF3B1*. Here, this patient classifies as sAML2 with Adverse^P^ risk and the predicted outcomes for each transition are displayed (Figure 6, S.Figure 42). To account for cases with clinical presentation outside the 95th quantile range of the training cohort and enhance interpretability of results, we introduce a warning sign for outlier cases and compute confidence intervals for all predictions in the calculator.

**Figure 6:**
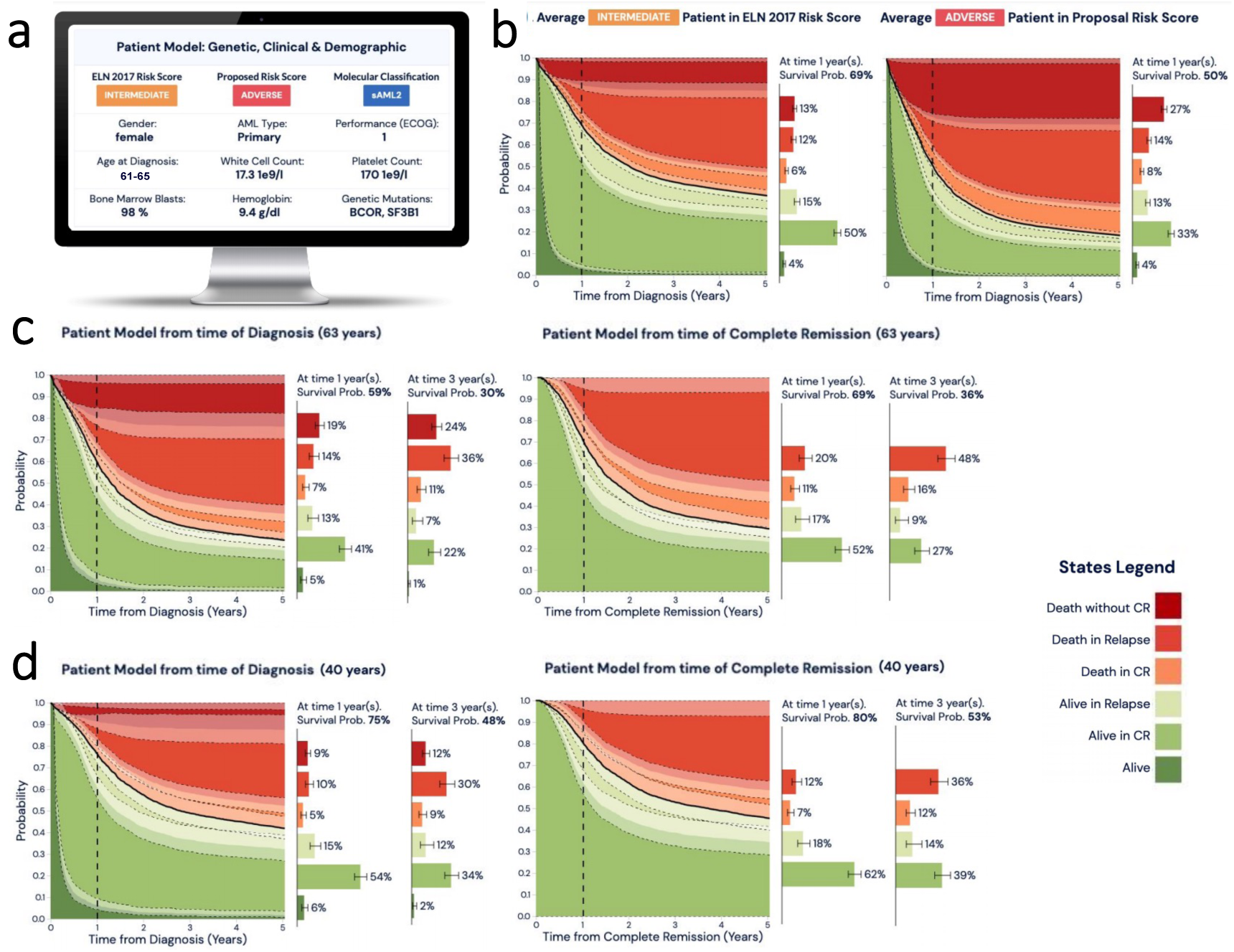
Example presentation of personalized clinical decision support tool for molecular classification and risk stratification. The calculator is derived using the multi-state models that consider data from (n=3,201 total patients, UK-NCRI and AMLSG) all intensively treated. **(a)**Input parameters to include cytogenetic, genetic, clinical and demographic are considered to **(b)**display each patient’s ELN^2017^ score alongside with the proposed molecular class and proposed risk group developed in this study. **(c**,**d)**Adjacent barplots show the relative contribution of each covariate (molecular, clinical, demographic) on each transition. Estimates can be dynamically derived for the time of diagnosis or upon attainment of CR and across timepoints (i.e. Year 1 post diagnosis or CR, Year 3 post diagnosis or CR etc). To further improve interpretation confidence, we provided confidence intervals for each sediment plot and probability estimate transitions. Precise age is replaced by 5-year range to remove identifying informations.

## Discussion

The scale and comprehensive analyses deployed in this study enabled us to validate some of the findings in the prior literature, establish novel insights and consolidate these into a global framework for the introduction of molecular biomarkers in clinical algorithms for AML patient management.

Using data from 3,653 patients, we develop and validate a unified framework for disease classification and risk-stratification in AML that is informed by cytogenetics and 32 genes. This framework classifies 100% of AML patients into one of 16 molecular subgroups and refines our understanding of established classes (e.g t(6;9)), as well as provisional WHO entities (e.g RUNX1). We identify novel clusters of prognostic relevance (sAML1, sAML2, WT1, trisomies) accounting for 33.3% of AML patients, demonstrate the importance of negative molecular findings (No events, mNOS) and highlight the broad implications of FLT3^ITD^-positivity irrespective of FLT3^ITD^ allelic ratio.

Implementation of multistate models that consider each transition during a patients journey through AML, such as attainment of CR, likelihood of relapse and risk of death fine-map the most likely temporal trajectories for the established classes, and importantly further dissect and add granularity to the novel classes, which were previously merged into a heterogeneous unknown or intermediate-risk group. This provides a blueprint linking the biological processes deregulated within each class to a patient’s likelihood of response to treatment, risk of disease progression, relapse and death. As an exemplar of this added clarity, the novel sAML2 class accounts for 24% of AML patients, is associated with chemorefractory disease, high relapse rates and poor survival, irrespective of early MRD negativity. However, for the subset of sAML2 patients who achieve CR, there appears to be a benefit of HSCT. Future studies, powered by adequate sample size are warranted to confirm these observations.

Building upon the ELN^2017^ guidelines, we propose a three-tier risk-score (Favorable^P^, Intermediate^P^, Adverse^P^) informed by class membership and FLT3^ITD^ status. Informed by the novel classes, this framework restratifies one in four AML patients and achieves significant improvement in prognostic accuracy. Moreover, despite demographic and clinical trial differences between our test and validation cohorts, our findings are reproducible across both the UK-NCRI and AMLSG cohorts. This is likely because the molecularly defined classes capture the spectrum of phenotypic and clinical heterogeneity observed amongst AML patients. Importantly, this demonstrates that findings from this study are generalizable across AML patients and importantly are representative of those seen in routine clinical practice, particularly those considered fit for therapeutic intervention, where clinical decision making is currently most problematic. Despite the emergence of adjunct therapeutic approaches in the management of AML^3,4^, the backbone regiments in our training and validation cohorts (“7+3 like”) are representative of AML treatment practices globally.

Recent correlative studies between molecular biomarkers and clinical outcomes primarily focus on single-genes or broad, heterogeneous risk groups ^13,31–33^. This has challenged integration of findings into generalizable clinical algorithms to guide patient management. More recently, we prototyped patient tailored clinical decision support tools in AML that deliver personalized risk scores^7^. However, in the absence of adjunct risk strata the utility of personalized risk scores in clinical trial design and interpretation of results is limited. Additionally, the complexity and “black box” nature of the models challenge clinical implementation and interpretation. Here, we deliver a simplified framework whereby mutations and cytogenetic findings at >100 loci can be summarized by 16 classes and the corresponding three risk strata. Class membership provides resolution in the heterogeneity observed in clinical presentation and delivers a rationalized schema for correlative studies as compared to single biomarkers or clinical cutoffs (e.g. % blast counts), which may dichotomize or group together heterogeneous and biologically distinct nosological entities. The delivery of a stable, biologically informed classification system further enables future studies of class-specific associations that extend beyond those of single gene-mutations and capture the most common patterns of co-mutation.

Integration of data from MRD and HSCT outcomes allowed us to prototype such correlative analyses using class-based associations. Indeed, the applicability of this framework to emerging treatment approaches in AML management to include less intensive regimens^4^ or emerging therapeutics^3^ needs further definition. Likewise, while we demonstrate a significant increase in risk hazard for many molecular classes in the presence of an additional *FLT3-ITD* mutation, we note that the survival advantage described in recent trials where FLT3 inhibitors were combined with intensive chemotherapy were modest^34–36^. It is noteworthy that patients with established adverse cytogenetics or genetic features (*TP53* mutations) associated with poor outcomes to intensive chemotherapy also have adverse outcomes in response to emerging treatments such as azacytidine and venetoclax^4^. Here we expand the definition of adverse risk to encompass patients with sAML2 and a broader cohort of patients with FLT3^ITD^ mutations.

As clinical management options evolve, studies focused on class associations with MRD status^19^, response to emerging therapeutics^3^ such as FLT3 or IDH^37,38^ as well as combination regimens (e.g azacytidine and venetoclax)^4^ are warranted to make definitive determinations for AML patient care.

## Data Availability

All data produced in the present work are contained in the manuscript and in supplementary files.

https://www.aml-risk-model.com/

## AUTHOR CONTRIBUTIONS

E.P., B.J.P.H, and S.M.D designed the study. Y.T. performed all statistical analysis. S.M.D. and E.P. supervised statistical analysis plan and execution. E.P. B.J.P.H, S.M.D, and N.R, supervised research and coordinated the study. I.T, A.G, S.F, S.J.J, R.H, R.D, A.G, L.B, K.D, O.O, R.A, H.D, A.B, N.R provided clinical data and DNA specimens. A.G., and E.P. coordinated sample acquisition. Y.T, J.A.O, Y.Z, Y.P, M.F.L, D.L. A.B and E.P, performed bioinformatic analysis. A.G, L.B, K.D, H.D, clarified validation cohort queries. Y.T, J.A.O, Y.Z, with input from M.F.L developed web based clinical decision support tool. Y.T. and E.P. prepared figures and tables. Y.T, S.M.D, B.J.P.H and E.P. wrote the manuscript. All authors reviewed and approved the manuscript during its preparation.

## ACKNOWLEDGEMENTS

E.P. is a Josie Robertson Investigator and is supported by the European Hematology Association, American Society of Hematology, Gabrielle’s Angels Foundation, V Foundation and The Geoffrey Beene Foundation and is a Damon Runyon Rachleff Innovator fellow. Work in the BJPH lab is funded by Cancer Research UK (C18680/A25508), the European Research Council (647685), MRC (MR-R009708-1), the Kay Kendall Leukaemia Fund (KKL1243), the Wellcome Trust (205254/Z/16/Z) and the Cancer Research UK Cambridge Major Centre (C49940/A25117). This research was supported by the NIHR Cambridge Biomedical Research Centre (BRC-1215-20014), and was funded in part, by the Wellcome Trust who supported the Wellcome - MRC Cambridge Stem Cell Institute (203151/Z/16/Z). The views expressed are those of the authors and not necessarily those of the NIHR or the Department of Health and Social Care. L.B., H.D. and B.J.P.H. are supported by the HARMONY Alliance (IMI Project No. 116026; https://www.harmony-alliance.eu/). The UK-NCRI AML working group trials were supported with research grants from the Medical Research Council (MRC), Cancer Research UK (CRUK), Blood Cancer UK and Cardiff University. We would like to thank all patients and investigators for their participation in the trials and the study.

## Disclosures and COI

E.P. is a founder and equity holder in Isabl, a cancer whole genome sequencing analytics company.

**Extended Figure 1:**
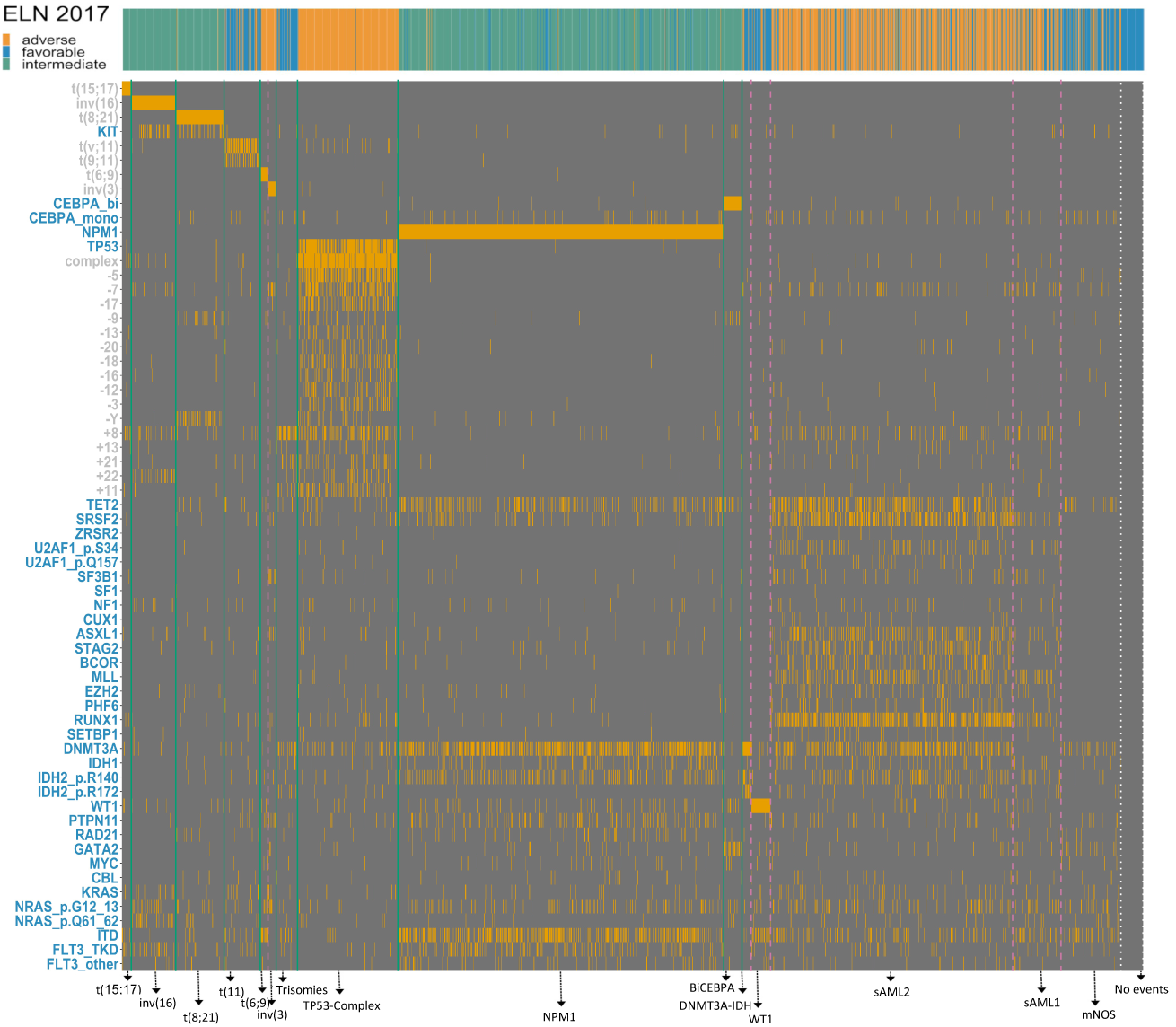
Heatmap of Bayesian Dirichlet Process clusters. Rows demarcate distinct genetic lesions, columns represent individual samples. Orange lines in heatmap indicate the presence of a specific genetic lesion. Green and pink dotted lines demarcate major clusters and sub-clusters as derived by the first and second iteration of the Bayesian Dirichlet Process. Dotted white lines demarcate patients in the molecularly not otherwise specified (mNOS) class and the no recurrent molecular findings. The top sidebar indicates ELN^2017^ risk score for each patient (blue for favourable, green for intermediate and orange for adverse). For more details on the class assignment process, refer to S.Appendix.

**Extended Figure 2:**
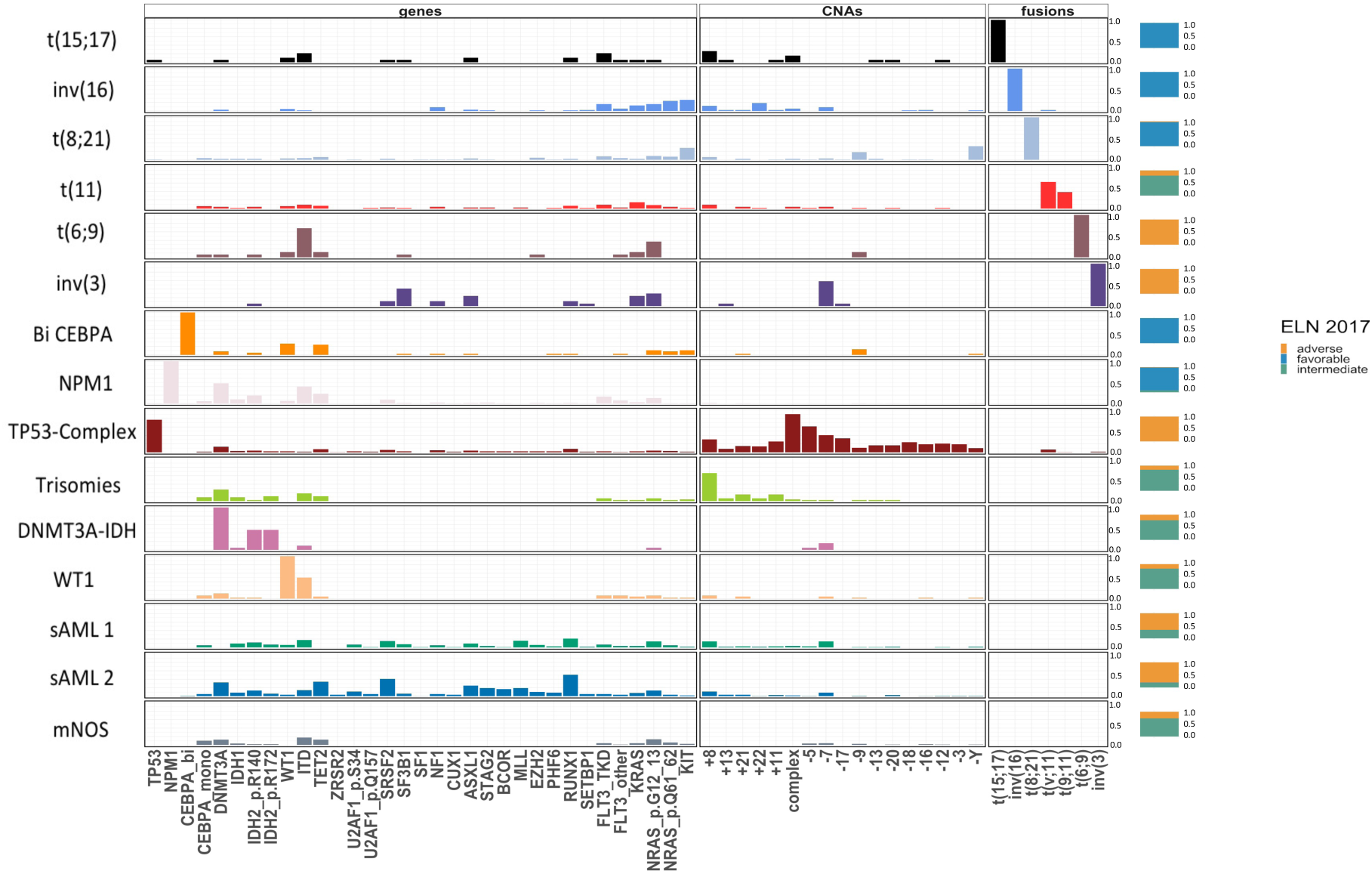
Genetic landscape of AML classes. Co-mutation bar plots for acquired gene mutations, chromosomal aneuploidies and fusion genes for each class. The y-axis represents the fraction of patients carrying the driver event (on the x-axis) within each class. For each class vertical sidebar denoting distribution of patients in class across the three (ELN^2017^) risk strata (blue for favorable, green for intermediate and orange for adverse).

**Extended Figure 3:**
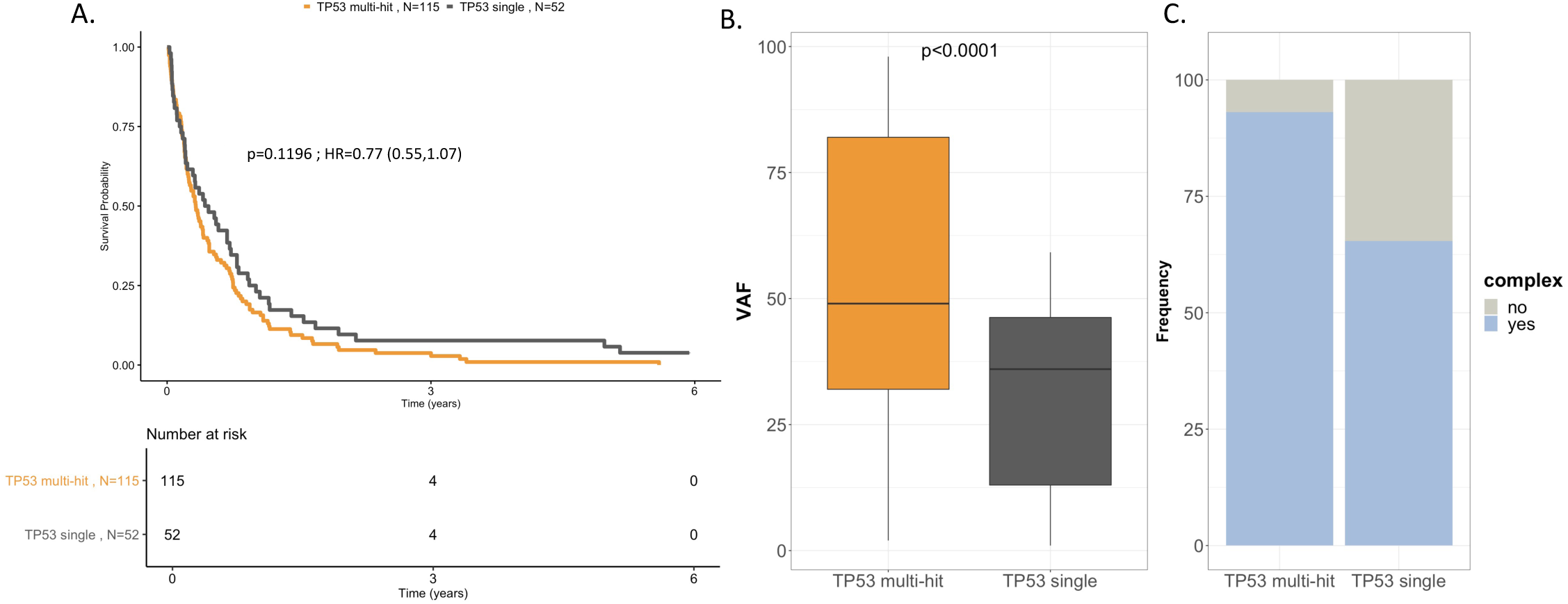
A. Kaplan-Meier Curves for OS and associated risk table comparing TP53 single and multi hit in the AML NCRI cohort (N=2,113). Log-rank tests compared the survival distributions between the 2 subgroups. B. Comparison of variant allele frequency distribution for TP53 single and multi-hit in the AML NCRI cohort (N=2,113). Pvalue was computed with a Wilcoxon rank-sum test. C. Comparison of frequency of complex karyotype patients for TP53 single and multi-hit in the AML NCRI cohort (N=2,113).

**Extended Figure 4:**
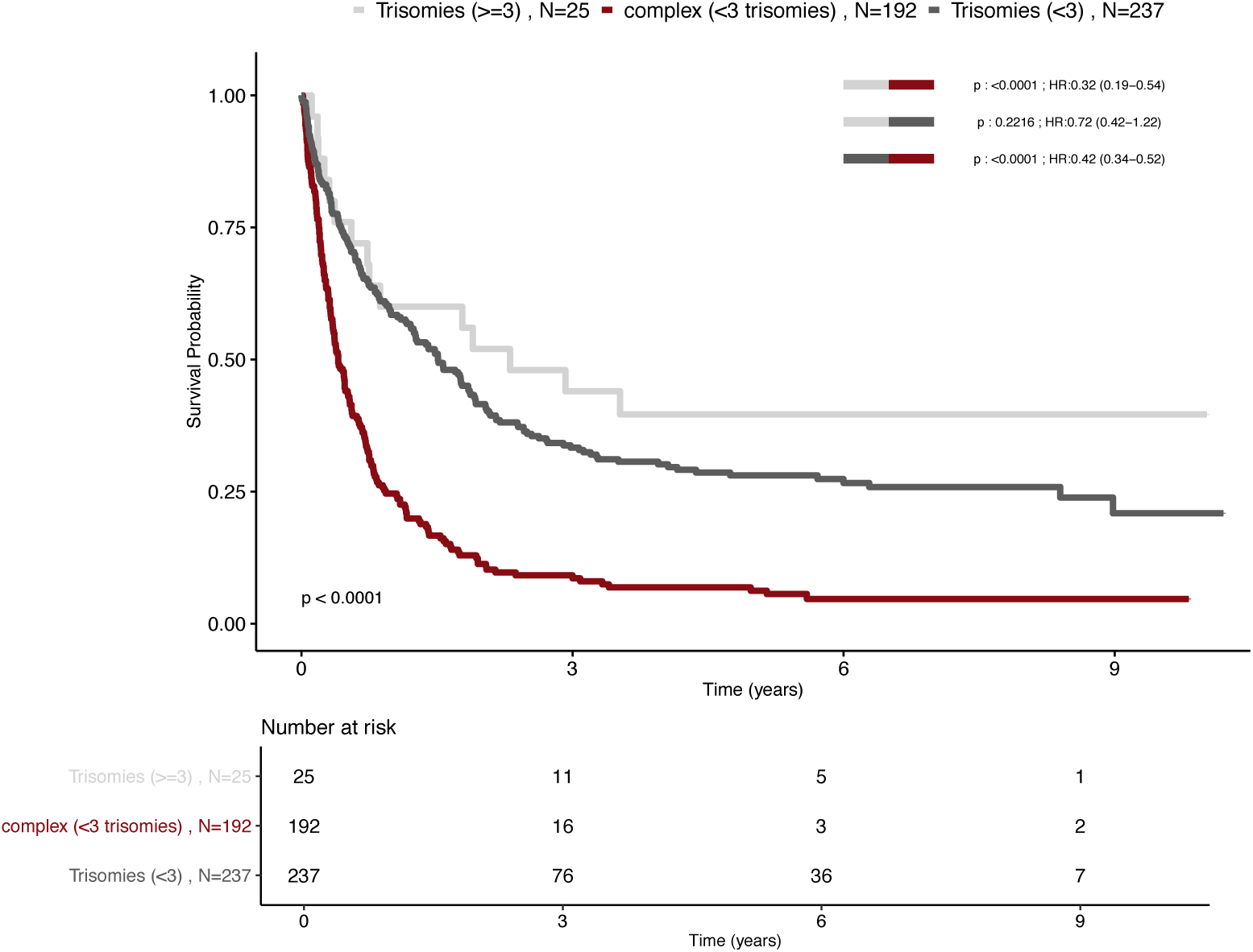
Kaplan-Meier and associated risk table for. overall survival curves for patients with trisomies (<3) (grey), trisomies (>=3) (lightgrey) and complex karyotype (burgundy) in the AML NCRI cohort (N=2,113). Log-rank tests compared the survival distributions between the 3 possible combinations of 2 subgroups.

**Extended Figure 5:**
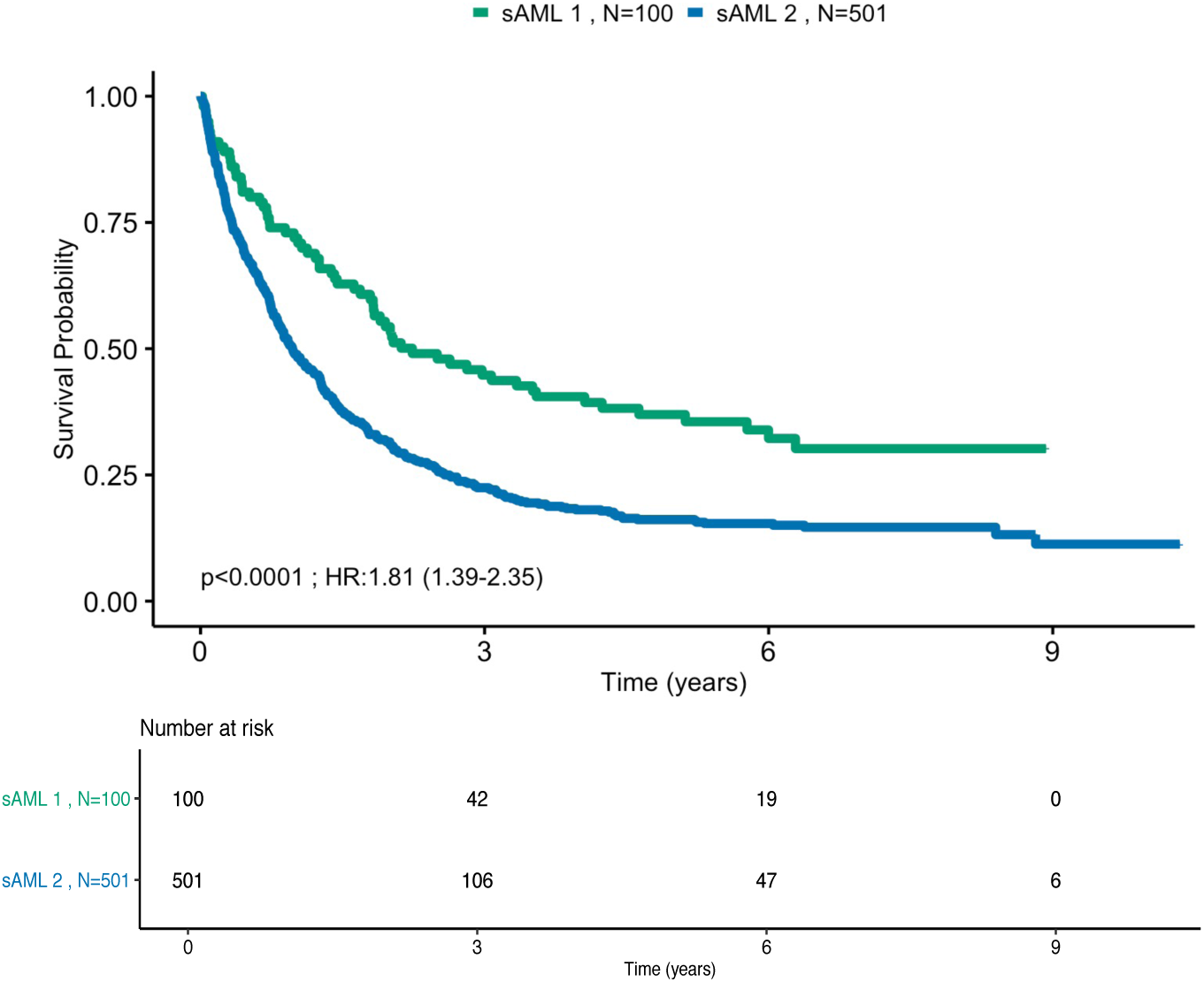
Kaplan-Meier and associated risk table for. overall survival curves for the 2 secondary AML like classes in the AML NCRI cohort (N=2,113). Log-rank tests compared the survival distributions between the 2 subgroups.

**Extended Figure 6:**
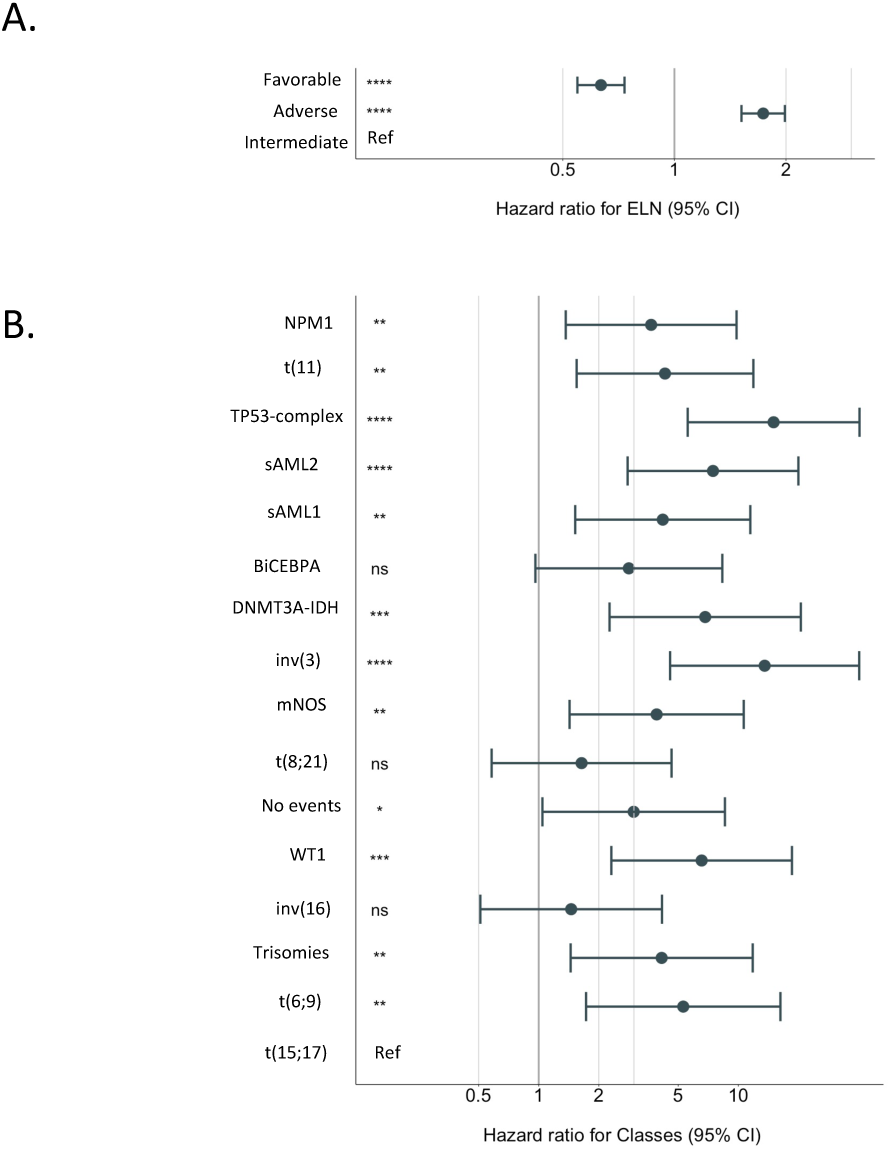
Forest plot multivariate Cox regression of. A. ELN^2017^ risk categories and B. Classes in NCRI trial study set (n= 2,113).

**Extended Figure 7:**
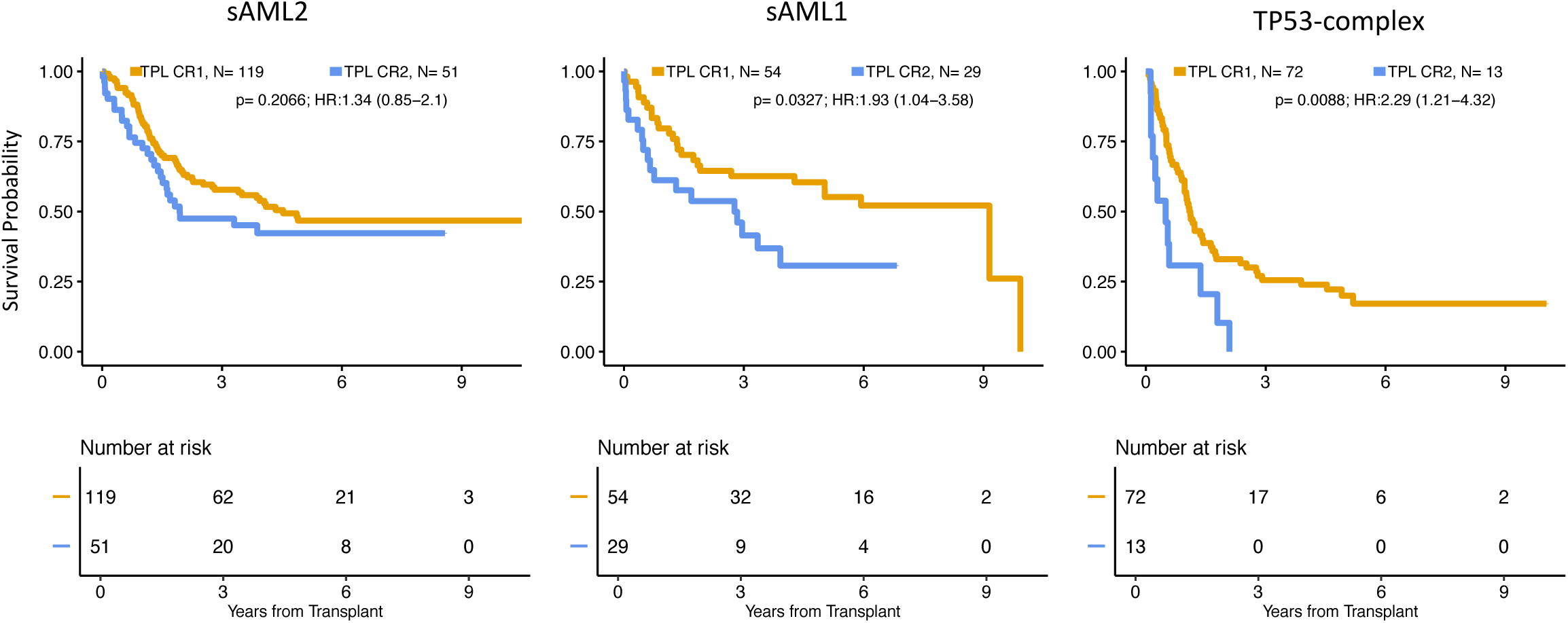
Kaplan-Meier overall survival curves and associated risk tables comparing patients who have been transplanted in CR1 to patients transplanted in CR2 for the selected classes.

**Extended Figure 8:**
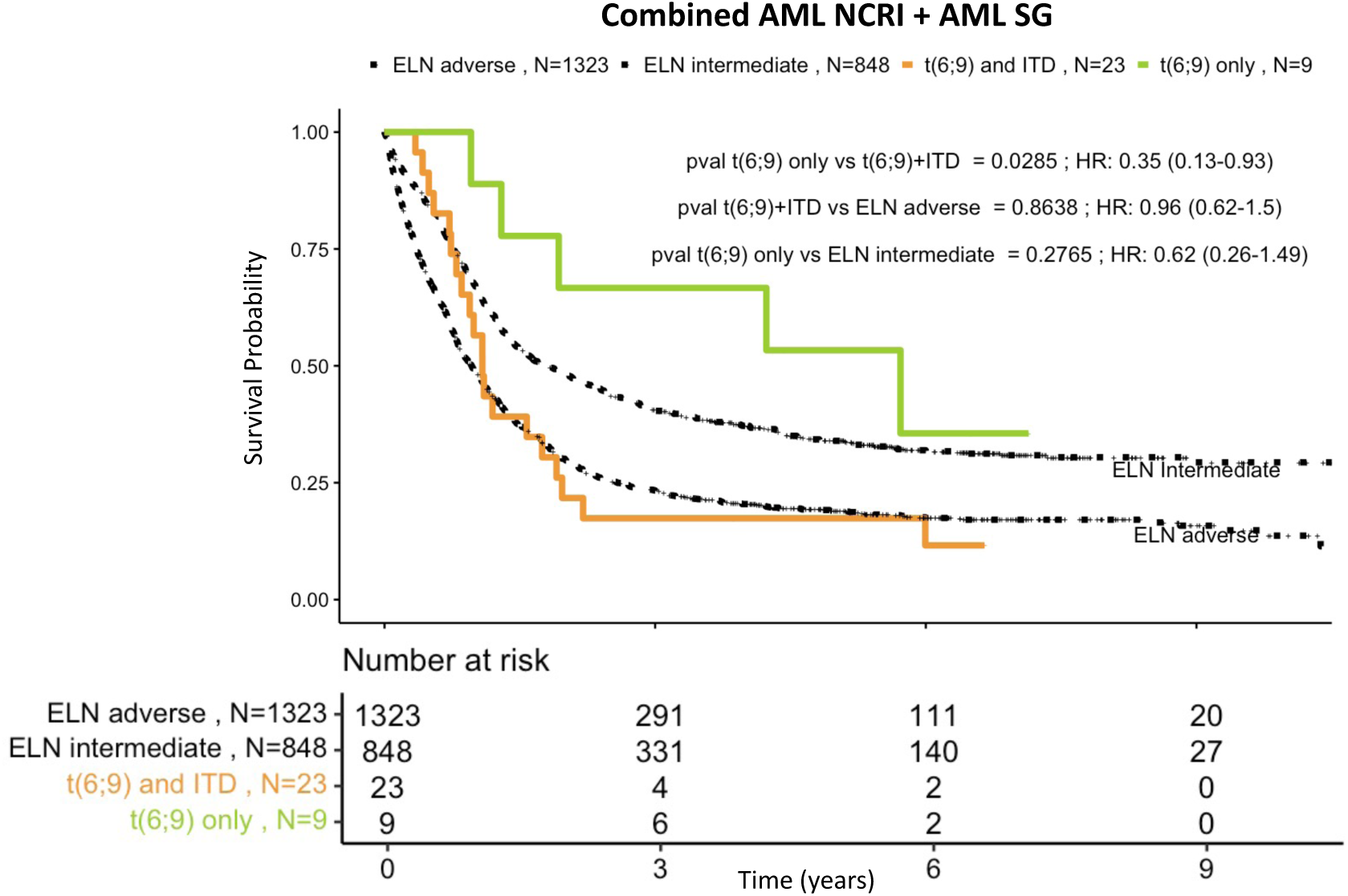
Kaplan-Meier curves for overall survival and associated risk table comparing. patients with t(6;9) with and without ITD with patients in intermediate and adverse ELN^2017^ on the combined (n=3,653) training AML NCRI (n=2,113) and validation AML SG cohort (n=1,540). Dotted curves represent the ELN^2017^ risk categories. Pvalues were computed using log-rank test.

**Extended Figure 9:**
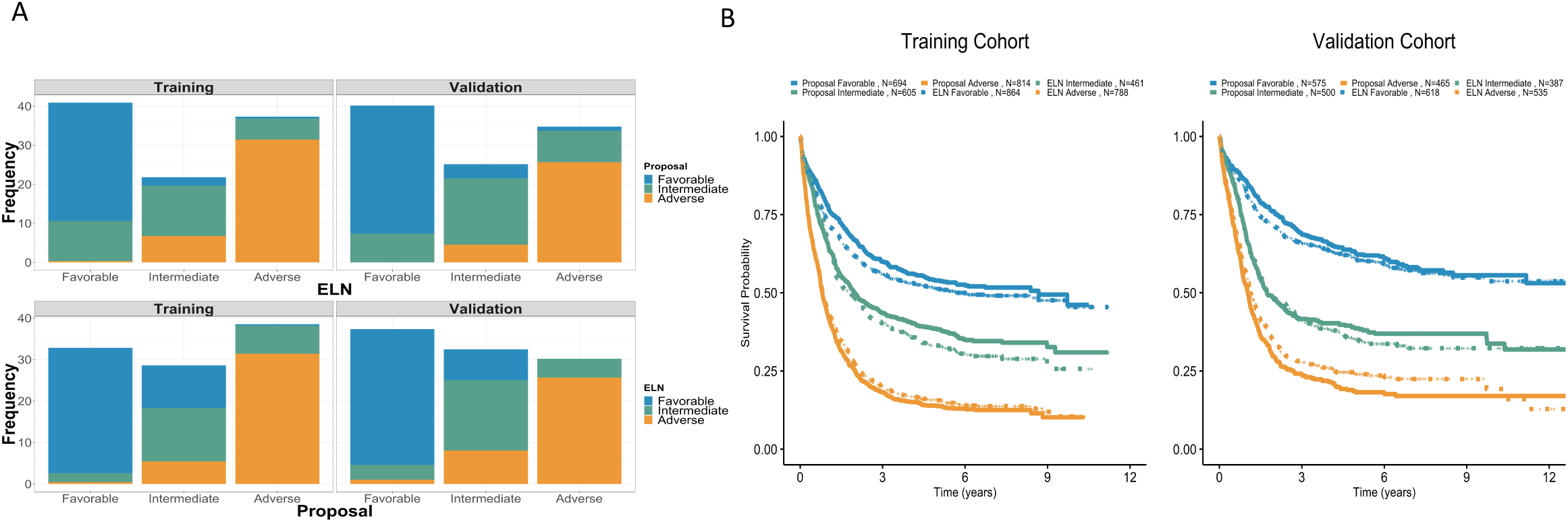
Comparison of ELN^2017^ and new risk proposal in the training AML NCRI (n=2,113) and validation AML SG Cohort (n=1,540). A. Top panel : Bar plots of frequencies of ELN^2017^ risk categories stratified by proposal categories in the training (n=2,113) and validation cohort (n=1,540). Bottom panel : Bar plots of frequencies of proposal risk categories stratified by ELN^2017^ categories in the training (n=2,113) and validation cohort (n=1,540). B. Kaplan-Meier curves for overall survival comparing the ELN^2017^ risk categories and the new risk proposal categories in the training (n=2,113) and validation cohort (n=1,540). Dashed curves are the three ELN^2017^ risk categories and plain curves are the three risk proposal categories.

**Extended Figure 10:**
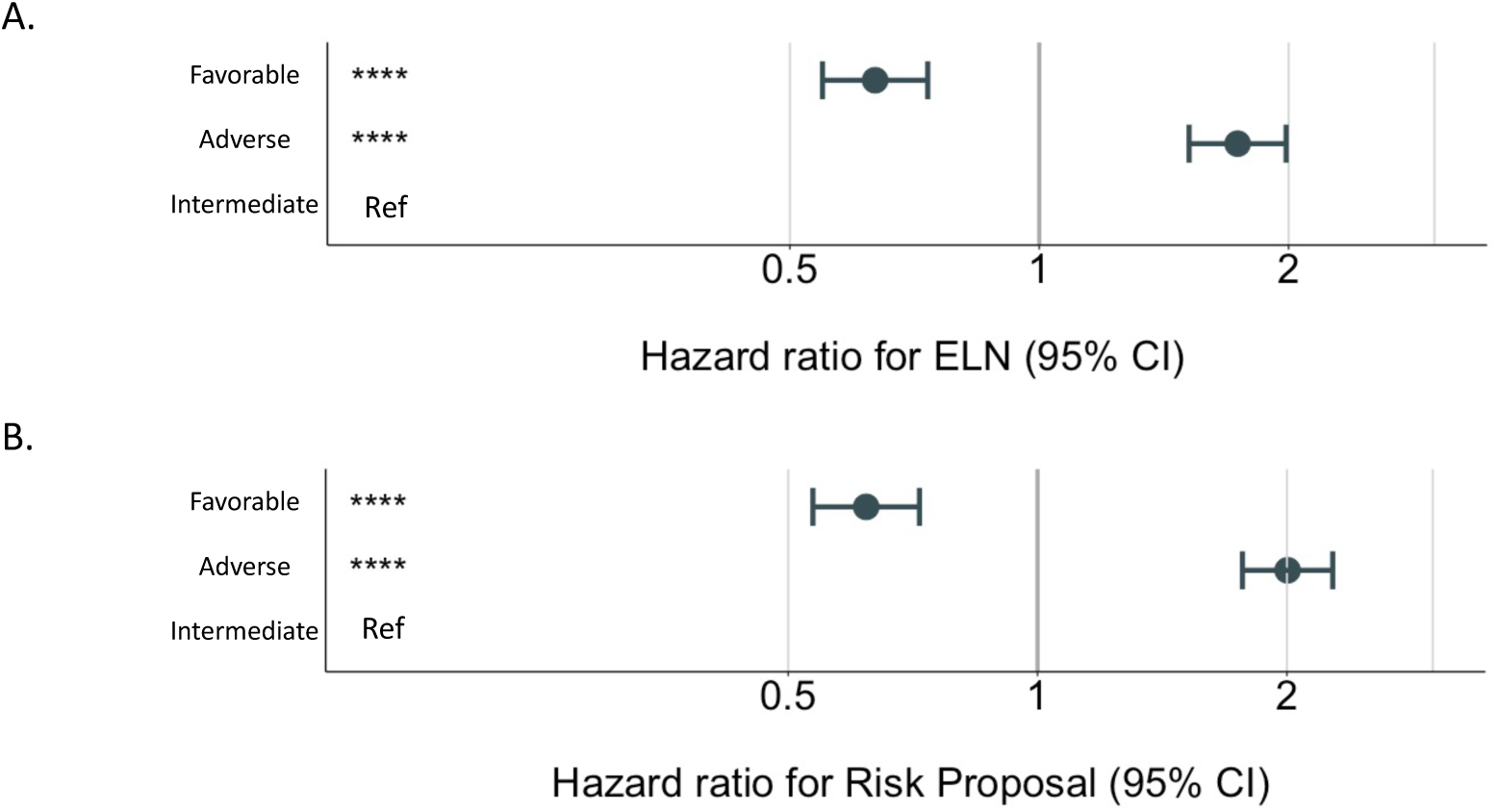
Forest plot multivariate Cox regression of. A. ELN^2017^ risk categories and B. risk proposal in NCRI trial study set (n= 2,113).

