## Supplementary material for "Unified classification and risk-stratification in Acute Myeloid Leukemia": Online methods

‡Shared second authorship

*Shared senior authorship

**Affiliations:**

1. Computational Oncology Service, Department of Epidemiology & Biostatistics, Memorial Sloan Kettering Cancer Center, New York, NY, USA.

2. Center for Hematologic Malignancies, Memorial Sloan Kettering Cancer Center, New York, NY, USA.

3. Tri-Institutional Computational Biology and Medicine PhD Program, Weill Cornell Medicine of Cornell University and Rockefeller University, New York, NY, USA.

4. The Rockefeller University, New York, NY, USA.

5. Centre for Trials Research, School of Medicine, Cardiff University, UK

6. Department of Haematology, School of Medicine, Cardiff University, UK

7. Institute of Immunology and Immunotherapy, University of Birmingham, UK

8. Nuffield Department of Population Health, University of Oxford, Oxford, UK

9. Department of Medical and Molecular Genetics, King's College, London, UK

10. Cancer, Ageing and Somatic Mutation Programme, Wellcome Sanger Institute, Hinxton, UK

11. Department of Hematology, Hemostasis, Oncology, and Stem Cell Transplantation, Hannover Medical School, Hannover, Germany

12. Department of Hematology, Oncology, and Tumorimmunology, Campus Virchow Klinikum, Berlin, Charité – Universitätsmedizin Berlin, corporate member of Freie Universität Berlin and Humboldt-Universität zu Berlin, Berlin, Germany

13. Department of Internal Medicine III, Ulm University, Ulm, Germany

14. Visiting Professor University of Glasgow, formerly Cardiff University, UK

15. The Christie NHS Foundation Trust, Manchester, U.K.

16. Department of Haematology, Nottingham University Hospital, Nottingham, UK

17. Department of Haematology and Wellcome Trust-MRC Cambridge Stem Cell Institute, University of Cambridge, UK

### ONLINE METHODS

#### 1. Clinical annotation

Demographic and diagnostic variables were ascertained at the time of diagnosis by the NCRI Clinical Trial Center. The variables collected included: Age of diagnosis (AOD), Gender, Bone marrow karyotype, AML type (AML: de novo AML, sAML: secondary AML, tAML: therapy related AML), Antecedent hematological disease (AHD), Performance status and peripheral blood counts to include white blood cell (WBC) counts, lactate dehydrogenase levels (LDH), platelet counts, hemoglobin levels (Hb). Outcome endpoints included status at last follow up, time from diagnosis to time of last follow up, event free survival and time to event, relapse free survival and time to relapse as well as whether complete remission was achieved, time to remission and time to transplant where transplant was received.

For patients in the NCRI AML17 trial flow cytometric MRD data were previously collected as described here[1].

#### 2. Molecular annotation

##### 2.1 Preparation of custom capture libraries for sequencing

Genomic DNA was extracted from peripheral blood or bone marrow mononuclear cells for adult AML patients ascertained through the UK NCRI AML Trials AML11, AML12, AML14, AML15, AML16, AML17 and AML Li1 (S.Table1). All samples were collected at diagnosis prior to any treatment. Custom RNA baits were designed per manufacturers' guidelines (SureSelect, Agilent, UK) complementary to all coding exons of 128 genes as well as genome wide SNP probes selected using the following criteria:1. Inter-marker distance every 3 MB; 2. Population Minor Allele Frequency >0.4; 3. Not within repeat regions. Detailed bait file design to include all regions targeted by RNA baits, alongside genomic coordinates and annotation are provided in Supplementary table 2.

A total of 125μl of 40ng/μl of WGA DNA was fragmented to an average insert size of 145bp (75I300) and subjected to Illumina DNA sequencing library preparation using the Bravo automated liquid handling platform. Individual samples were indexed using a unique DNA barcode via 6 cycles of PCR. Equimolar pools of 16 libraries were prepared and hybridized to custom RNA baits following the Agilent SureSelect protocol. Enriched pools of 96 cases were sequenced on a lane of an Illumina HiSeq Genome Analyzer machine using the 75 base pair paired-end protocol up to a median coverage of 600x. Only data that passed FASTQC MultiQC criteria (coverage >100x, duplicate reads, GC bias) were retained in the study.

All raw data has been deposited in the European Genome-Phenome Archive EGAS00001000570.

#### 2.2 Sequencing data alignment

Raw sequence data were aligned to the human genome (NCBI build 37) using BWA[2]. Unmapped reads, PCR duplicates and reads mapping to regions outside of the target region (merged exonic regions + 10bp either side of each exon) were excluded from analysis. Bedtools® coverage v2.15.0[5]was subsequently used to determine the coverage depth at each base. Genes with median target coverage < 20x were removed from the study and samples with median overall coverage < 50x were also excluded from downstream analysis and are not reported in this study.

#### 2.3 Variant calling

##### 2.3.1 Substitutions

Base substitutions were mapped using established bioinformatics approaches as previously described[3–5]. Single base, somatic substitutions were called independently in each sample using an in-house algorithm CaVEMan: Cancer Variants through Expectation Maximisation[6]. The algorithm compares sequence data from each tumor sample to an unrelated normal sample and calculates a mutation probability at each base-pair position locus. A number of post-processing filters were applied to improve specificity. Filters applied to targeted capture data required that:

1. At least a third of the alleles containing the mutant must have base quality >= 25.
2. If mutant allele coverage >= 10, there must be a mutant allele of at least base quality 20 in the middle 3rd of a read. If mutant allele coverage is < 10, a mutant allele of at least base quality 20 in the first 2/3 of a read is acceptable[3].
3. The mutation position is marked by <3 reads in any sample in the unmatched normal panel.
4. If the mean base quality is <20 then less than 96% of mutation-carrying reads are in one direction.
5. Variants were cross-referenced with a panel of unmatched normal samples (n= 300) to allow definition of base pair specific errors in the panel/
6. Previously reported bona fide somatic variants presenting in the unmatched normal panel were not filtered out from the dataset.

##### 2.3.2 Small insertion and deletions

Small somatic insertions and deletions (indels) were identified using an in-house modified version of Pindel[7]. Post-processing filters were applied as previously described[8,9]. The following steps were taken to improve specificity for calling non-coding indels:

1. 'SUM-MS' score (sum of the mapping scores of the reads used as anchors) >=200
2. 'Previously Rejected Score' (PRS) is ==0
3. Bidirectional (evidence in both read directions (forward and reverse) in Pindel or BWA reads)
4. Variant allele is not a unit within a homopolymer track presenting with variant allele fraction < 8%.
5. Variants did not present in approximately 300 unmatched normal samples and did not have a COSMIC ID with confirmed somatic status in the literature.
6. Artifactual indels occur at recurrent loci across multiple samples, often as a consequence of highly repetitive sequence. To ensure that such variants were not retained in the data, in-house databases for recurrently rejected Pindel calls interrogation were performed. All variants were visually inspected prior to removal.

Regions enriched for GC content and low target coverage were manually reviewed (i.e. *CEBPA, SRSF2)* and, where available, prior data derived by a CLIA approved diagnostic laboratory, were cross-referenced and for *FLT3$^{ITD}$*, *CEBPA* mutations and *NPM1* mutations incorporated in the dataset.

Additionally, for *FLT3*$^{ITD}$ detection a custom analysis script that performs a localized query for reads consistent with an inverted tandem duplication within the *FLT3* locus was developed.

Visual inspection using visualisation software (IGV) was performed of all variants in the targeted gene screen dataset after applying these filters.

#### 2.4. Quality control and variant annotation

From the list of high confident somatic variants, putative oncogenic variants were distinguished from variants of unknown significance (VUS) based on:

- Recurrence in the Catalogue Of Somatic Mutations in Cancer (COSMIC)[9], in myeloid disease samples registered in cBioPortal[10,11] or in the study dataset.

- The inferred consequence of a mutation; where nonsense mutations, splice site mutations and frameshift indels were considered oncogenic for likely tumor suppressor genes (from COSMIC Cancer Census Genes or OncoKB Cancer Gene List).
- Presence in pan-cancer hotspot databases[12,13].
- Annotation in the human variation database ClinVar[14].
- Annotation in the precision oncology knowledge database OncoKB[4].
- Recurrence with somatic presentation in a set of in-house data derived from >6,000 myeloid neoplasms[3,5,9].

Briefly using these database variant annotation parameters are listed by variant type as follows:

**Oncogenic**

- Known oncogenic variants previously reported in the literature or databases;
- Novel recurrent variants ($n \geq 2$) that cluster with known somatic variants in well characterised myeloid driver genes;
- Truncating variants (nonsense mutations, essential splice mutations or frameshift indels) in genes implicated in myeloid malignancies through acquisition of loss of function mutations;

Details of all mutations annotated as oncogenic and retained for bioinformatic analyses are provided in S.Table 3.

**Variants of unknown significance**

Variants identified outside the range of frequent driver variants in genes with known oncogenic variants; Variants in genes whose role in myeloid disease is not yet established or variants presenting uniquely in the dataset were not considered in the study.

#### 2.5 Cytogenetic findings and copy number alterations (CNA)

Karyotypes for 983/2113, 46.5% were ascertained in accordance with the International System for Human Cytogenetic Nomenclature from diagnostic assessment. Recurrent alterations including fusion genes and copy number alterations were included in downstream analysis. Additionally we used Allele Specific Copy Number Analyses of Tumors (ASCAT[15]) to derive aberrant copy number segments at an arm or chromosome level resolution. Findings from CNA and cytogenetic findings from karyotype analyses were evaluated for concordance, as well as matched for patient gender, as a means to quality control that the derivative sample does indeed match the corresponding clinical data. Detail annotation of the recurrent copy number alterations and cytogenetic findings incorporated in the analyses is provided in S.Table 4.

#### 2.6 Annotation of TP53 allelic state

*TP53* allelic state[16] was annotated as follows:

- **Mono-allelic annotation** for samples with only one putative somatic mutation (substitution, small insertion or deletion, splice site mutation).
- **Multi-hit annotation** for samples that satisfied the following criteria:
    a. At least 2 mutations in TP53 (substitution, small insertion or deletion, splice site mutation).
    b. At least one mutation of TP53 and a concomitant deletion of the TP53 locus to include focal deletions, 17p deletions or whole chromosome 17 deletions.
    c. At least one mutation in TP53 at a VAF estimate > 65%, indicative of LOH.

### 3. Statistical methodology

#### 3. 1 Bayesian Dirichlet Process for derivation of class membership

We performed an *ab initio* evaluation of molecular classification in AML. We used a Bayesian Dirichlet process (BDP)(https://github.com/nicolaroberts/hdp), which defines a potentially infinite prior distribution for the number and proportions of clusters in a mixture model as previously described[3]. Briefly, using this process a dataframe of 2150 rows (for each patient) with 153 columns representing recurrent genetic alterations as binary variables (1 for present, 0 for wild type) is used as input (S.Table 4). The optimal number of clusters is learned from the data by Markov chain Monte Carlo (MCMC) methods. This approach does not constrain genetic features within one cluster but rather allows genetic lesions to be shared across clusters. This means that the resulting clusters can share genetic features. As the clusters are defined, patient samples obtain a probability of class membership for each cluster.

A 2-step Bayesian Dirichlet Process with Gaussian distribution was implemented. In the first iteration a total of x high confidence clusters were defined assigning x patients with a high probability of assignment >x%. Once components are determined, for each patient, a probability of assignment to each of the components is derived on the basis of the representative features for each class. Hyperparameter selection was dependent upon: 1. The total number of high-confidence components; 2. Maximum probability of assignment for each patient; 3. The delta between the maximum probability of assignment and the second highest probability of assignment. A second iteration followed, with input the subset of patients that were not classified in the first iteration, which led to the x patients that did not classify within the first iteration.

Selection of the execution parameters (i.e. hyperparameter) selection was performed using a gridsearch (S.Table 14).

- Cosine similarity threshold applied on the measure of distance between clusters to differentiate distinct components.
- Initial number of clusters: random initialisation to this predefined number of clusters before applying the Gibbs sampling procedure.
- Base distribution: The distribution from which the parent node will draw from representing the shape of the data.
- Concentration Parameter $\alpha_A$: shape hyperparameter for the gamma prior over the concentration parameter.
- Concentration Parameter $\alpha_B$: rate hyperparameter for the gamma prior over the concentration parameter.

**Post processing**

High confidence components were defined as non-overlapping clusters with uniform molecular composition that resulted in the highest probability of assignment of the constituent patients. Whilst each patient derives distinct probabilities of assignment for each cluster (i.e. Let Patient 1 with 60% assignment probability for cluster 1, 24% probability for cluster 2 and 16% probability for cluster 3), Patient 1 would be assigned to cluster 1. Patients were only assigned in a given component if the main class defining genes or cytogenetic alterations were present (S.Figure 9). For example, for a patient to be part of the NPM1 cluster, the patient has to have an *NPM1* mutation. Certain components, owing to similarity in patterns of commutation were assigned within the same cluster. These included t(15;17) and t(11;x), WT1 and t(6;9). A manual split into independent components was applied. For 91 patients in the study (4.3%) assignment criteria to more than 1 class post processing were fulfilled.

##### 3.2 Hierarchical class assignment

An overall hierarchy for AML classification was informed by the presence of unique and non-overlapping molecular subgroups, class defining alteration frequency, class size. For overlap cases, size, clinical presentation and severity of clinical phenotype and outcomes was prioritized in the hierarchy (i.e. established WHO entities preceded novel entities, *TP53* and complex karyotype preceded the hierarchy a patient with *WT1* mutations).

Main Figure 1E outlines the hierarchy of assignment in the proposed classification in an R snippet code

#### 3.3 Survival analysis

Overall survival (OS) defined as the time to death or last follow-up was the primary endpoint of the study. Survival analyses was used by implementation of established statistical methodologies for modelling outcomes to include:

##### 3.3.1.Cox Penalized Models

Cox regression model[17,18] with regularization using a Lasso penalty[19]. The degree of shrinkage $\lambda$ was selected internally by cross validation (S. Table 15). The Lasso penalty has the effect of forcing some of the coefficient estimates, with a minor contribution to the model, to be exactly equal to zero. Cox Random Effects Models (https://github.com/mg14/CoxHD) were applied to introduce a Ridge type regularisation shrinking the effect of correlated variables (S. Table 15).

##### 3.3.2 Cox Boosting $L^2$ Models

Cox likelihood-based boosting[20] approach with the negative $L^2$-norm penalised partial likelihood were performed (S. Table 15).

##### 3.3.3Random Survival Forests

Random Survival Forest[21] Models with the logrank splitting criteria were used (S. Table 15).

##### 3.3.4 Support Vector Machine Survival (SVM)

SVM[22] with truncated Newton optimization and order statistic trees that did not rely on the number of comparable pairs of events was applied (S. Table 15).

#### 3.4 Evaluation Metric using the concordance index(C-Index):

The discriminaton of various models were compared using the concordance index (C-Index)[23]. The C-Index applied for all comparisons used an inverse probability of censoring weight (IPCW) to account for censored observations.

Models that consider different feature groups (genes mutations, cytogenetic findings, clinical variables, demographic variables, molecular classes and/or ELN risk strata) were evaluated across algorithmic approaches using the following workflow: 75% of patients in the dataset were randomly selected for the training and cross-validation. Model performances were evaluated on the 25% of remaining patients to compute the C-Index (evaluation metric).95% confidence intervals for the C-Index estimates were constructed using bootstrap resampling (100 bootstrap resamples). P-values were calculated using the C-Index estimates along with the standard error estimates from the bootstrap resampling. The same process was conducted for training and validation cohorts respectively (2113 patients for AML NCRI cohort and 1540 for AML SG cohort).

A number of algorithmic approaches were evaluated to include following hyperparameters selection:

- Cox Model with penalisation:
    - different values of $\alpha$: (0 to 1 with 0.2 increment) controlling the tradeoff Ridge-Lasso
    - internal validation of $\lambda$ parameter controlling the weight given to the coefficients penalisation
- Cox Boost with the following hyperparameters:
    - maxstepno=500: maximal number of steps to evaluate
    - K=10: number of folds to be used for cross-validation to find the optimal number of boosting steps
    - type="verweij": used to compute the partial likelihood in the hold-out folds with Verweij method
    - penalty=100: penalty value for the update of an individual element of the parameter vector in each boosting step.
- Cox Random Effects with the following hyperparameters:
    - Groups: as many groups as number of variables and with
    - $\nu$ = 0 meaning no hyperprior

- $\sigma_0$: the variance of a si-chisq hyperprior on the variance
- max.iter=500: maximal number of iterations
- tolerance=0.01: the stopping criteria

- Random Forest Survival:
  - Nodesize: 5,10,20
  - Number of trees: 100 to 1200 with 50 increments
  - Split rule: log-rank splitting
- Support Vector Machine Survival:
  - Kernels: linear, polynomial, radial basis function and sigmoid
  - $\alpha$: range 1e-6 to 1 multiplying by 10 to penalize the square hinge loss in the objective function. Internal cross validation was used to select this parameter.
  - Avltree optimizer named after inventors Adelson-Velsky and Landis
  - Max iter = 1000: maximal number of iterations
  - Tolerance = 1e-6

Details about softwares and packages used are available in S. Table 15.

Algorithmic comparison on the best performing feature combination was also investigated in S.Figure 43.

#### 3.5 Feature importance

A permutation strategy was developed to evaluate feature importance. The reference C-Index (evaluation metric) was computed and the values of each feature were permuted 50 times to compare the new C-Index with the reference C-Index without permutation. With this framework, each feature is ranked from the most to the least important for model performance using the values of the ratio = ref_CI / permuted_CI. This strategy is useful to characterize the relative association of each covariate with the given endpoint (i.e. survival) without information on direction of effect. This strategy was also compared with 1. the variable importance returned by random forest and 2. the coefficients given by the Cox penalised models and results were ranked similarly. This permutation strategy was applied for different feature combinations on the following models: Cox Lasso, Cox Ridge, Cox ElasticNet, Cox Random Effects and Random Forest Survival with the same parameters as presented in the above section.

#### 3.6 Multi-state models to model transitions across clinical endpoints

Multi-state models[24,25] were developed to describe a stochastic process where patients experience a succession of events (alive (received induction chemotherapy) , complete remission, relapse and death) and transition between different states overtime. Markov assumption is implicitly present in the likelihood of state transition: the future depends on history only through the present.

First, a non-parametric approach was performed to estimate overall transition probabilities within the cohort. Second, a semi-parametric approach was used by incorporating covariates of interest into the modeling of transition probabilities with different baseline transitions. The implementation to model those transitions was performed using the *mstate* package in R.

#### 3.7 Additional Statistical tests

Fisher's exact test[26] was used to compare patterns of co-mutations for each genetic abnormality. Due to multiple comparisons, all P-values were adjusted using the false discovery rate (Benjamin-Hochberg procedure). To compare continuous correlates (age,wbc,hb,plt and blasts) by mutational or class status, a Wilcoxon rank-sum test[27] was used. To compare other categorial correlates (gender,ahd,perf_status,secondary,eln) by mutational or class status, a chi-square or Fisher's exact test were conducted, as appropriate. A test of interaction within a Cox proportional hazards model was used to evaluate whether a giving class modifies the effect of MRD or transplant on survival or relapse. Lastly, survival was compared across two or more groups using a logrank test[28].

#### 3.8 Regression analysis for clinical variables

Univariate regression volcano plots**:** Linear model was fitted using the *lm* package in R for continuous outcomes and a logistic regression (logistf package in R) for binary outcomes. In the volcano plot, each covariate coefficient on the x axis and the log (base 10) false discovery rate adjusted p-value value on y axis were plotted.

Multivariate regression coefficient rankings**:** Linear and logistic Lasso models were fitted using the *cv.glmnet* R package with internal cross validation of the regularization parameter $\lambda$. Each model used the entire dataset 100 times and the coefficients were aggregated for each of the covariates to evaluate absolute ranking.

#### 3.9 Hazard risk over time and risk density

The *bshazard* package in R was applied to obtain a non-parametric smooth estimate of the hazard function based on B-splines[29].

In Figure 2 of the manuscript, the hazard density across the ELN$^{2017}$ risk categories and the classes were plotted to visualize a shift in the hazard across the classes. The x-axis represents the hazard and the y-axis was rescaled to aid for the visualization of the hazards.

#### 4. Estimation of contributing factors in the multistate model

We used the Cox semi-parametric approach to estimate the different transition probabilities where we specify covariates to be incorporated in the model. The transition hazard takes the following form:

- $h_{ij}(t|Z) = h_{ij,0}(t) \exp(\beta^{T}_{ij} Z_{ij})$ , where $\beta^{(k)}_{ij}$ is the coefficient computed from the Cox transition model corresponding to covariate (k) for the transition from state i to state j , $Z_{ij}^{(k)}$ is the value for covariate (k) for that transition.

Thus, we compute the contributing factors for each transition :

$Factor^{CONTRIB} = \beta_{ij}{}^{*}(Z_{ij}-Z_{ij,median})$ values, with $Z_{ij,median}$ the median value for that covariate in this transition.

A negative $Factor^{CONTRIB}$ value corresponds to a low risk covariate value transition while a positive value corresponds to a high risk covariate value transition.

We omitted n=96 patients from the multi-state model for disease progression (2,113-96=2,017) due to missing timepoints.

### 5. Development of study web portal

#### 5.1 Data accessibility and code reproducibility

All raw data have been deposited on EGAS00001000570. Additionally. a web portal to accompany this publication has been deployed on: https://www.aml-risk-model.com The portal includes 1. A direct link to cbio portal containing detailed molecular and clinical findings in the study; 2. List of gene panel used to develop the AML classification and risk stratification model; 3. A gene by gene description of all genotype-clinical, genotype-genotype, and genotype-outcome associations in the study; 4. A class by class description of all class-clinical, class-genotype and class-outcome associations; and 5. A personalized risk calculator tool. Additionally a GitHub https://github.com/yanistazi/AML_Repo has been deployed that contains all input data, code and data visualizations relating to this publication.

#### 5.2 AML Risk calculator

For the development of a personalized risk calculator we consider 3,201 intensively treated patients from the AML NCRI and AMLSG cohorts. For each patient,cytogenetic findings and mutations in the panel of 32 genes were considered to derive class assignment, ELN[2017] and proposed risk scores.

Cox multi-state models[22,23] were used to estimate the transition covariate coefficients across 6 clinical endpoints (Alive (received induction chemotherapy) → Alive in CR; Alive → Death no CR; Alive in CR → Alive in Relapse; Alive in CR → Death in CR; Alive in Relapse → Death in Relapse) for the following 4 models: 1. AML Classes; 2. ELN[2017]; 3. Proposed risk score; 4. AML Classes, demographic and clinical parameters. For detailed information on the coefficients please refer to: https://www.aml-risk-model.com/calculator

For any patient outside of the cohort, given any combination of molecular, clinical and demographic parameters the web calculator determines a patient's class assignment, ELN[2017] and proposed risk score and estimates the transition probabilities for each of the 6 clinical endpoints. All of the 32 genes and cytogenetic findings are assumed to be wild type unless specified by the user. Clinical and demographic parameters that were not specified were imputed as the median for the cohort.

To increase confidence in the model interpretation, we have added the following features in the calculator :

1. Display of the overall cohort distribution for all clinical variables. This allows a practicing physician to appreciate whether the presentation of a given patient is within the expected or outlier ranges of the cohort.
2. Where input parameters for a patient represent outlier values, we have introduced a warning sign alerting the end user that the data supporting predictions for this patient may be less powered.
3. For the sediment plots, displaying probabilities of transitioning between each clinical point, we have added confidence intervals for the predictions. Confidence intervals are also available for the probability estimates.

#### 5.3 Implementation in AWS

A web application was developed, where the risk model is deployed as a serverless lambda function available through a restful API, and consumed by a single page javascript application. The web calculator is built using Python and Javascript open-source libraries for web development, and the R multi-state model is uploaded and executed on AWS Lambda in real-time to adjust for personalized transition probabilities prediction relative to the user's input parameters. Details of the services infrastructure and the cloud implementation using Amazon Web Services are shown in S.Figure 44.

20. De Bin R. Boosting in Cox regression: a comparison between the likelihood-based and the model-based approaches with focus on the R-packages CoxBoost and mboost. Comput Stat

2016;31(2):513–31.
