## Supplementary Appendix : Figures and Tables for "Unified classification and risk-stratification in Acute Myeloid Leukemia"

†Shared second authorship

*Shared senior authorship

**Affiliations:**

1. Computational Oncology Service, Department of Epidemiology & Biostatistics, Memorial Sloan Kettering Cancer Center, New York, NY, USA.

2. Center for Hematologic Malignancies, Memorial Sloan Kettering Cancer Center, New York, NY, USA.

3. Tri-Institutional Computational Biology and Medicine PhD Program, Weill Cornell Medicine of Cornell University and Rockefeller University, New York, NY, USA.

4. The Rockefeller University, New York, NY, USA.

5. Centre for Trials Research, School of Medicine, Cardiff University, UK

6. Department of Haematology, School of Medicine, Cardiff University, UK

7. Institute of Immunology and Immunotherapy, University of Birmingham, UK

8. Nuffield Department of Population Health, University of Oxford, Oxford, UK

9. Department of Medical and Molecular Genetics, King's College, London, UK

10. Cancer, Ageing and Somatic Mutation Programme, Wellcome Sanger Institute, Hinxton, UK

11. Department of Hematology, Hemostasis, Oncology, and Stem Cell Transplantation, Hannover Medical School, Hannover, Germany

12. Department of Hematology, Oncology, and Tumorimmunology, Campus Virchow Klinikum, Berlin, Charité – Universitätsmedizin Berlin, corporate member of Freie Universität Berlin and Humboldt-Universität zu Berlin, Berlin, Germany

13. Department of Internal Medicine III, Ulm University, Ulm, Germany

14. Visiting Professor University of Glasgow, formerly Cardiff University, UK

15. The Christie NHS Foundation Trust, Manchester, U.K.

16. Department of Haematology, Nottingham University Hospital, Nottingham, UK

17. Department of Haematology and Wellcome Trust-MRC Cambridge Stem Cell Institute, University of Cambridge, UK

### Supplementary Figures

**S.Figure 1: Study Cohort.** A. Training cohort characteristics for UK AML NCRI Trial study set (n=2,113)**.** B. Validation cohort characteristics for AML SG study set (n=1,540).

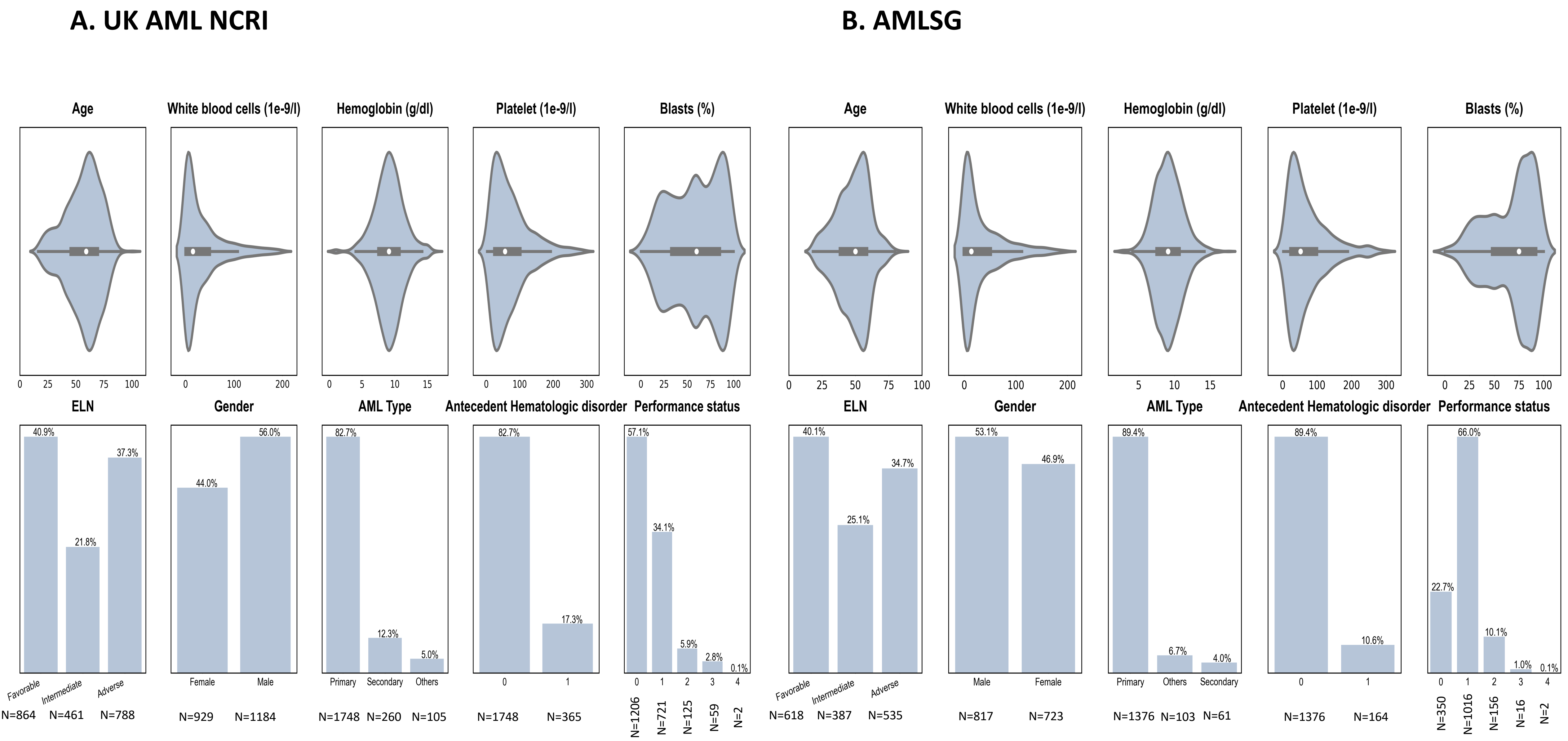

**S.Figure 2: Molecular characteristics.** Distribution of gene mutations and cytogenetic abnormalities in A. AML NCRI trial study set (2,113 patients and 8,460 driver events) and B. AML SG study set (1,540 patients and 5,043 driver events).

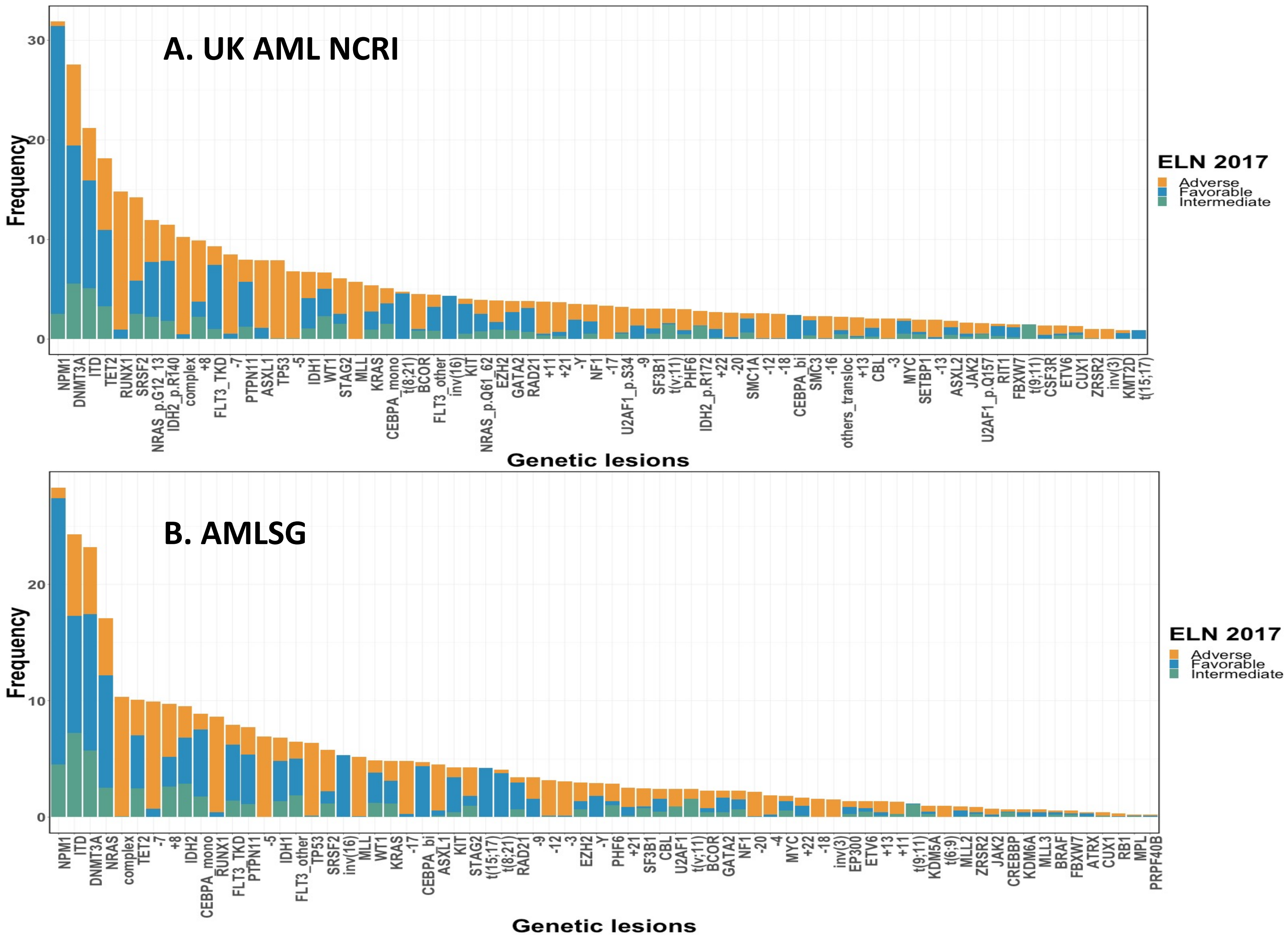

**S.Figure 3: Age and ELN2017 relationships to survival.** Kaplan-Meier curves for overall survival by age and ELN2017 stratification in A. NCRI trial study set (n=2.113) and B. AMLSG study set (n=1,540).

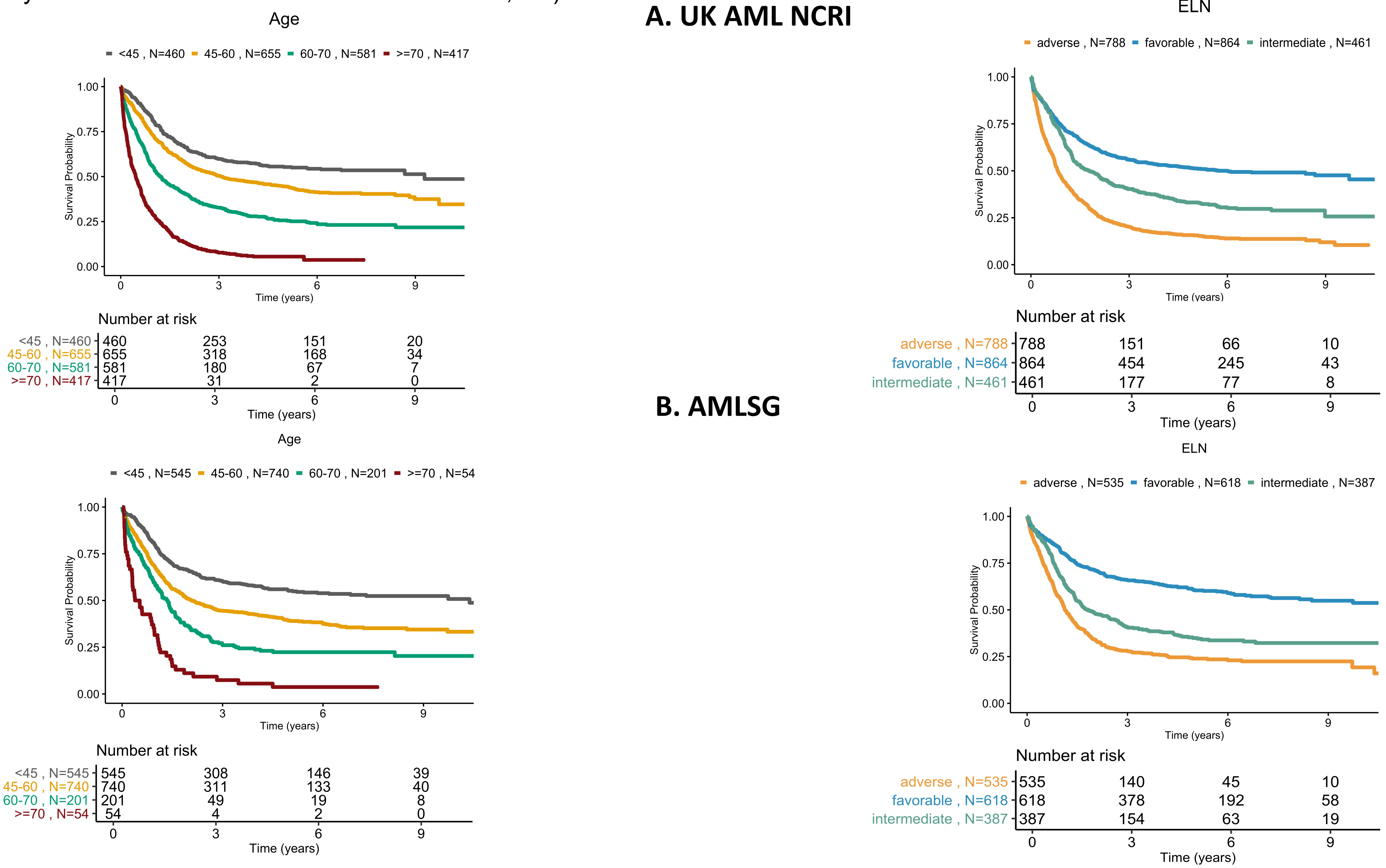

**S.Figure 4: Molecular predictors of clinical correlates.** A. Volcano plots for univariate regression analysis for each clinical parameter (Antecedent hematologic disorder, Age, Bone marrow blasts, Platelets, White blood cell counts and hemoglobin) in NCRI trial study set (n= 2,113). Genes are colour coded in blue, whilst cytogenetic events in grey. The size of each point corresponds to the frequency of the event. The horizontal dotted curve corresponds to the p-value threshold of 0.05 (on the y-axis) and the vertical one corresponds to β=0 (x axis). Significant predictors are highlighted (p-value less than 0.05).
B. Multivariate regression ranking of clinical correlates in NCRI trial study set (n= 2,113). The numbers correspond to the ranking of the absolute value of the β coefficients while the colour intensities represent the absolute coefficient values ranking (red with high intensity for largest negative value and blue with high intensity for largest positive value). The regression process is explained in S.Appendix. Abbreviations: ahd refers to antecedent hematologic disorder, plt to platelet, wbc to white blood cells and hb to hemoglobin.

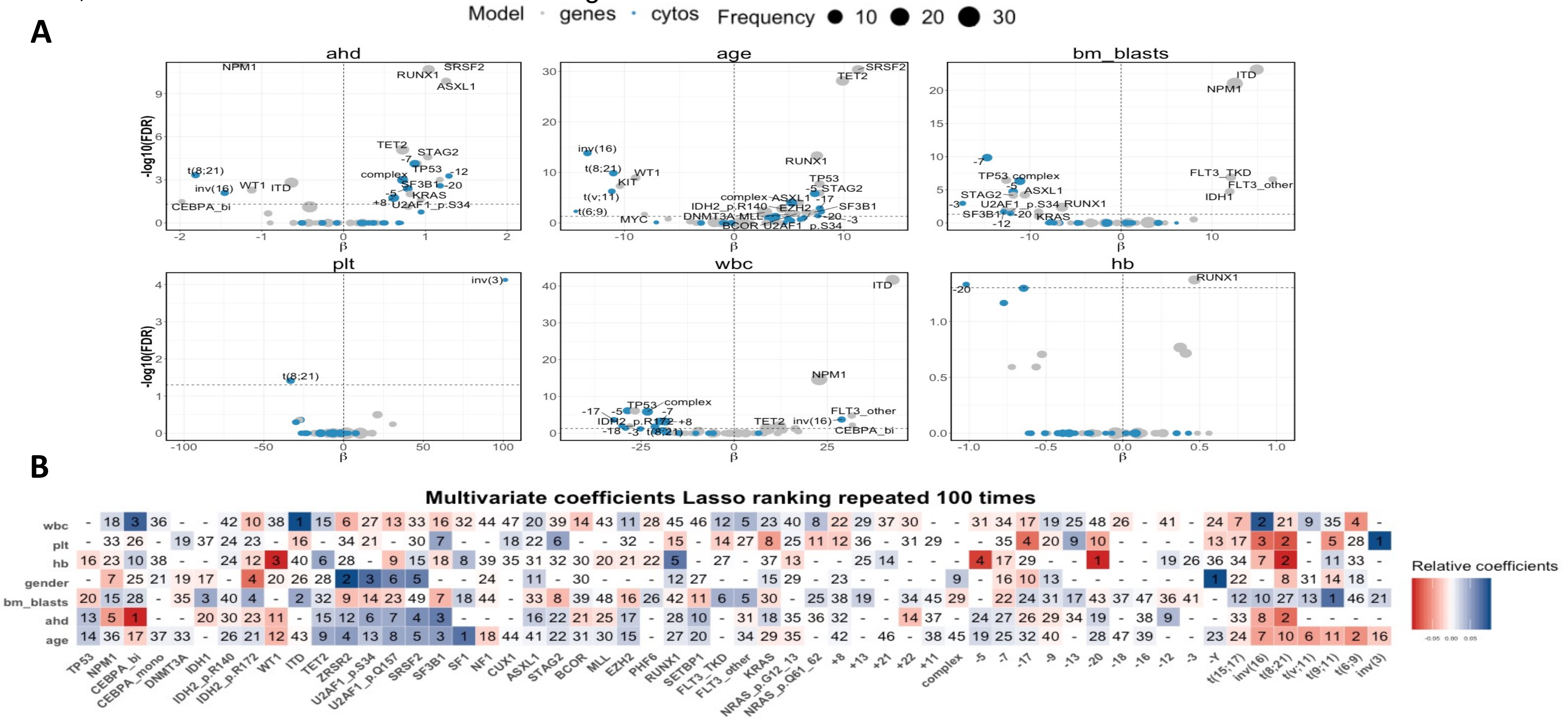

**S.Figure 5: Repartition of AML NCRI Cohort (n= 2,113) per WHO 2016 guidelines to include the provisional categories defined by *RUNX1* and t(9;22).**

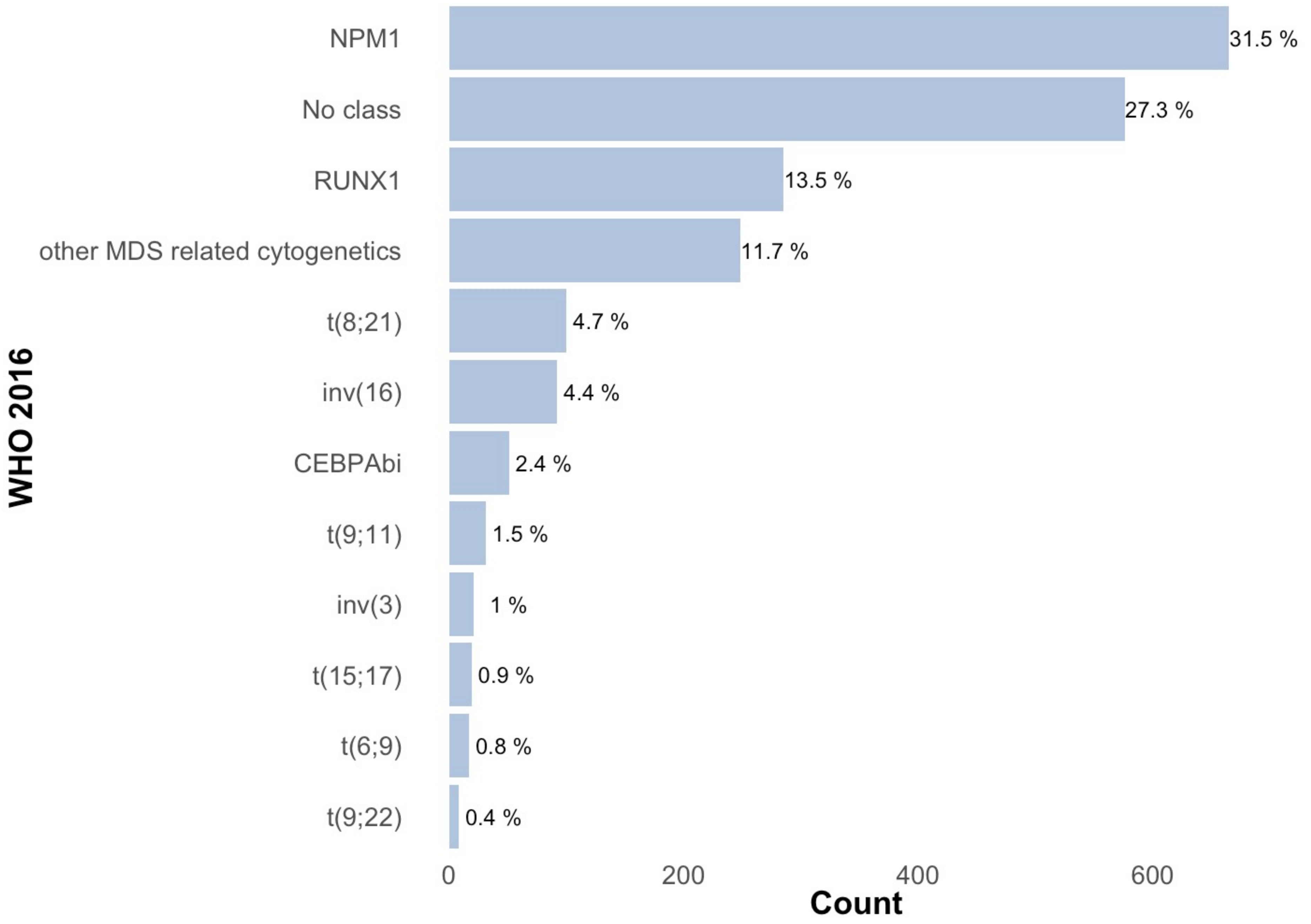

**S.Figure 6: Molecular classifications in AML.** A. Representation of the AML NCRI cohort (n=2,113 patients) using the WHO classification, the New England Journal of Medicine 2016 classification and the proposed molecular classification in this study. B. Sankey Plots comparing those different classifications. NOS means not otherwise specified.

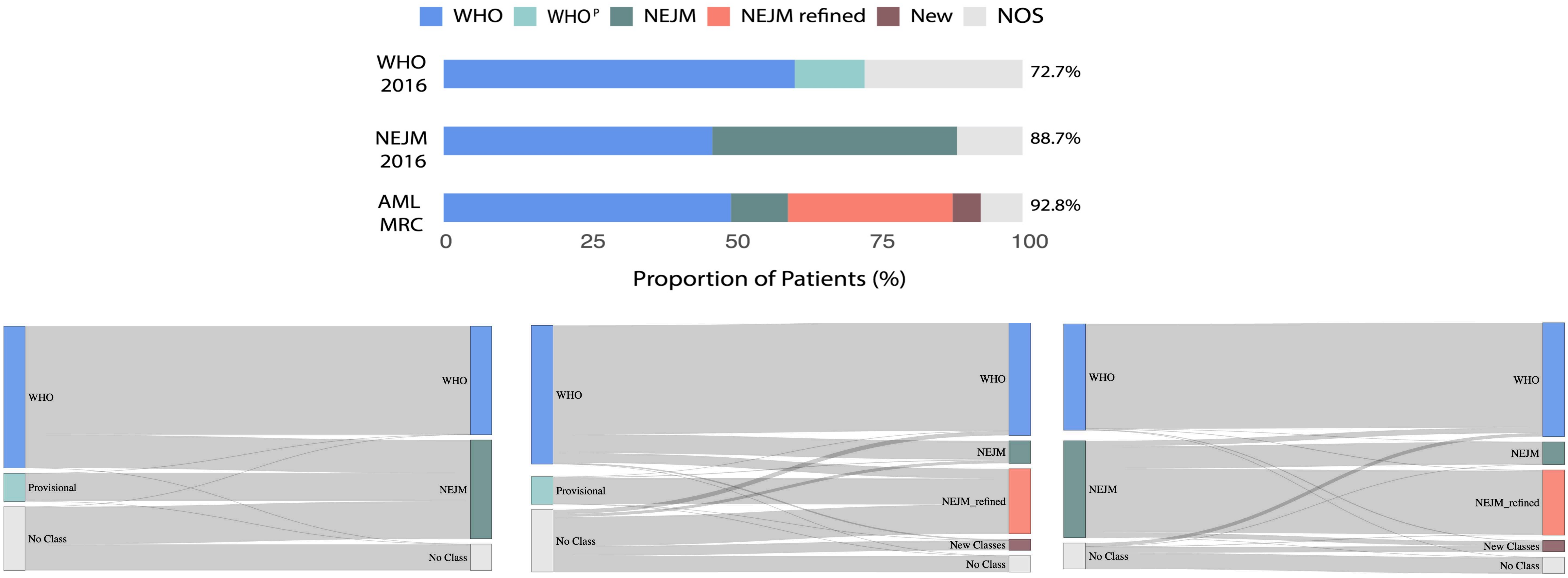

**S.Figure 7: Prognostic relevance of mutation number in sAML Like subgroups.** A. Kaplan-Meier Curves for overall survival comparing AML class with sAML1 (green plain line) and sAML2 (blue plain line) mutations to ELN[2017] risk groups in AML NCRI cohort (training, n=2,113) and AMLSG (validation cohort, n=1,540). B. Kaplan- Meier curves for overall survival comparing outcomes associated with specific gene mutations in the context of sAML1 and sAML2 subgroup in AML NCRI cohort (n=2,113). Log-rank tests compared the survival distributions between the 2 subgroups.

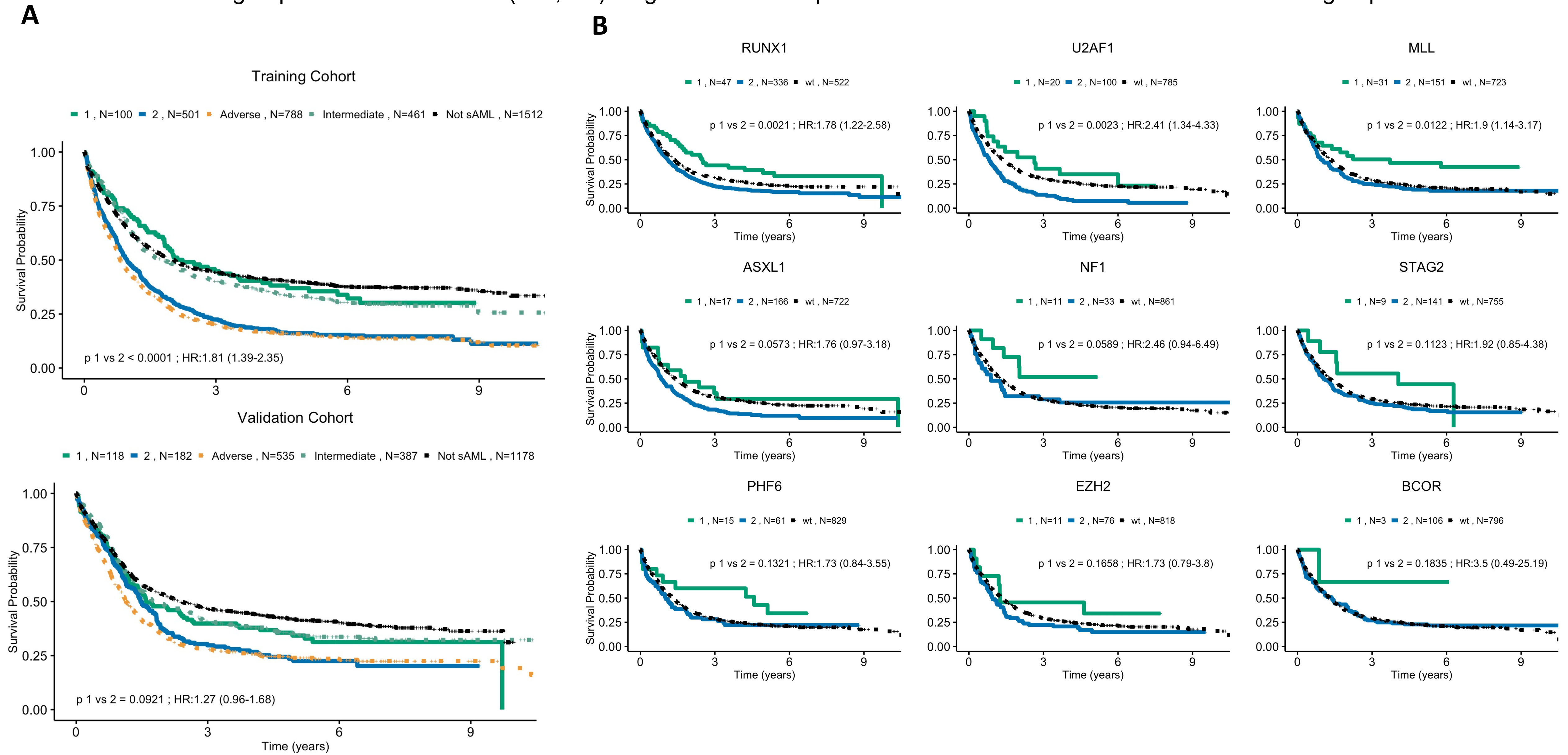

**S.Figure 8: Prognostic relevance of AHD in sAML2.** Kaplan-Meier curves for overall survival and associated risk table comparing outcomes in sAML2 patients (training, n=501) on the basis of AHD status. ahd=0 corresponds to patients without antecedent hematologic disorder while ahd=1 corresponds to patient with antecedent hematologic disorder. Log-rank tests compared the survival distributions between the 2 subgroups.

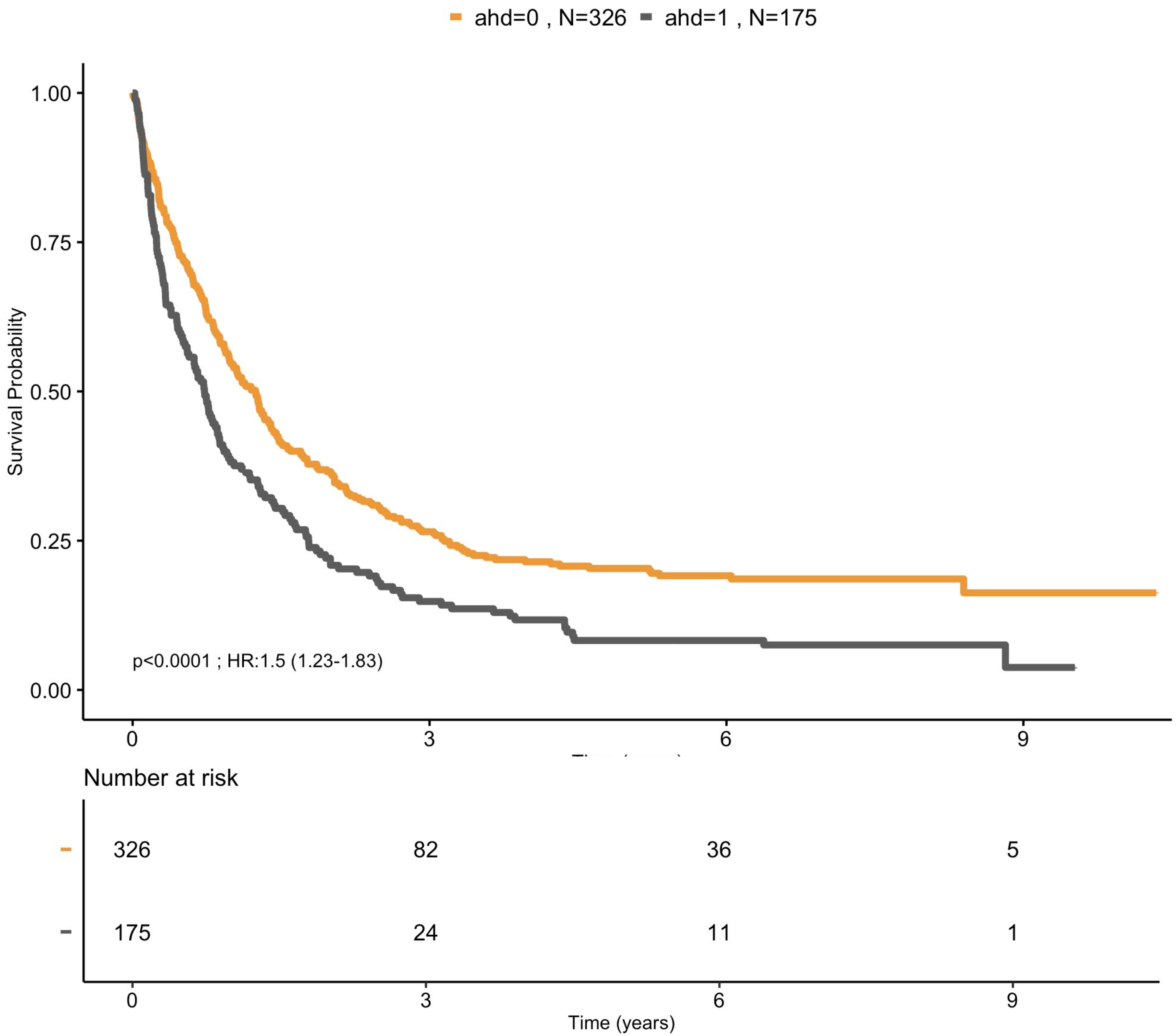

**S.Figure 9: Kaplan-Meier and associated risk table for RUNX1 mutation** for the 2 secondary AML like classes in the AML NCRI cohort (N=2,113). Log-rank tests compared the survival distributions between the 2 subgroups.

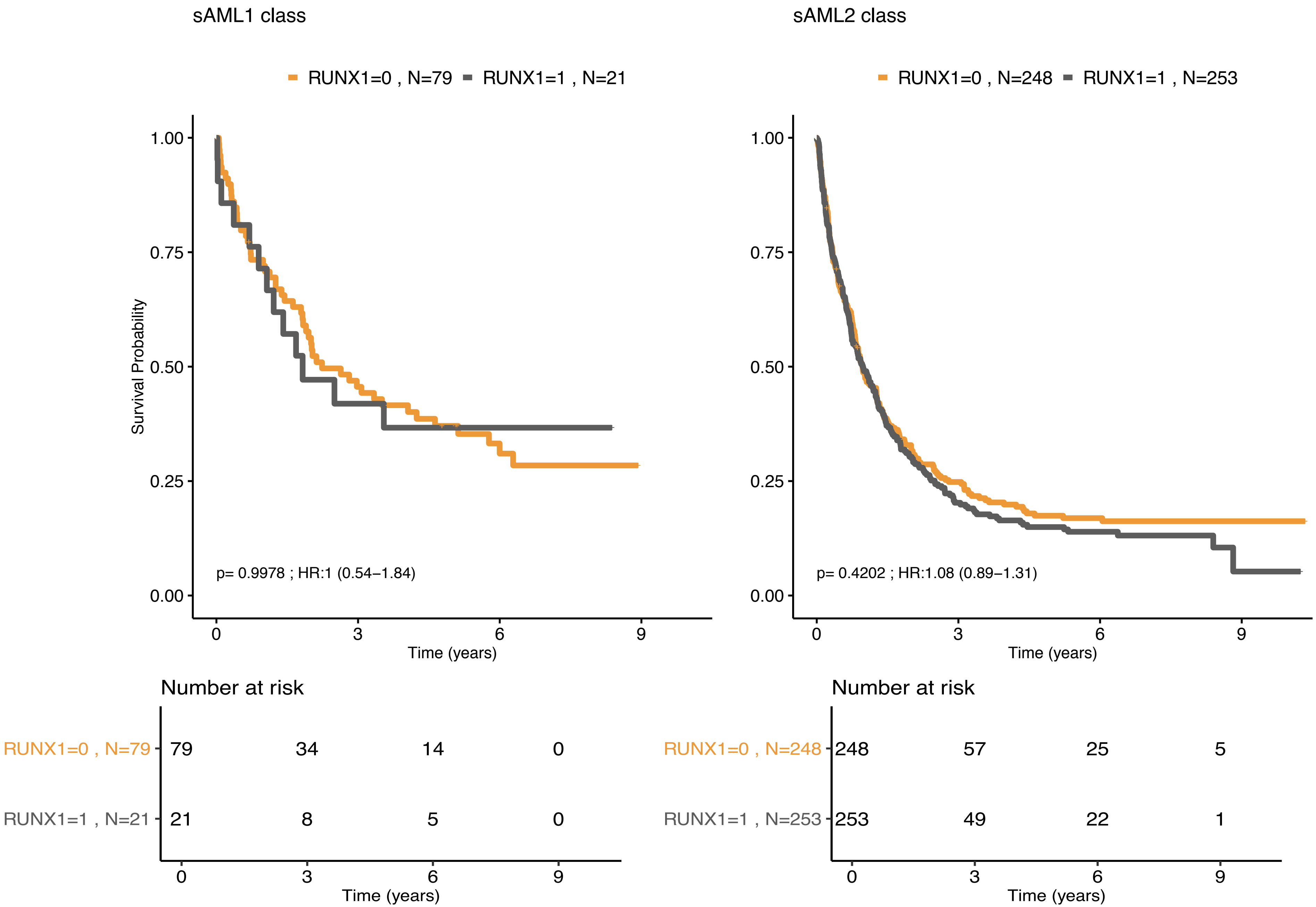

**S.Figure 10: WT1 Class.** A. Distribution of age for patients in WT1 class compared to other AML. Pvalue was computed with a Wilcoxon rank-sum test. B. Kaplan-Meier Curves for overall survival and associated risk table comparing WT1 class with $FLT3^{ITD}$ to $ELN^{2017}$ adverse risk group in AML NCRI cohort (n=2,113). Log-rank tests compared the survival distributions between the 2 subgroups. C. Kaplan-Meier Curves for overall survival and associated risk table for patients in WT1 class without $FLT3^{ITD}$ to $ELN^{2017}$ intermediate risk group in AML NCRI cohort (n=2,113). Log-rank tests compared the survival distributions between the 2 subgroups.

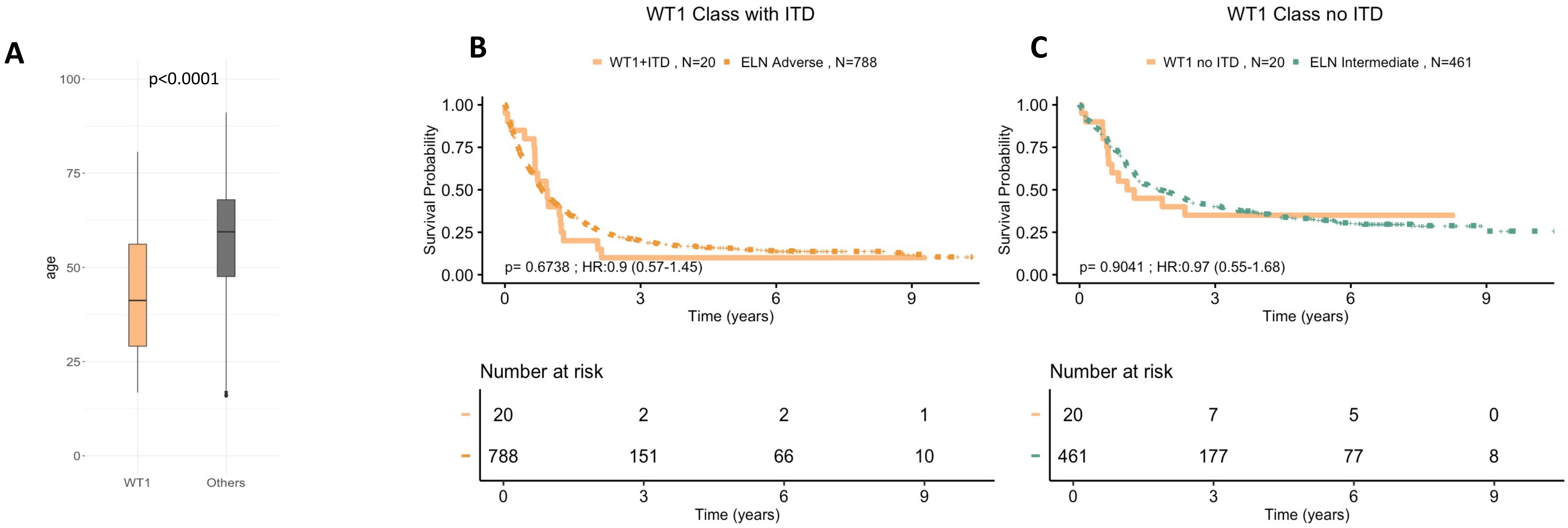

**S.Figure 11: DNMT3A-IDH class.** Kaplan-Meier curves for overall survival and associated risk tables comparing
A. AML class DNMT3A IDH to ELN$^{2017}$ intermediate risk groups and other AML patients in AML NCRI cohort (training, n=2,113).
B. AML subclass DNMT3A IDH2 $^{R140}$ to ELN$^{2017}$ intermediate risk groups and other AML patients in AML NCRI cohort (training, n=2,113).
C. AML subclass DNMT3A IDH2 $^{R172}$ to ELN$^{2017}$ intermediate risk groups and other AML patients in AML NCRI cohort (training, n=2,113).

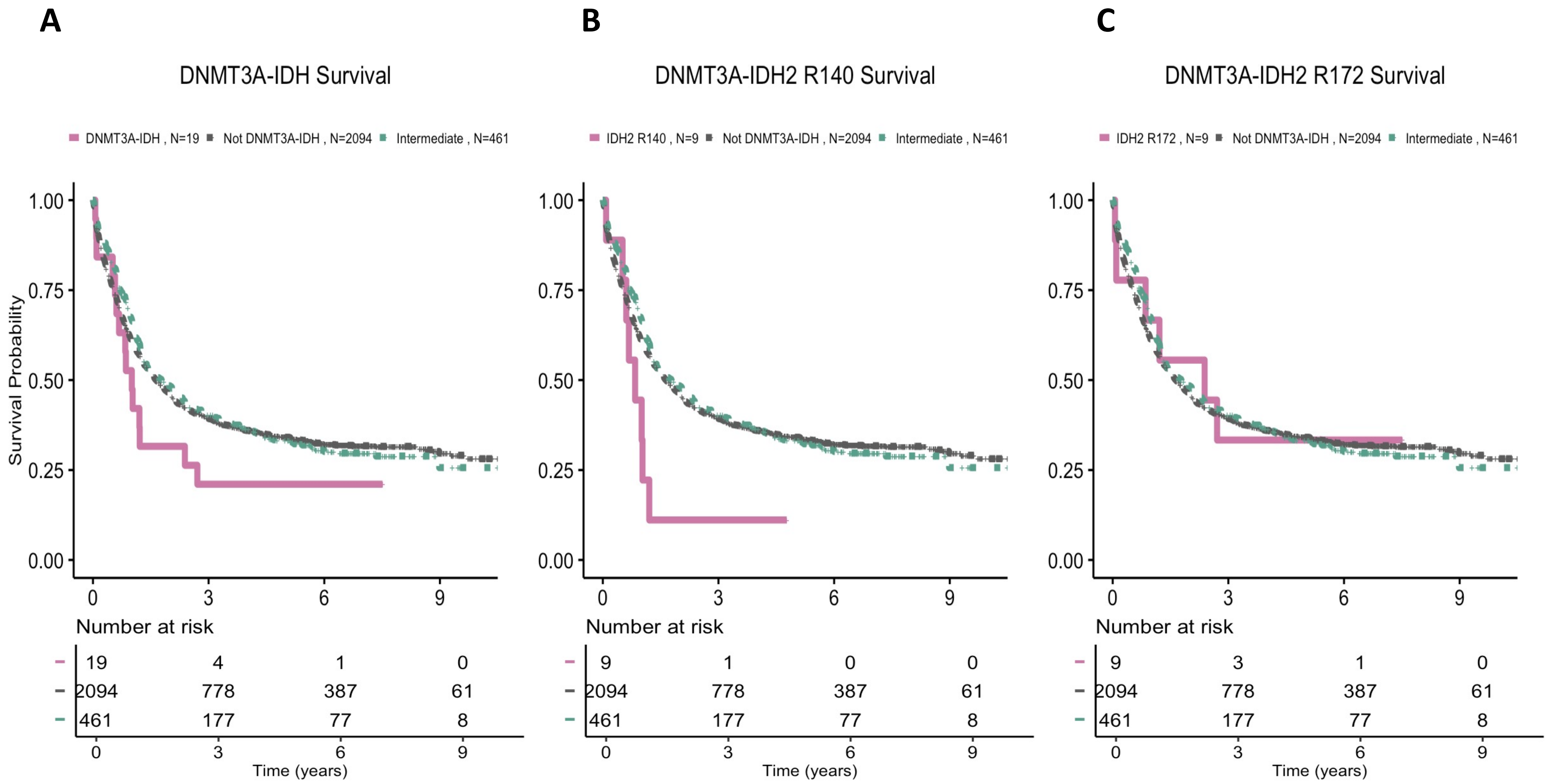

**S.Figure 12: Overall survival K-M curves by AML class.** A. Kaplan-Meier curves for overall survival comparing all AML classes and ELN$^{2017}$ risk strata in AML NCRI cohort (n=2,113). B. Risk stratification table by AML class and ELN$^{2017}$ score in AML NCRI cohort (n=2,113). C. 10 years 95% confidence interval survival estimates by AML class and ELN$^{2017}$ score in AML NCRI cohort (n=2,113).

**A.**

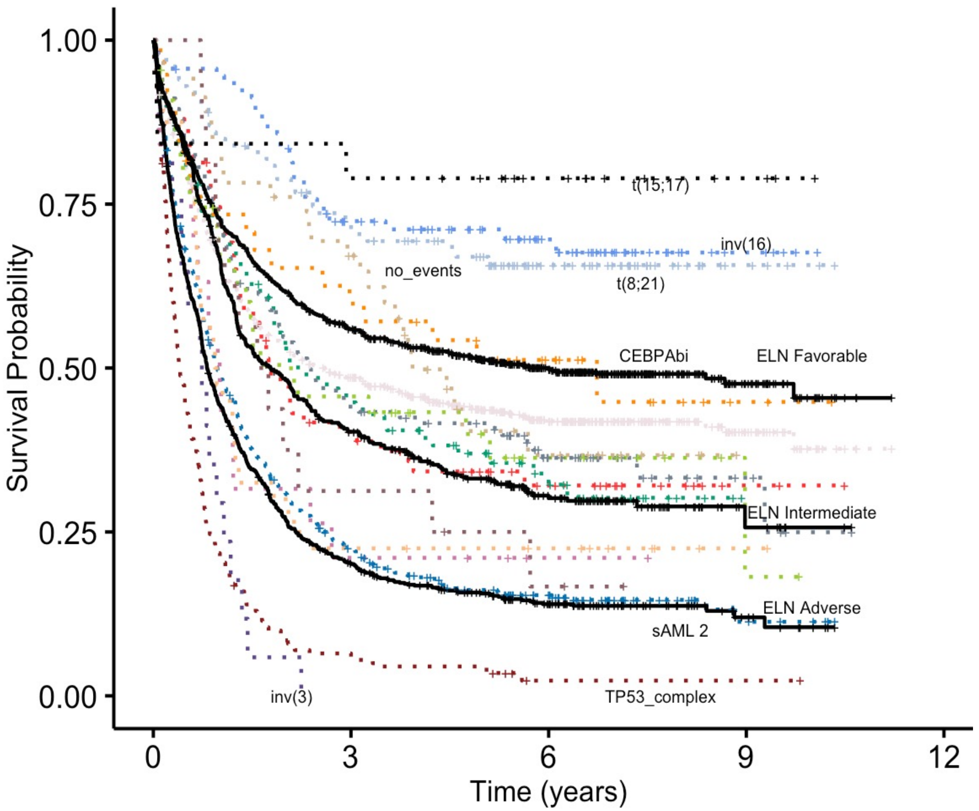

**B.**

Number at risk

| | 0 | 3 | 6 | 9 | 12 |
|---|---|---|---|---|---|
| NPM1 , N=674 | 674 | 309 | 162 | 30 | 0 |
| t(11) , N=75 | 75 | 28 | 14 | 2 | 0 |
| TP53_complex , N=208 | 208 | 13 | 1 | 1 | 0 |
| sAML2 , N=501 | 501 | 106 | 47 | 6 | 0 |
| sAML1 , N=100 | 100 | 42 | 19 | 0 | 0 |
| CEBPAbi , N=38 | 38 | 22 | 13 | 2 | 0 |
| DNMT3A_IDH1_2 , N=19 | 19 | 4 | 1 | 0 | 0 |
| inv(3) , N=17 | 17 | 0 | 0 | 0 | 0 |
| mNOS , N=124 | 124 | 54 | 27 | 5 | 0 |
| t(8;21) , N=100 | 100 | 67 | 34 | 5 | 0 |
| no_events , N=46 | 46 | 29 | 10 | 0 | 0 |
| WT1 , N=40 | 40 | 9 | 7 | 1 | 0 |
| inv(16) , N=92 | 92 | 60 | 35 | 5 | 0 |
| Trisomies , N=44 | 44 | 19 | 7 | 1 | 0 |
| t(6;9) , N=16 | 16 | 5 | 2 | 0 | 0 |
| t(15;17) , N=19 | 19 | 15 | 9 | 3 | 0 |
| ELN Favorable , N=864 | 864 | 454 | 245 | 43 | 0 |
| ELN Intermediate , N=461 | 461 | 177 | 77 | 8 | 0 |
| ELN Adverse , N=788 | 788 | 151 | 66 | 10 | 0 |

Time (years)

**C.**

| | |
|---|---|
| **NPM1** | 0.38 (0.32-0.45) |
| **t(11)** | 0.32 (0.23-0.45) |
| **TP53-complex** | 0.02 (0.01-0.07) |
| **sAML2** | 0.11 (0.07-0.17) |
| **sAML1** | 0.3 (0.22-0.42) |
| **CEBPAbi** | 0.45 (0.3-0.68) |
| **DNMT3A-IDH** | 0.21 (0.09-0.5) |
| **inv(3)** | 0 |
| **mNOS** | 0.25 (0.13-0.47) |
| **t(8;21)** | 0.66 (0.57-0.76) |
| **No events** | 0.37 (0.24-0.56) |
| **WT1** | 0.22 (0.13-0.4) |
| **inv(16)** | 0.68 (0.58-0.79) |
| **Trisomies** | 0.18 (0.04-0.77) |
| **t(6;9)** | 0.17 (0.05-0.54) |
| **t(15;17)** | 0.79 (0.63-1) |
| **ELN Favorable** | 0.45 (0.4-0.51) |
| **ELN Intermediate** | 0.26 (0.19-0.34) |
| **ELN Adverse** | 0.1 (0.07-0.15) |

**S.Figure 13: Kaplan-Meier and associated risk tables** for overall survival curves for the sAML 1, sAML 2, trisomies, WT1, no event and mNOS subgroups, separated by ELN$^{2017}$ scores.

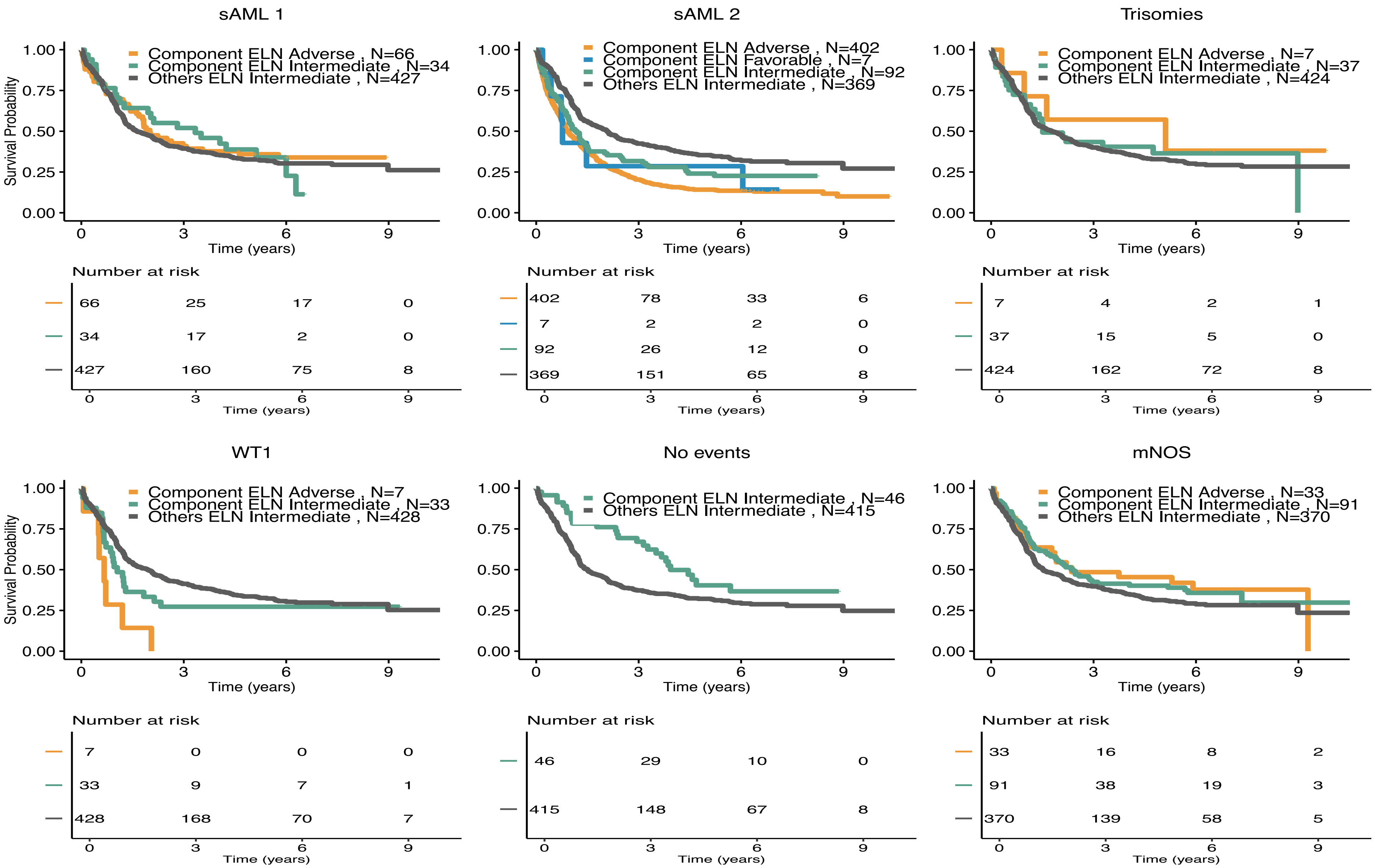

**S.Figure 14: Validation of class outcomes in the AML SG Cohort (n=1,540).** A. Kaplan-Meier curves for overall survival comparing all AML classes and ELN[2017] risk strata in AML SG cohort (n=1,540). B. risk stratification table by AML class and ELN[2017] score in AML SG cohort (n=1,540). C. 10 years 95% confidence interval survival estimates by AML class and ELN[2017] score in AML SG cohort (n=1,540).

**A.**

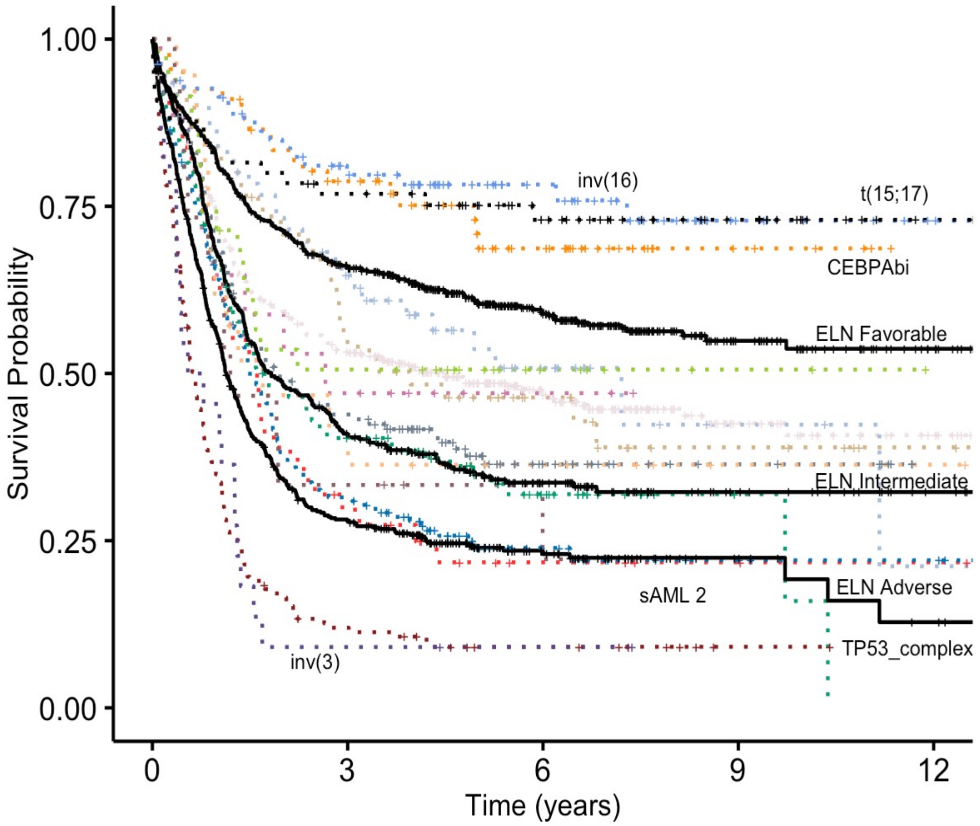

**B.**

Number at risk

| | 0 | 3 | 6 | 9 | 12 |
|---|---|---|---|---|---|
| NPM1 , N=436 | 436 | 221 | 102 | 29 | 12 |
| t(11) , N=47 | 47 | 13 | 4 | 2 | 1 |
| TP53_complex , N=165 | 165 | 18 | 8 | 1 | 0 |
| sAML2 , N=185 | 185 | 55 | 15 | 5 | 3 |
| sAML1 , N=119 | 119 | 45 | 17 | 2 | 0 |
| CEBPAbi , N=67 | 67 | 48 | 25 | 5 | 0 |
| DNMT3A_IDH1_2 , N=17 | 17 | 8 | 2 | 0 | 0 |
| inv(3) , N=22 | 22 | 2 | 1 | 0 | 0 |
| mNOS , N=148 | 148 | 61 | 20 | 7 | 0 |
| t(8;21) , N=61 | 61 | 36 | 13 | 3 | 1 |
| no_events , N=55 | 55 | 29 | 15 | 6 | 2 |
| WT1 , N=21 | 21 | 8 | 5 | 1 | 1 |
| inv(16) , N=82 | 82 | 58 | 33 | 8 | 2 |
| Trisomies , N=35 | 35 | 17 | 6 | 2 | 0 |
| t(6;9) , N=15 | 15 | 5 | 2 | 0 | 0 |
| t(15;17) , N=65 | 65 | 48 | 32 | 16 | 3 |
| ELN Favorable , N=618 | 618 | 378 | 192 | 58 | 18 |
| ELN Adverse , N=535 | 535 | 140 | 45 | 10 | 3 |
| ELN Intermediate , N=387 | 387 | 154 | 63 | 19 | 4 |

Time (years)

**C.**

| | |
|---|---|
| **NPM1** | 0.41 (0.35-0.48) |
| **t(11)** | 0.22 (0.12-0.38) |
| **TP53-complex** | 0.09 (0.06-0.15) |
| **sAML2** | 0.22 (0.16-0.3) |
| **sAML1** | 0.16 (0.04-0.66) |
| **CEBPAbi** | 0.69 (0.58-0.82) |
| **DNMT3A-IDH** | 0.47 (0.28-0.78) |
| **inv(3)** | 0.09 (0.02-0.34) |
| **mNOS** | 0.36 (0.29-0.46) |
| **t(8;21)** | 0.42 (0.27-0.66) |
| **No events** | 0.39 (0.27-0.57) |
| **WT1** | 0.36 (0.2-0.65) |
| **inv(16)** | 0.73 (0.62-0.85) |
| **Trisomies** | 0.51 (0.36-0.7) |
| **t(6;9)** | 0.22 (0.08-0.65) |
| **t(15;17)** | 0.73 (0.63-0.85) |
| **ELN Favorable** | 0.54 (0.49-0.59) |
| **ELN Adverse** | 0.19 (0.14-0.27) |
| **ELN Intermediate** | 0.32 (0.28-0.38) |

**S.Figure 15: Kaplan-Meier curves for overall survival and associated risk tables comparing**
A. AML class sAML2 to ELN$^{2017}$ adverse risk group in AML NCRI cohort on the subset of intensively treated patients (n=1,755). B. AML class TP53-complex to ELN$^{2017}$ adverse risk group in AML NCRI cohort on the subset of intensively treated patients (n=1,755). C. AML class sAML2 to ELN$^{2017}$ adverse risk group in AML NCRI cohort on the subset of non-intensively treated patients (n=358). D. AML class TP53-complex to ELN$^{2017}$ adverse risk group in AML NCRI cohort on the subset of non-intensively treated patients (n=358). Log-rank tests compared the survival distributions between the 2 subgroups.

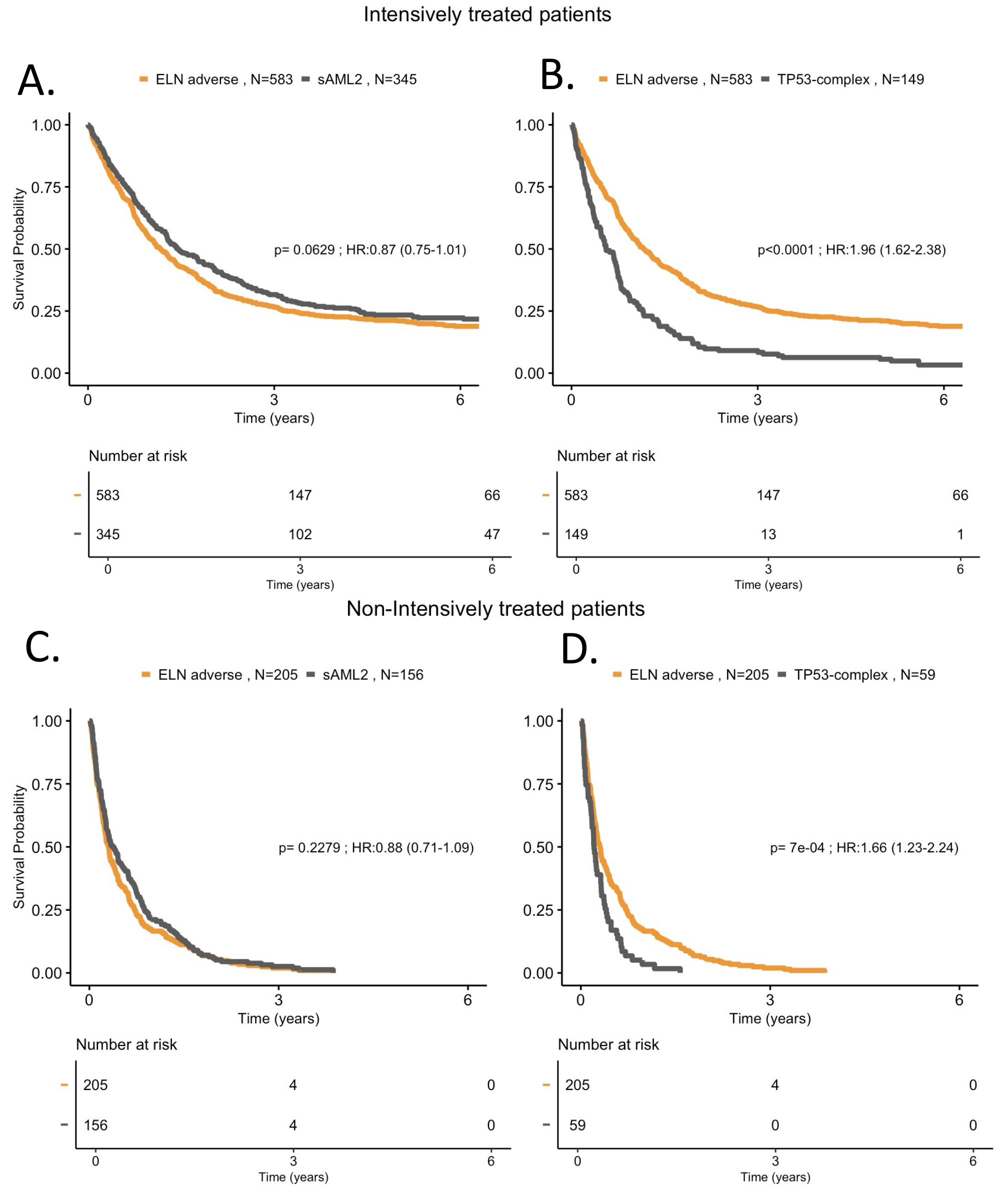

**S.Figure 16: Forest plot multivariate Cox regression of classes and monosomal karyotype** as defined by Breems et al. in NCRI trial study set (n= 2,113).

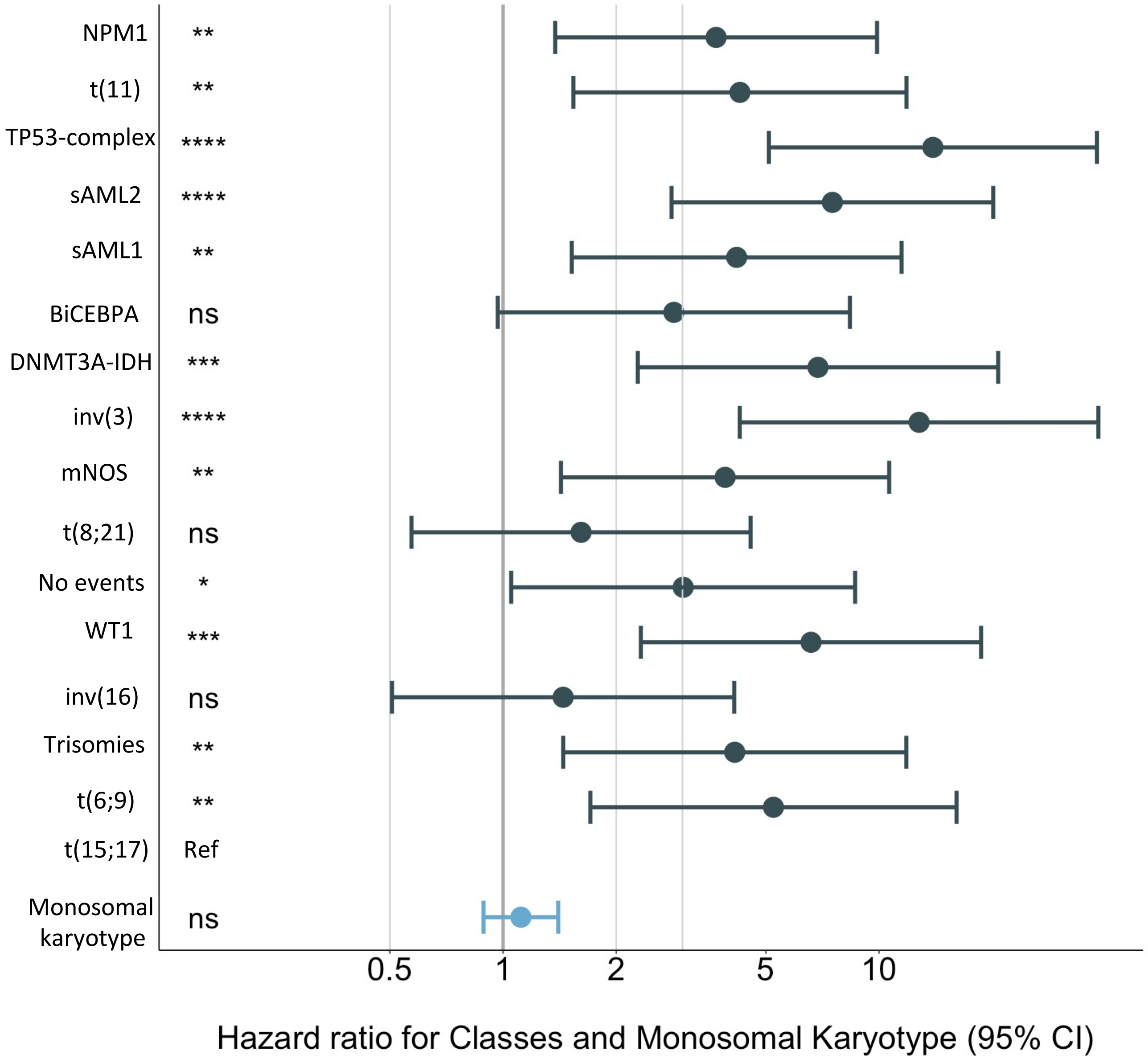

**S.Figure 17: Estimates of the concordance index (C-index) derived from Random Forest Survial model** that consider $ELN^{2017}$ strata; gene mutations; molecular classes; molecular classes + $FLT3^{ITD;}$ genetic data (gene mutations and cytogenetics); clinical and demographic; genetic, clinical and demographic ; classes, $FLT3^{ITD}$, clinical and demographic features in the AML NCRI trial (n=2,113).

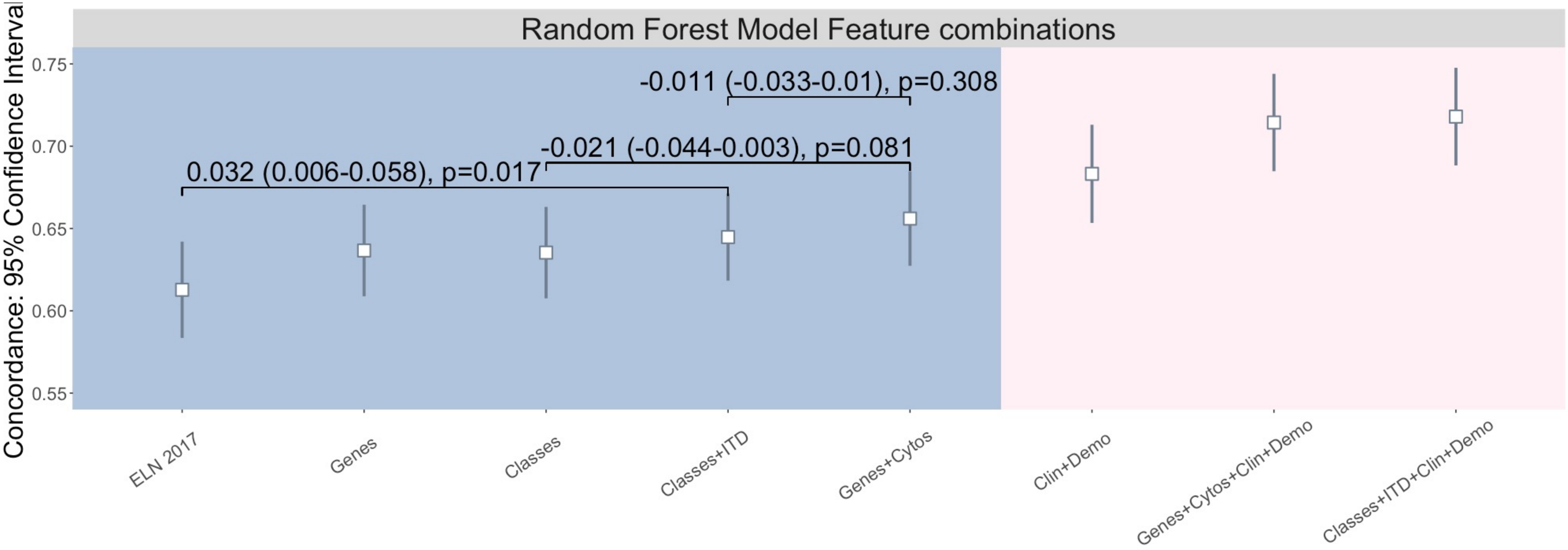

**S.Figure 18: Feature Importance in the class based model in AML NCRI Trial (n=2,113).**
The y axis corresponds to the different features evaluated by the models and ordered by importance. The x axis corresponds to the reshuffling ratio metric for each feature ( metric = reference C-Index / permuted C-Index ). The higher the ratio, the more sensitive the model is to that particular feature. The results are stratified by algorithms (Random Forest, Lasso, Ridge and Elastic-Net). For more details, please refer to S.Appendix. We omitted Random Effects algorithms for this class based model as the C-index ratio was constant across features.

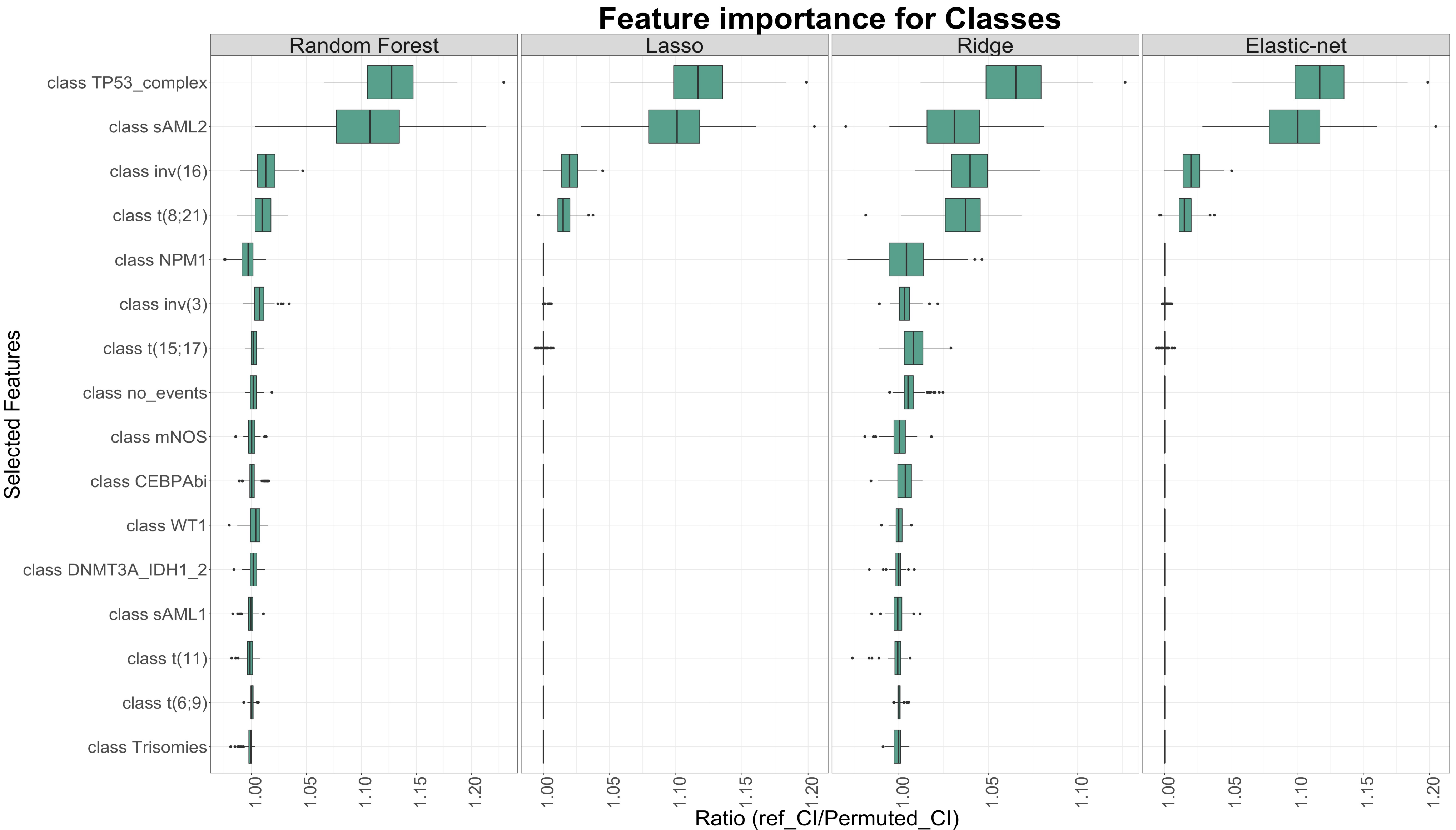

**S.Figure 19: Feature Importance in the genes based model (top 30) in AML NCRI Trial (n=2,113).**
The y axis corresponds to the different features evaluated by the models and ordered by importance. The x axis corresponds to the reshuffling ratio metric for each feature ( metric = reference C-Index / permuted C-Index ). The higher the ratio, the more sensitive the model is to that particular feature. The results are stratified by algorithms (Random Forest, Random Effects, Lasso, Ridge and Elastic-Net). For more details, please refer to S.Appendix.

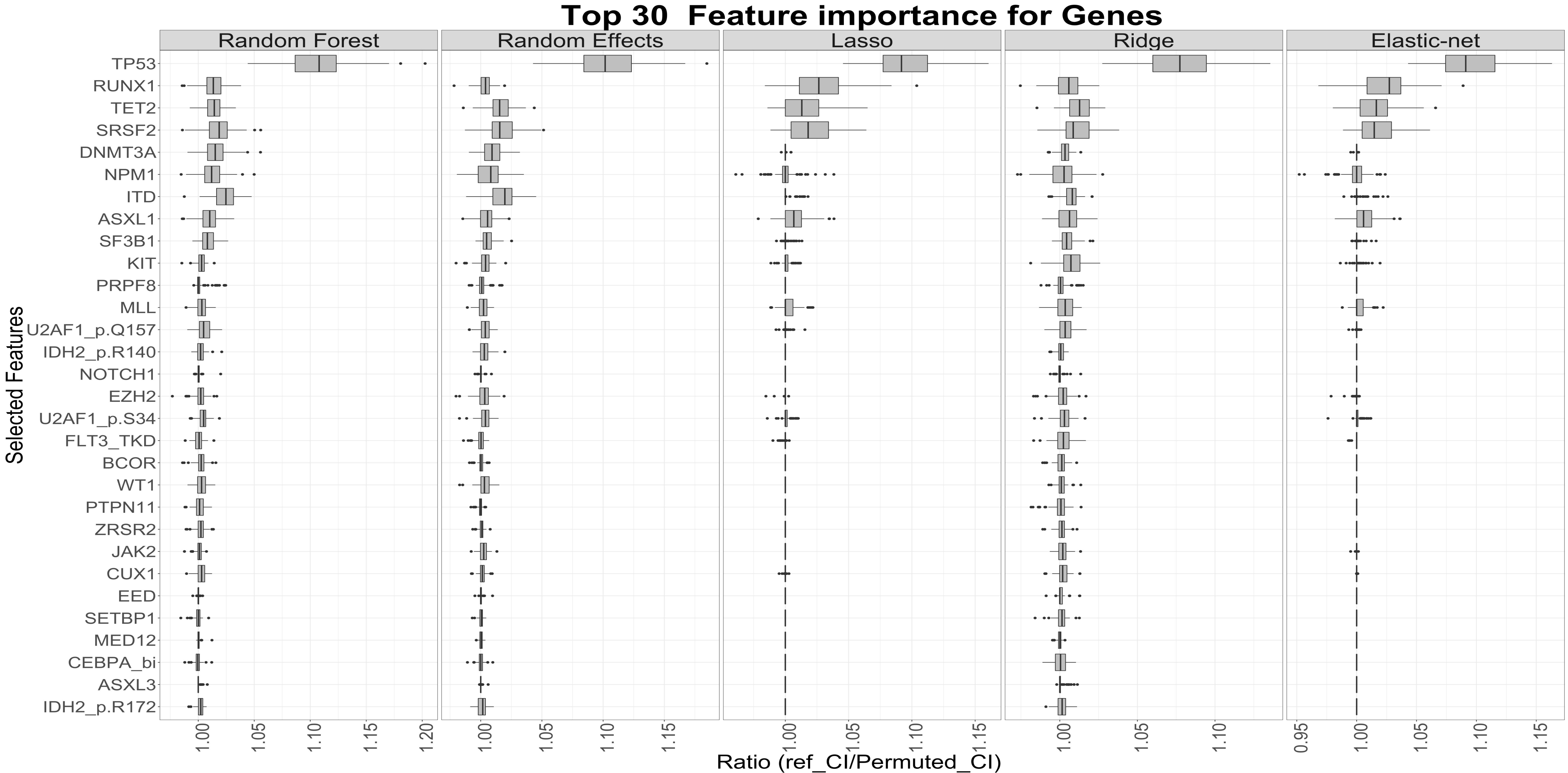

**S.Figure 20: Feature Importance in the class + genes + cytogenetics based model (top 30) in AML NCRI Trial (n=2,113).**
The y axis corresponds to the different features evaluated by the models and ordered by importance. The x axis corresponds to the reshuffling ratio metric for each feature ( metric = reference C-Index / permuted C-Index ). The higher the ratio, the more sensitive the model is to that particular feature. The results are stratified by algorithms (Random Forest, Random Effects, Lasso, Ridge and Elastic-Net). For more details, please refer to S.Appendix.

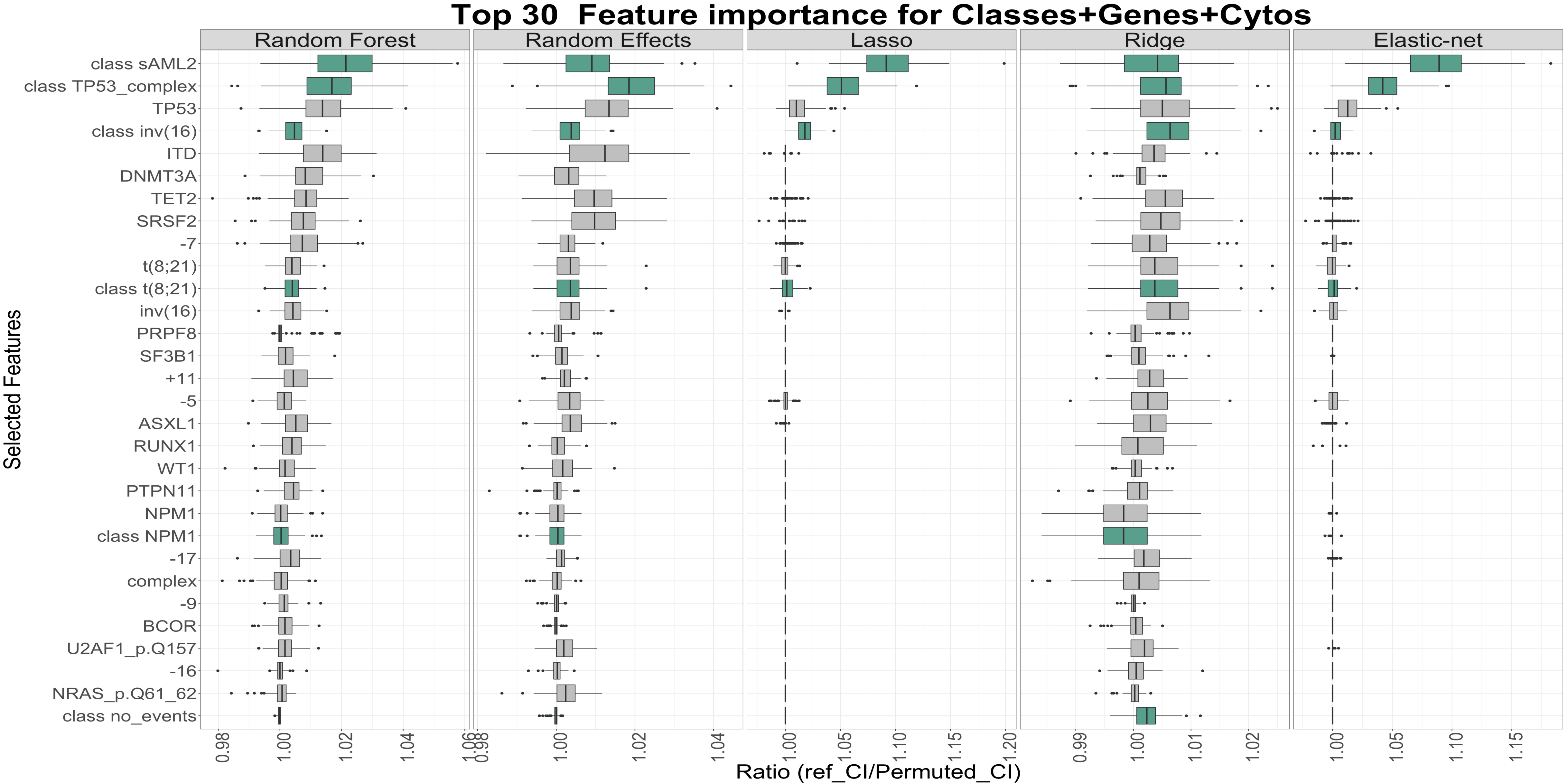

**S.Figure 21: Feature Importance in the ELN + genes + cytos based model (top 30) in AML NCRI Trial (n=2,113).**
The y axis corresponds to the different features evaluated by the models and ordered by importance. The x axis corresponds to the reshuffling ratio metric for each feature ( metric = reference C-Index / permuted C-Index ). The higher the ratio, the more sensitive the model is to that particular feature. The results are stratified by algorithms (Random Forest, Random Effects, Lasso, Ridge and Elastic-Net). For more details, please refer to S.Appendix.

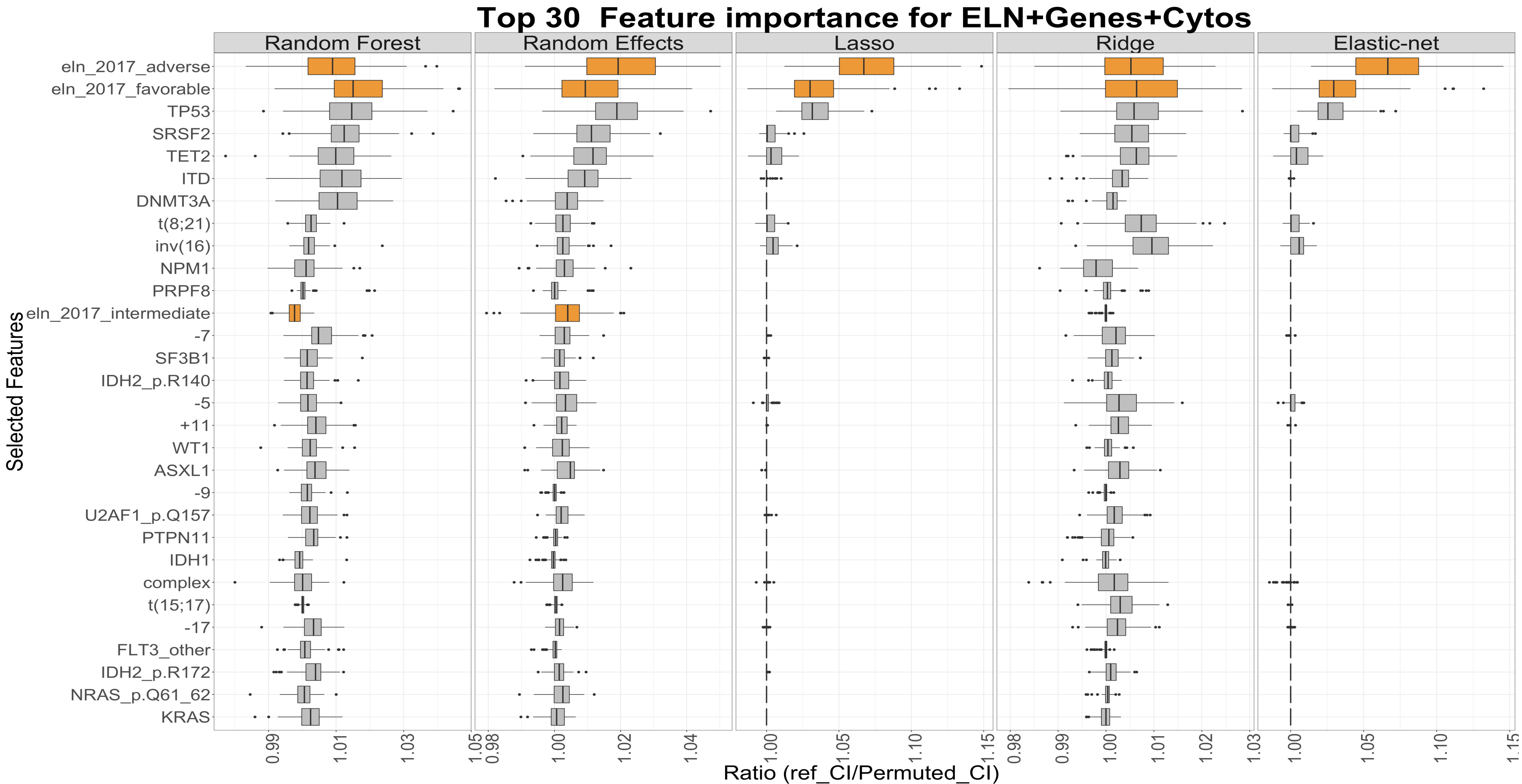

**S.Figure 22: Feature Importance in the ELN + genes based model (top 30) in AML NCRI Trial (n=2,113).**
The y axis corresponds to the different features evaluated by the models and ordered by importance. The x axis corresponds to the reshuffling ratio metric for each feature ( metric = reference C-Index / permuted C-Index ). The higher the ratio, the more sensitive the model is to that particular feature. The results are stratified by algorithms (Random Forest, Random Effects, Lasso, Ridge and Elastic-Net). For more details, please refer to S.Appendix.

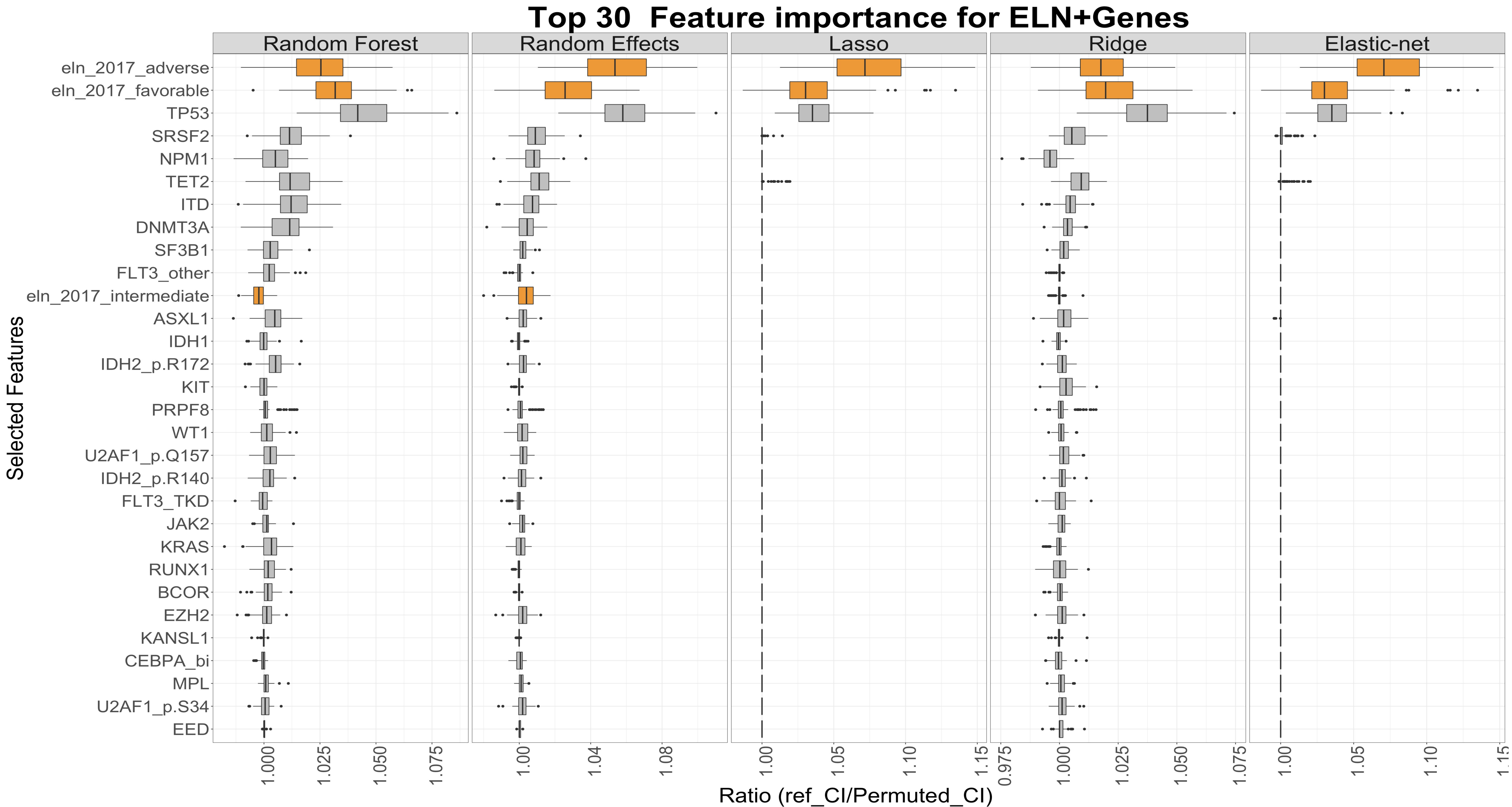

**S.Figure 23: Feature Importance in the full Model (ELN + Genes + Cytos + Clin + Demo) in AML NCRI Trial (n=2,113).**
The y axis corresponds to the different features evaluated by the models and ordered by importance. The x axis corresponds to the reshuffling ratio metric for each feature ( metric = reference C-Index / permuted C-Index ). The higher the ratio, the more sensitive the model is to that particular feature. The results are stratified by algorithms (Random Forest, Random Effects, Lasso, Ridge and Elastic-Net). For more details, please refer to S.Appendix.

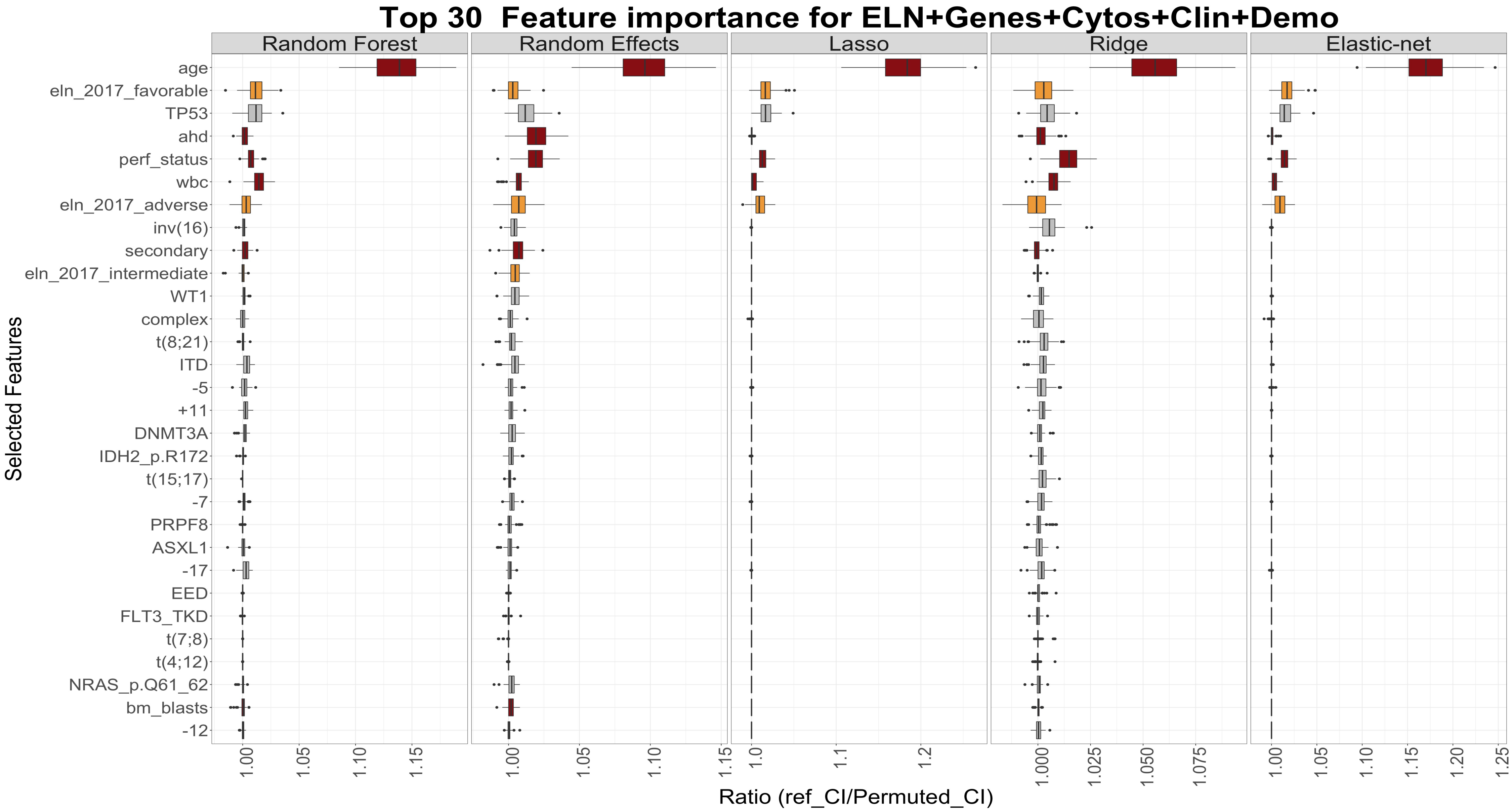

**S.Figure 24: Explained variation and randomness using Nagelkerke's pseudo $R^2$ in the AML NCRI Trial (n=2,113).** A. Explained variation and randomness using different subset of the covariates to include: ELN2017, classes, classes + ITD, genes, clinical data, genes + cytogenetics, classes + clinical data, genes + cytogenetics + clinical data**.** B. Explained variation and randomness for each covariate.
Pseudo $R^2$ are relative measures indicating how well a model/feature explains the data.

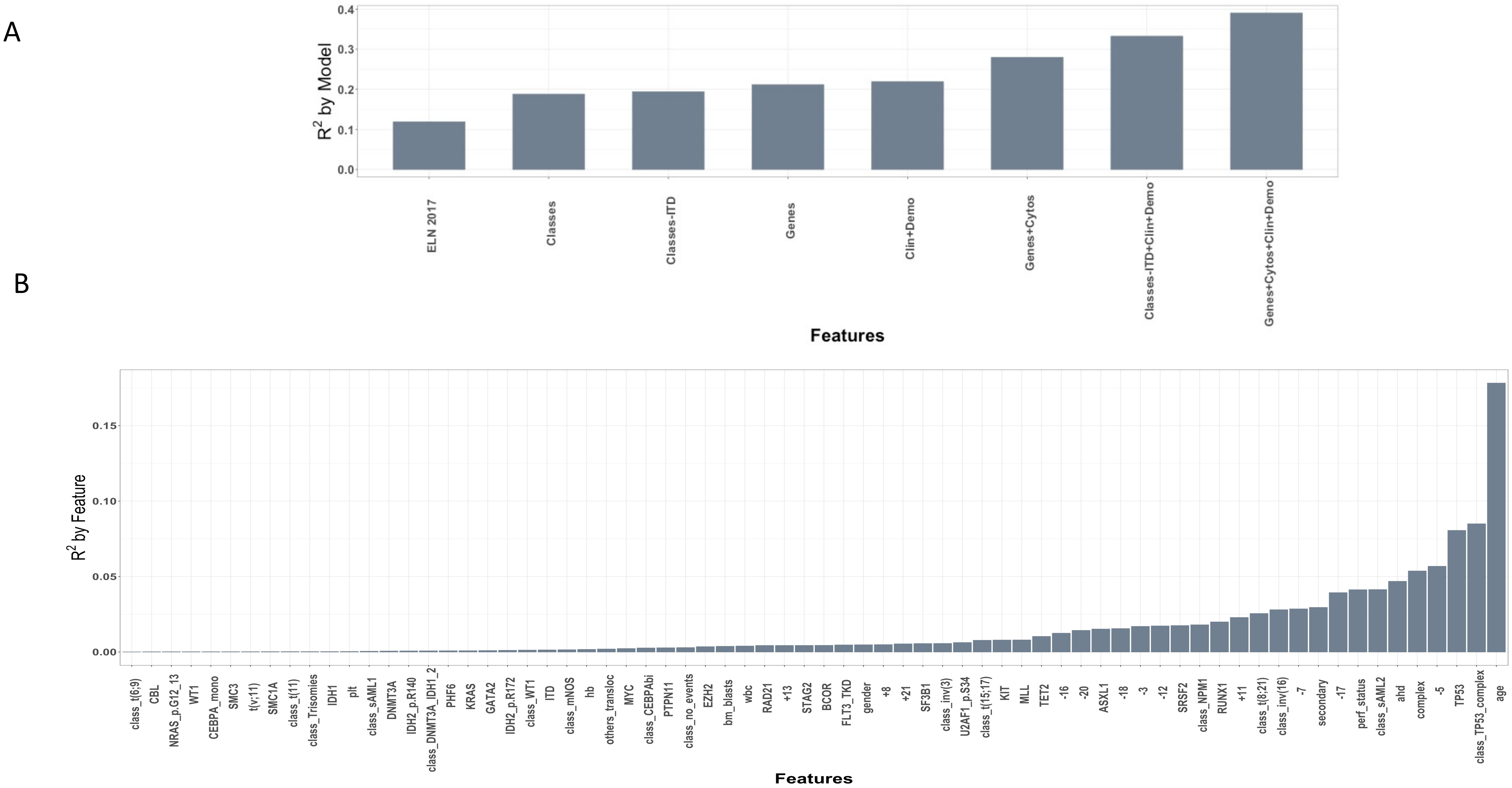

**S.Figure 25: Prognosis Evolution Validation of C-Index in the AML SG Cohort (1,540 patients).**

A. Bar plots of number of features for the different models that were evaluated to include: ELN[2017], classes, classes + ITD, genes, clinical data, genes + cytogenetics, classes + clinical data, genes + cytogenetics + clinical data.

B. Concordance Index (C-Index) measured using a Cox Ridge model on different subsets of the features (ELN[2017], classes, classes + ITD, genes, clinical data, genes + cytogenetics, classes + clinical data, genes + cytogenetics + clinical data) with internal 5 fold cross-validation for the regularization parameter λ. We randomly reshuffled the data (n=1,540) and used 75% for training and cross validation and the remaining 25% for testing. We used 100 Bootstraps iterations on the test set to produce the box plots with 95%confidence interval and we evaluated the pvalues by comparing the differences in C-index distributions on the displayed models. We trained different models (penalized Cox Models, Random Forest, Cox Boosting, Cox Random Effects and Support Vector Machines) and here we display the results for the Ridge Cox Model. For more details, please refer to S.Appendix. The bar plots represent the number of features used to evaluate the C-Index on the different subsets of features.

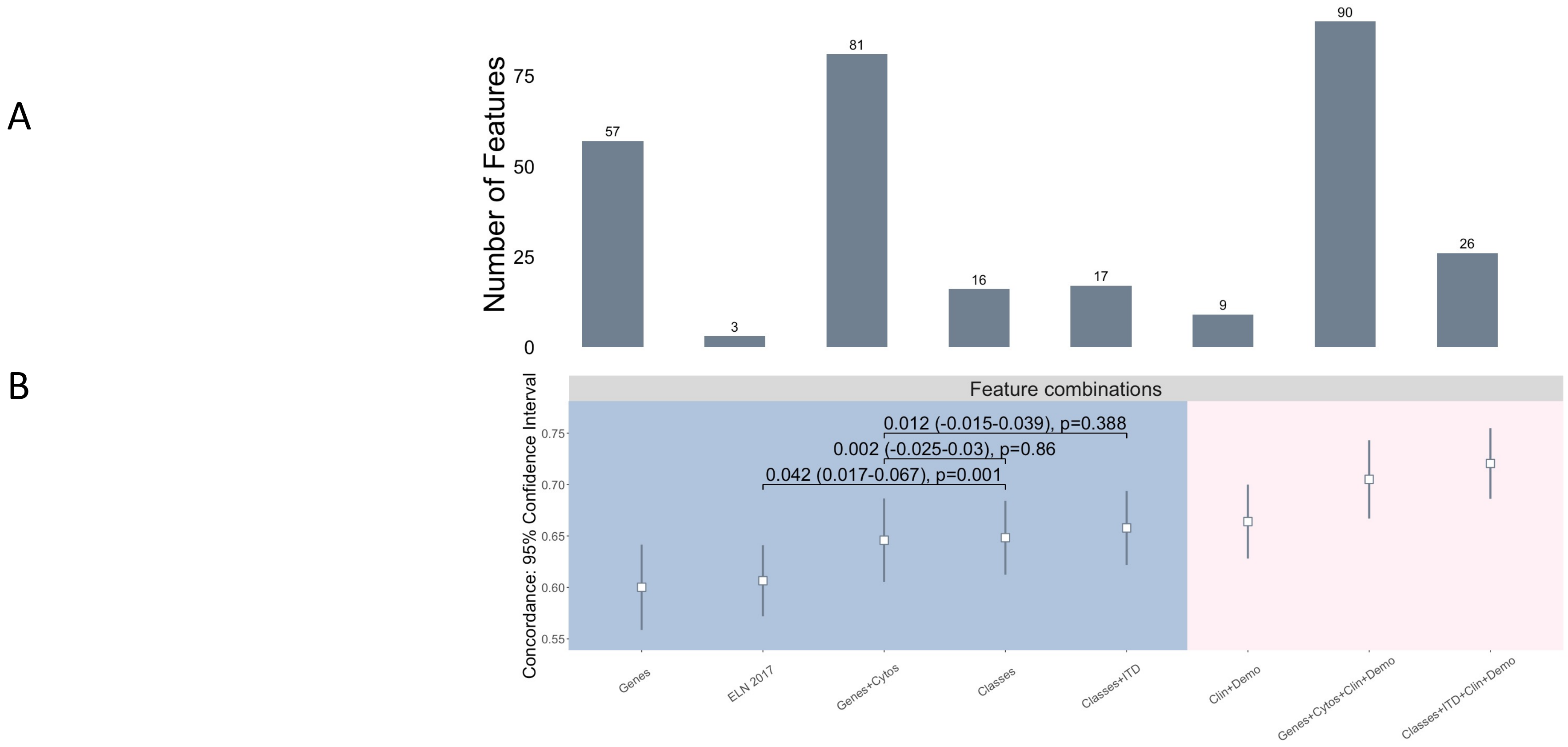

**S.Figure 26: Explained variation and randomness using Nagelkerke $R^2$ in the validation AML SG cohort (n=1,540).** A. Explained variation and randomness using different subset of the covariates to include: $ELN^{2017}$, classes, classes + ITD, genes, clinical data, genes + cytogenetics, classes + clinical data, genes + cytogenetics + clinical data**.** B. Explained variation and randomness for each covariate.
Pseudo $R^2$ are relative measures indicating how well a model/feature explains the data.

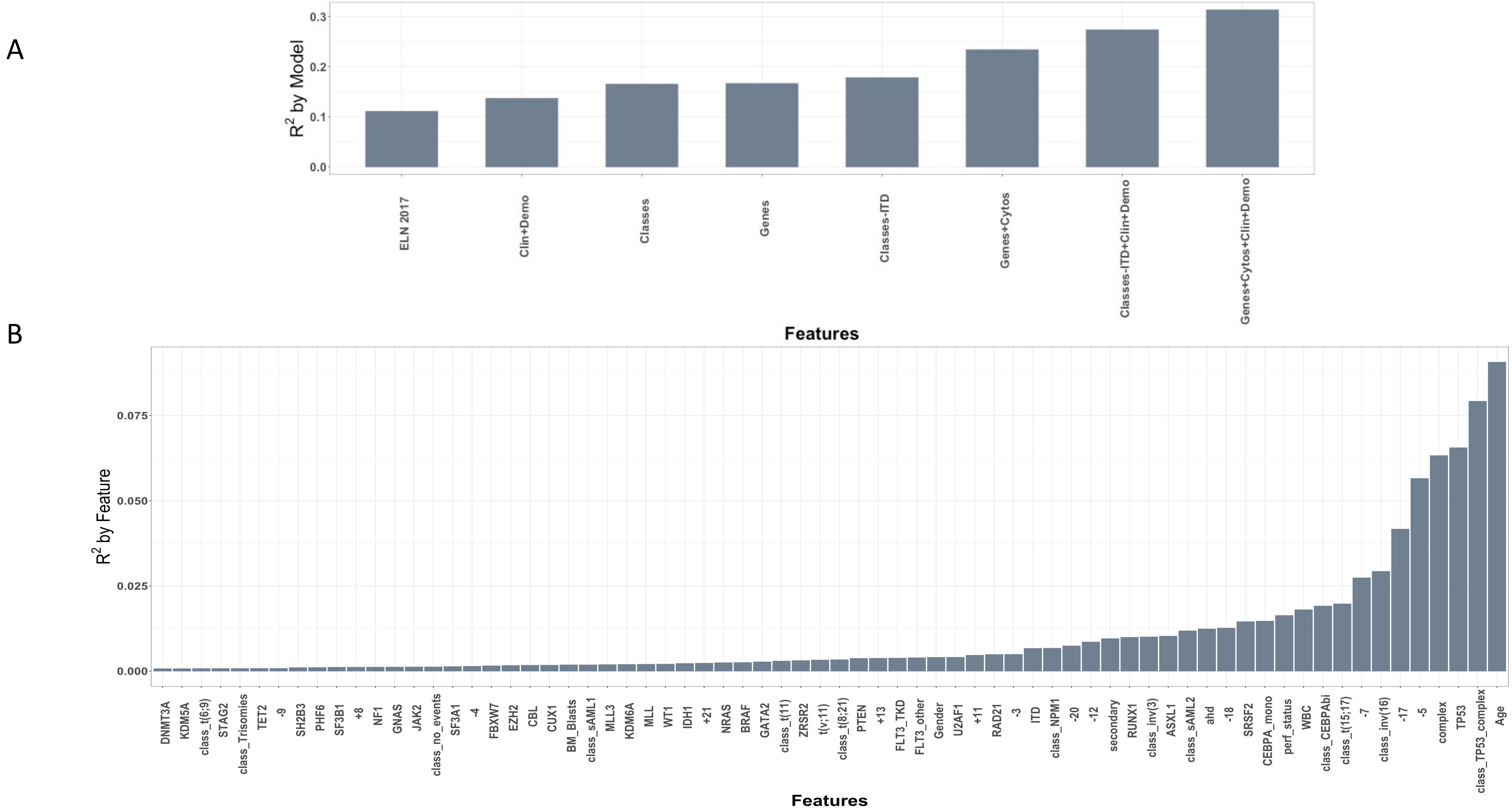

**S.Figure 27: Multi-state model for disease progression in the AML NCRI cohort (n=2,017).**

A. Representation of patient transitions (in numbers) across clinical endpoints (alive (meaning received induction chemotherapy); alive in complete remission; alive in relapse; death without complete remission; death in complete remission; death in relapse). B. Non-parametric multi-state transition probability with 95% confidence bands for the AML NCRI cohort for patients that received intensive treatment (n=1,661). C. Stacked transition probabilities with 95% confidence bands for each class (y-axis) across time (x-axis).

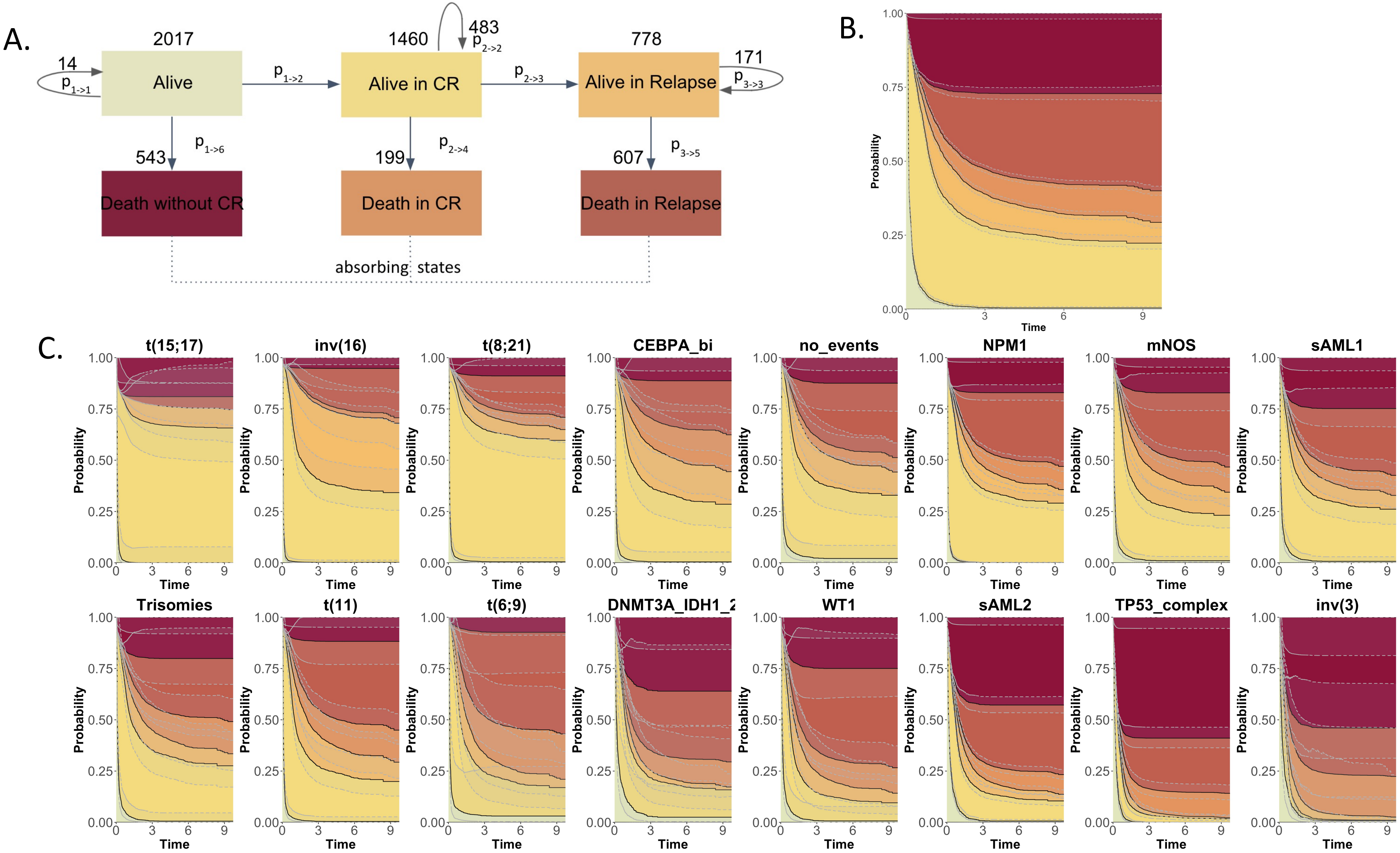

**S.Figure 28: Multi-state model for disease progression for patients that received intensive treatment (n=1,661).**
A. Representation of patient transitions (in numbers) across clinical endpoints (alive (meaning received induction chemotherapy); alive in complete remission; alive in relapse; death without complete remission; death in complete remission; death in relapse). B. Non-parametric multi-state transition probability with 95% confidence bands for the AML NCRI cohort for patients that received intensive treatment (n=1,661). C. Stacked transition probabilities with 95% confidence for each class (y-axis) across time (x-axis).

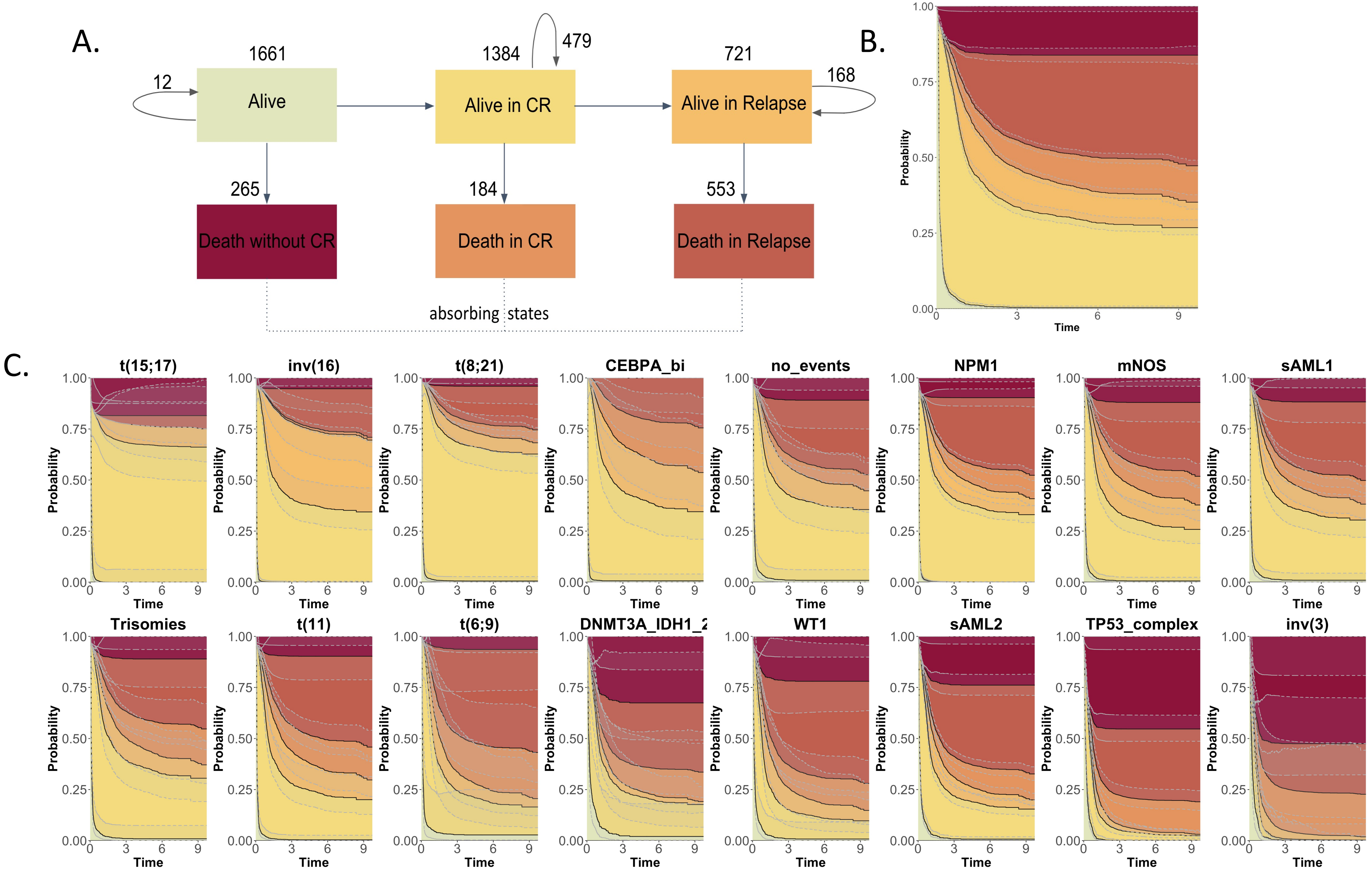

**S.Figure 29: Cumulative incidence and risk outcomes for different subsets in the AML NCRI Trial (n=2,017).**

A. Cumulative incidence curves comparing inv(16) and t(8;21) classes in the AML NCRI Trial (n=2,017). Pvalues were computed with Gray's test.

B. Cumulative incidence curves comparing patients with and without $FLT3^{ITD}$ in the AML NCRI Trial (n=2,017). Pvalues were computed with Gray's test.

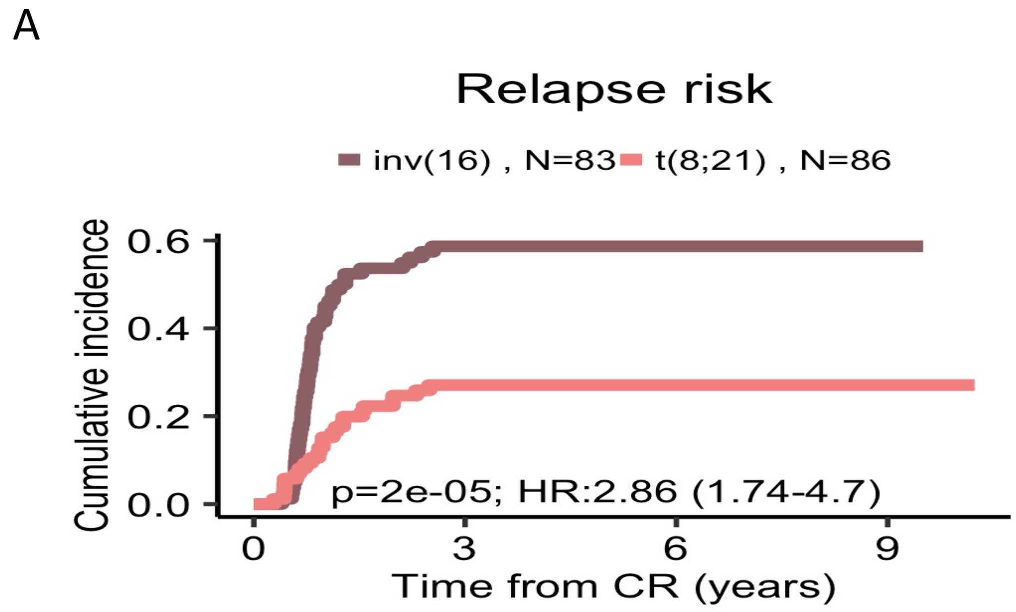

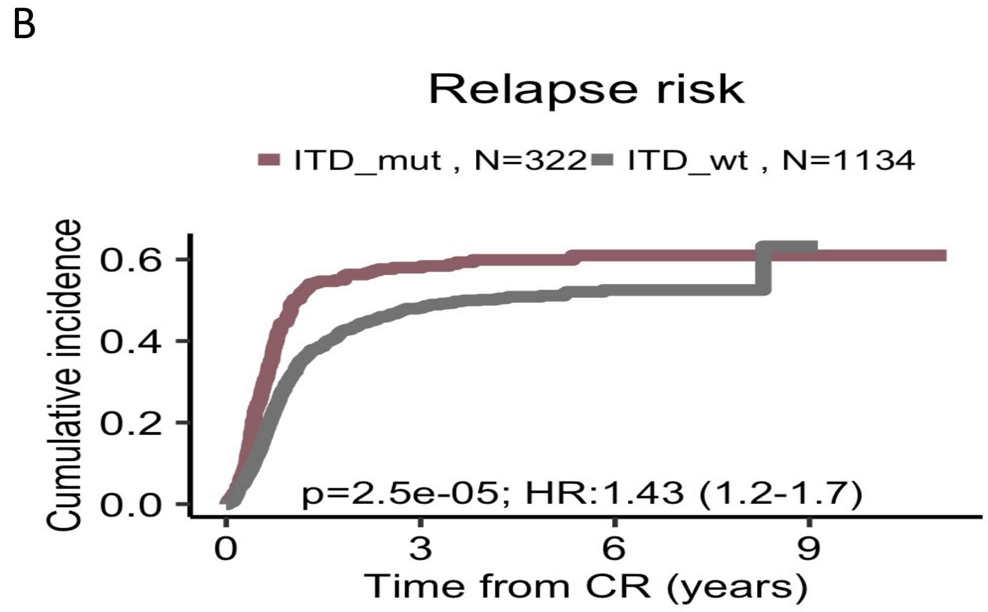

**S.Figure 30: Multi-state semi-parametric Cox model incorporating FLT3$^{ITD}$ shift for relevant components in the AML NCRI Trial (n=2,017).** This is a semi-parametric Cox multi-state transition probability plot with 95% confidence bands for selected classes with and without the presence of FLT3$^{ITD}$. The model contains 6 possible states: alive (meaning received induction chemotherapy); alive in complete remission; alive in relapse; death without complete remission; death in complete remission; death in relapse. For more details about the semi-parametric transition hazard model, please refer to S.Appendix. The bold lines represent the death states.

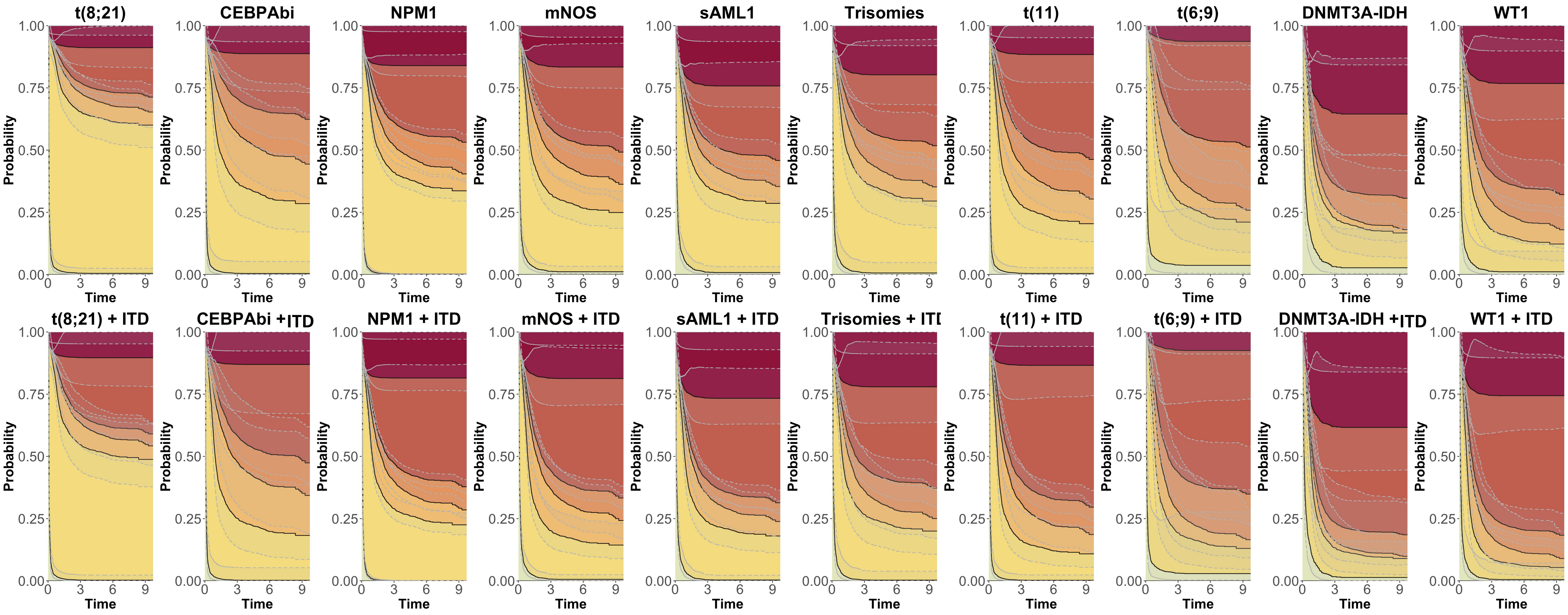

**S.Figure 31: Validation of multi-state model in the validation AML SG Cohort (n=1,540).** A. Multi-state transitions with number of patients in each possible state in the AML SG Cohort (n=1,540). The model contains 6 possible states: alive (meaning received induction chemotherapy); alive in complete remission; alive in relapse; death without complete remission; death in complete remission; death in relapse. B. Non-parametric multi-state transition probability with 95% confidence bands for the overall AML SG Cohort (n=1,540). C. Multi-state semi-parametric transition probabilities with 95% confidence bands for each class in the AML SG Cohort (n=1,540). The bold lines represent the death states. For more details about the semi-parametric transition hazard model, please refer to S.Appendix.

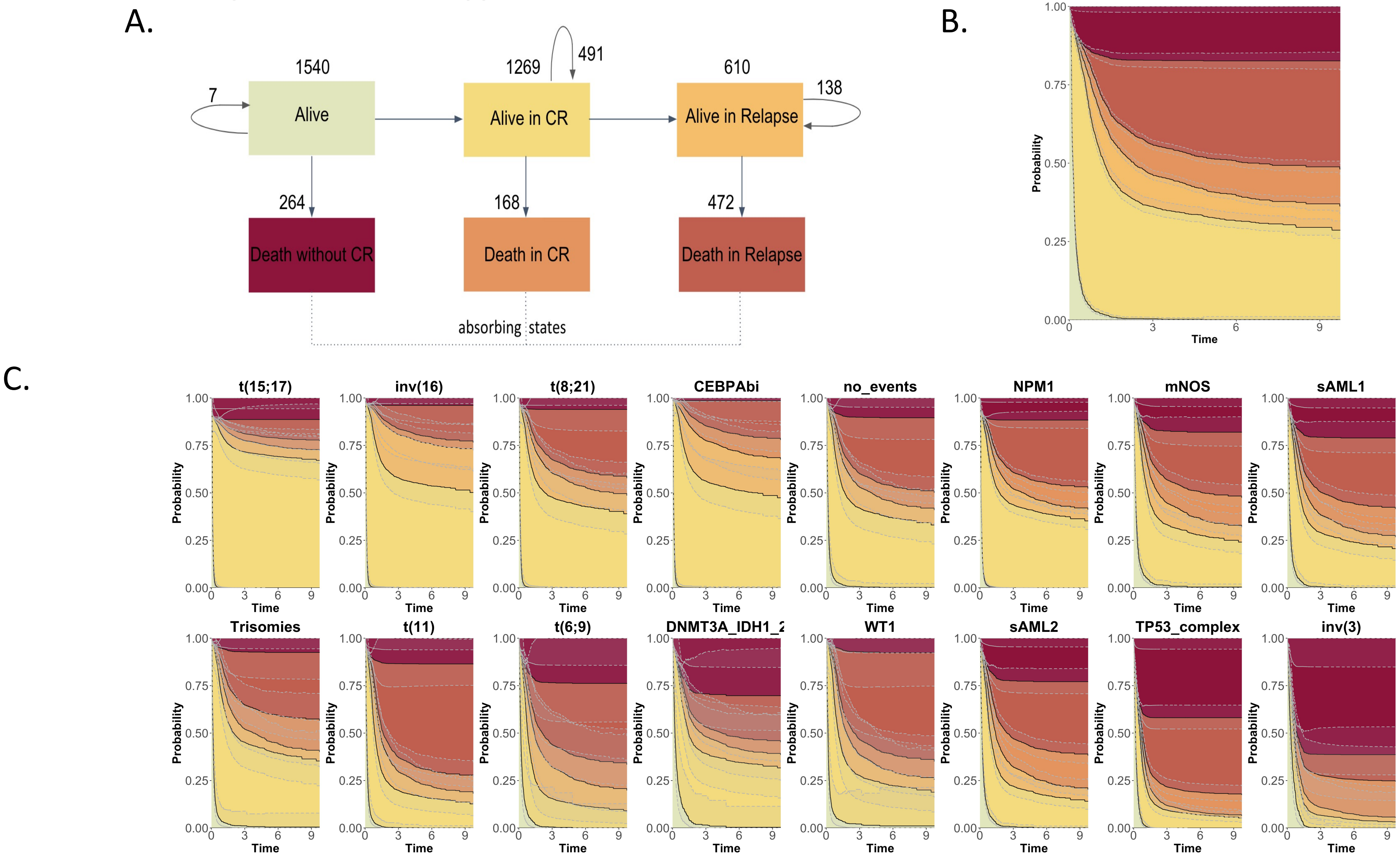

**S. Figure 32: Kaplan-Meier overall survival curves, cumulative incidence of relapse and associated risk tables** for patients that attained CR in AML17 trial subset, stratified by MRD status post course 1 (n=523). Pvalues were computed with Gray's test and  log-rank test.

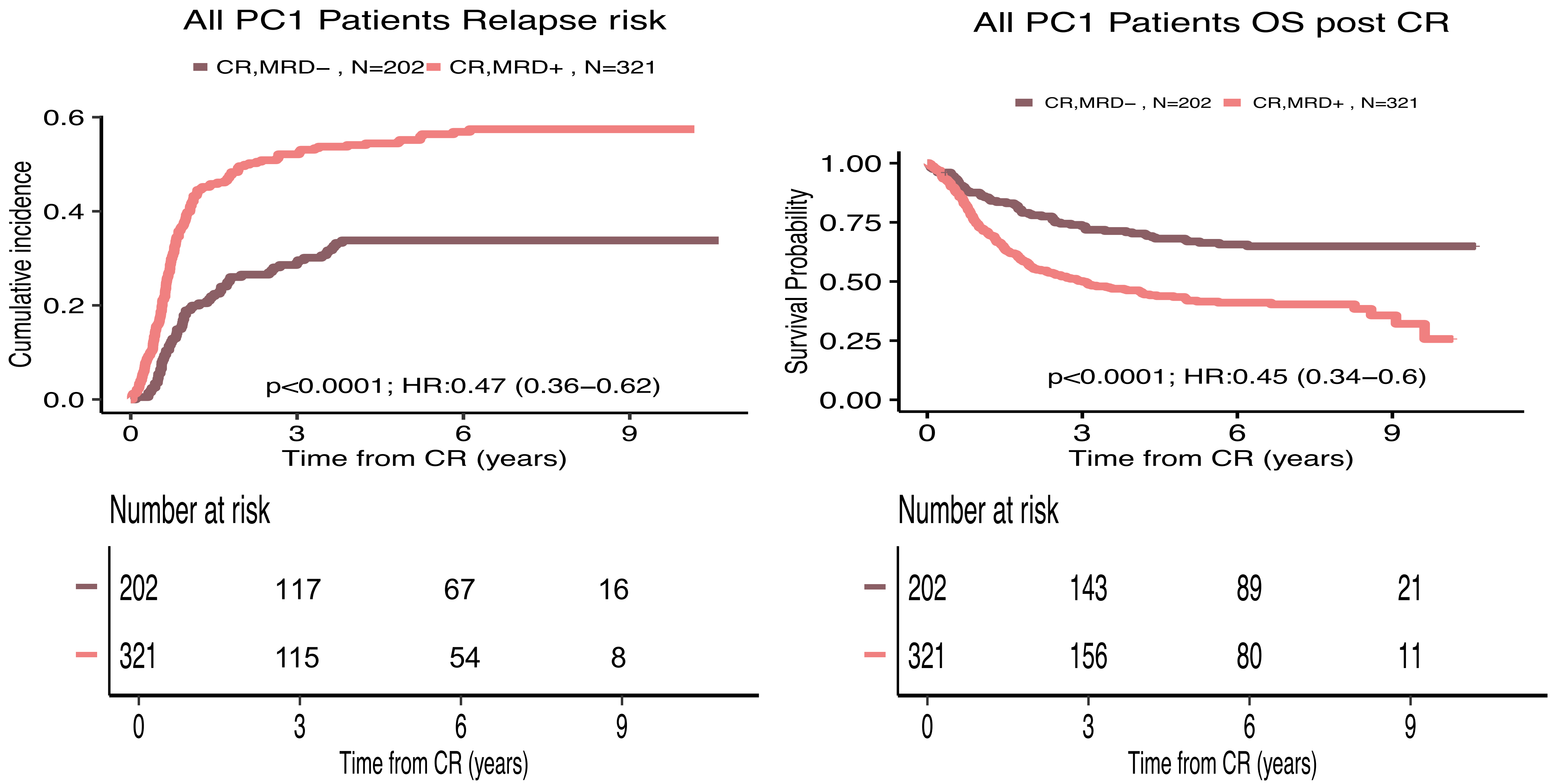

**S. Figure 33: Kaplan-Meier overall survival curves, cumulative incidence of relapse and associated risk tables for the sAML2, sAML1 subgroups** that attained CR in AML17, stratified by MRD status post course 1. Pvalues were computed with Gray's test and log-rank test.

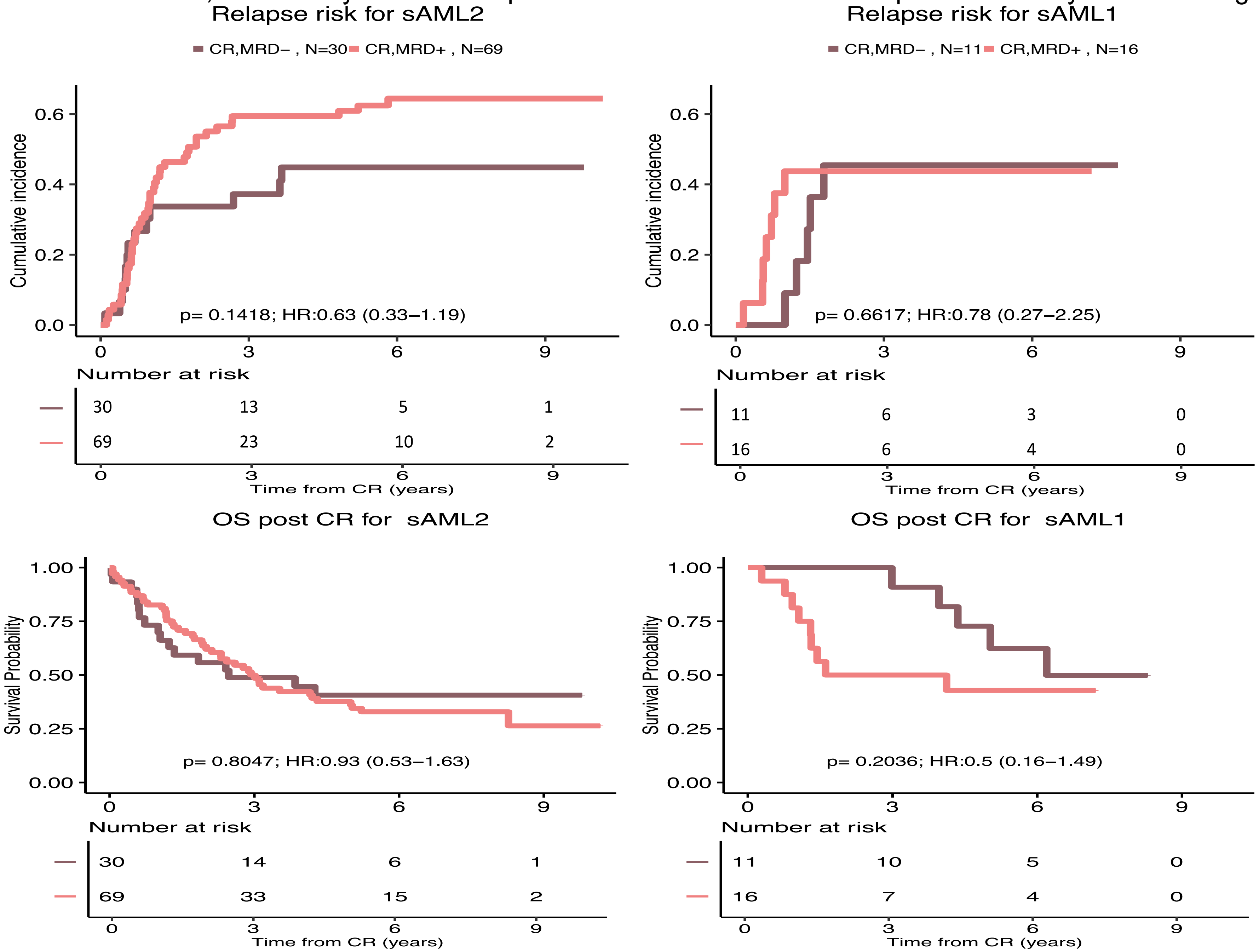

**S.Figure 34: Summary by class for cumulative incidence and Kaplan-Meier curves for survival post complete remission on AML 17 NCRI Trial Cohort with post course 1 minimal residual disease (MRD) analysis analysis (n=523).** The first panel represents the cumulative incidence of relapse for patients in that specific class stratified by MRD status (p values were computed with Gray test) and the second panel represents the Kaplan-Meier curve for survival post course 1 complete remission stratified by MRD status. We omitted classes with less than 3 patients with MRD positive or negative. Panels where comparator group has less than 10 patients have been removed for robust statistical interpretation.

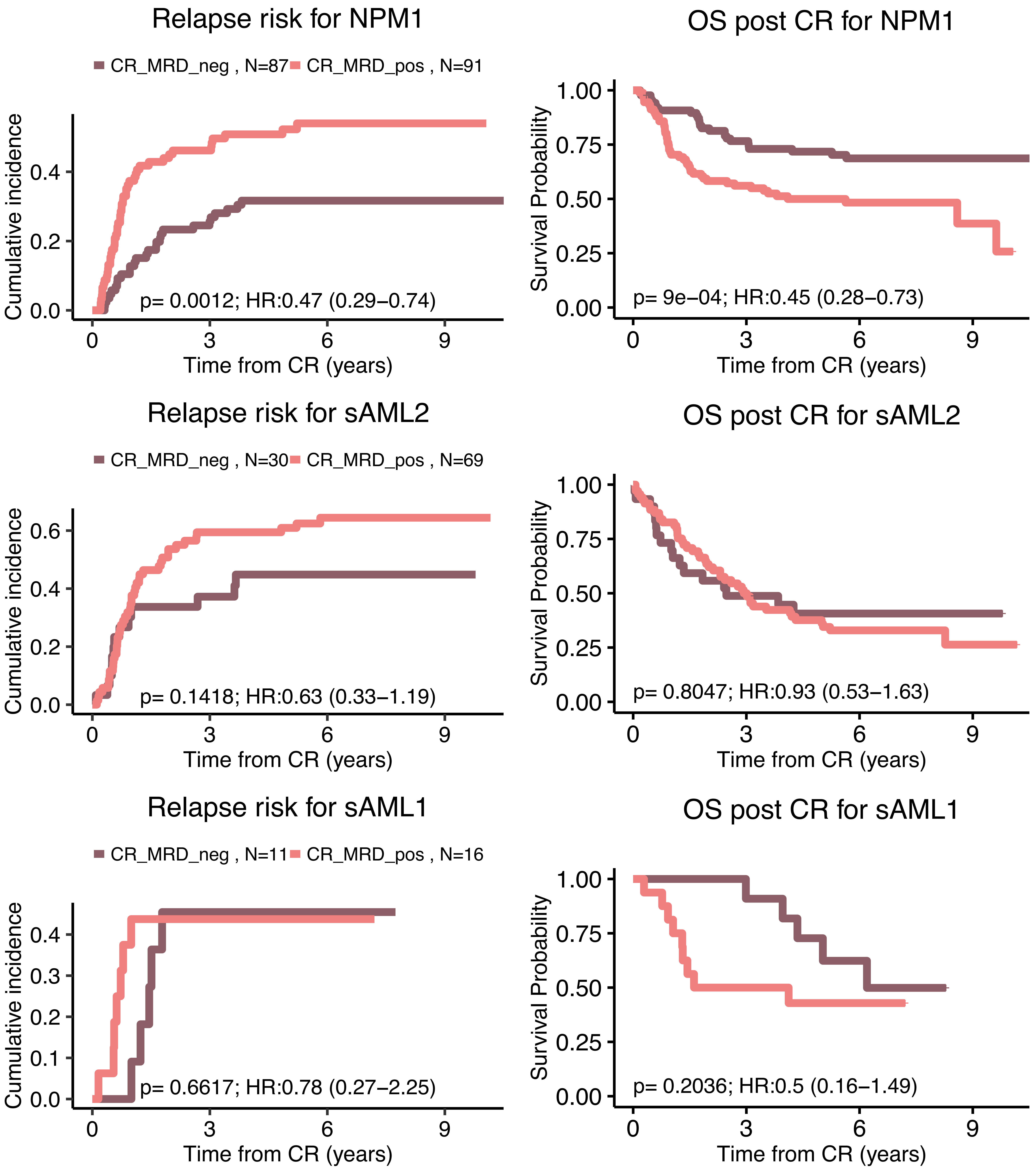
Relapse risk for NPM1
CR_MRD_neg , N=87
CR_MRD_pos , N=91
Cumulative incidence
0.0
0.2
0.4
p= 0.0012; HR:0.47 (0.29–0.74)
0
3
6
9
Time from CR (years)
OS post CR for NPM1
Survival Probability
0.00
0.25
0.50
0.75
1.00
p= 9e–04; HR:0.45 (0.28–0.73)
0
3
6
9
Time from CR (years)
Relapse risk for sAML2
CR_MRD_neg , N=30
CR_MRD_pos , N=69
Cumulative incidence
0.0
0.2
0.4
0.6
p= 0.1418; HR:0.63 (0.33–1.19)
0
3
6
9
Time from CR (years)
OS post CR for sAML2
Survival Probability
0.00
0.25
0.50
0.75
1.00
p= 0.8047; HR:0.93 (0.53–1.63)
0
3
6
9
Time from CR (years)
Relapse risk for sAML1
CR_MRD_neg , N=11
CR_MRD_pos , N=16
Cumulative incidence
0.0
0.1
0.2
0.3
0.4
p= 0.6617; HR:0.78 (0.27–2.25)
0
3
6
9
Time from CR (years)
OS post CR for sAML1
Survival Probability
0.00
0.25
0.50
0.75
1.00
p= 0.2036; HR:0.5 (0.16–1.49)
0
3
6
9
Time from CR (years)

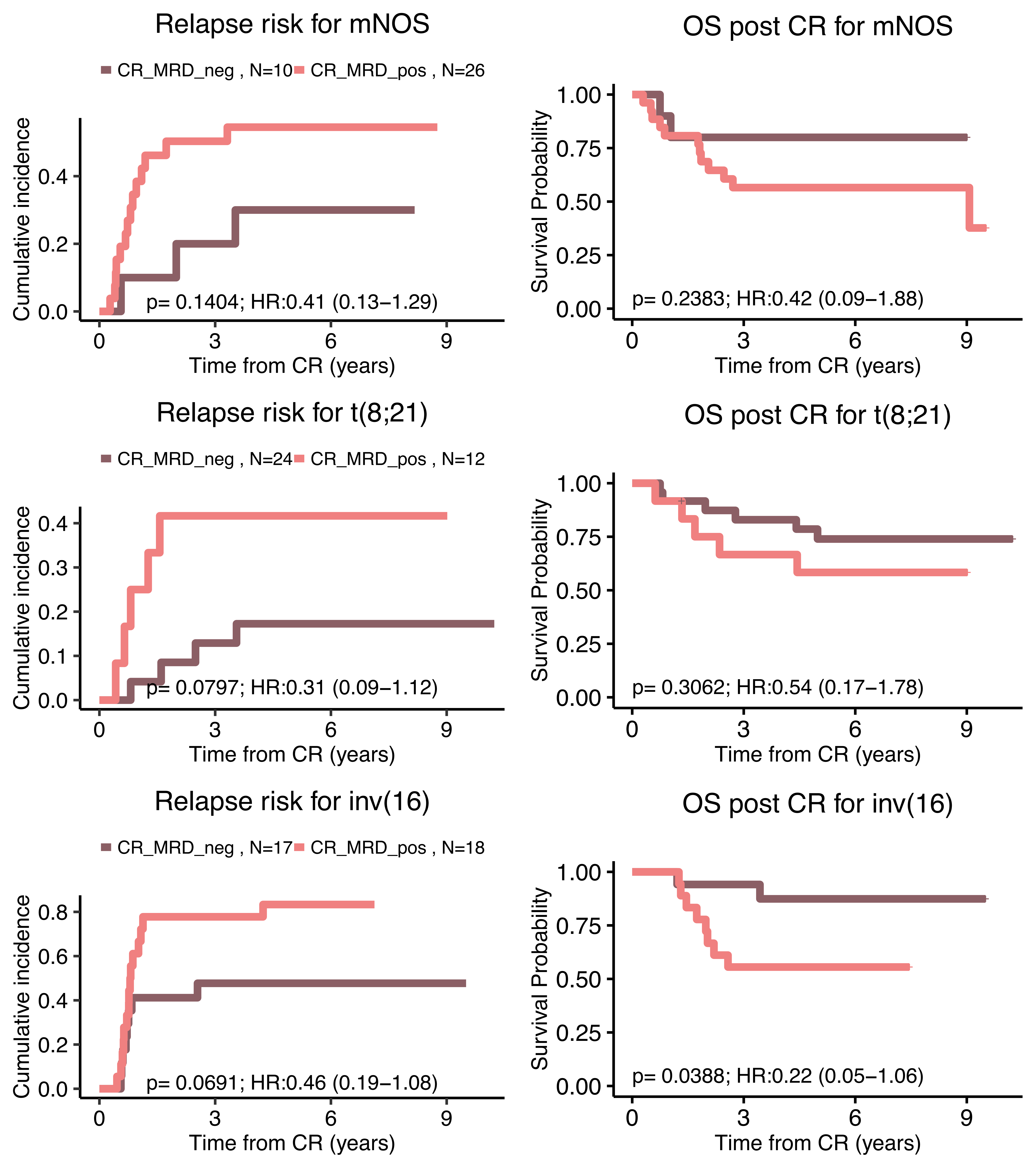
Relapse risk for mNOS
CR_MRD_neg , N=10
CR_MRD_pos , N=26
Cumulative incidence
p= 0.1404; HR:0.41 (0.13−1.29)
Time from CR (years)
OS post CR for mNOS
Survival Probability
p= 0.2383; HR:0.42 (0.09−1.88)
Time from CR (years)
Relapse risk for t(8;21)
CR_MRD_neg , N=24
CR_MRD_pos , N=12
Cumulative incidence
p= 0.0797; HR:0.31 (0.09−1.12)
Time from CR (years)
OS post CR for t(8;21)
Survival Probability
p= 0.3062; HR:0.54 (0.17−1.78)
Time from CR (years)
Relapse risk for inv(16)
CR_MRD_neg , N=17
CR_MRD_pos , N=18
Cumulative incidence
p= 0.0691; HR:0.46 (0.19−1.08)
Time from CR (years)
OS post CR for inv(16)
Survival Probability
p= 0.0388; HR:0.22 (0.05−1.06)
Time from CR (years)

**S.Figure 35: Summary by genes mutations and cytogenetics abnormalities for cumulative incidence and Kaplan-Meier curves for survival post complete remission on AML 17 NCRI Trial Cohort with post course 1 minimal residual disease (MRD) analysis (n=523).** The first panel represents the cumulative incidence of relapse for patients in that specific event stratified by MRD status (p values were computed with Gray test) and the second panel represents the Kaplan-Meier curve for survival post course 1 complete remission stratified by MRD status. We omitted events with less than 5 patients with MRD positive or negative.
Panels where comparator group has less than 10 patients have been removed for robust statistical interpretation.

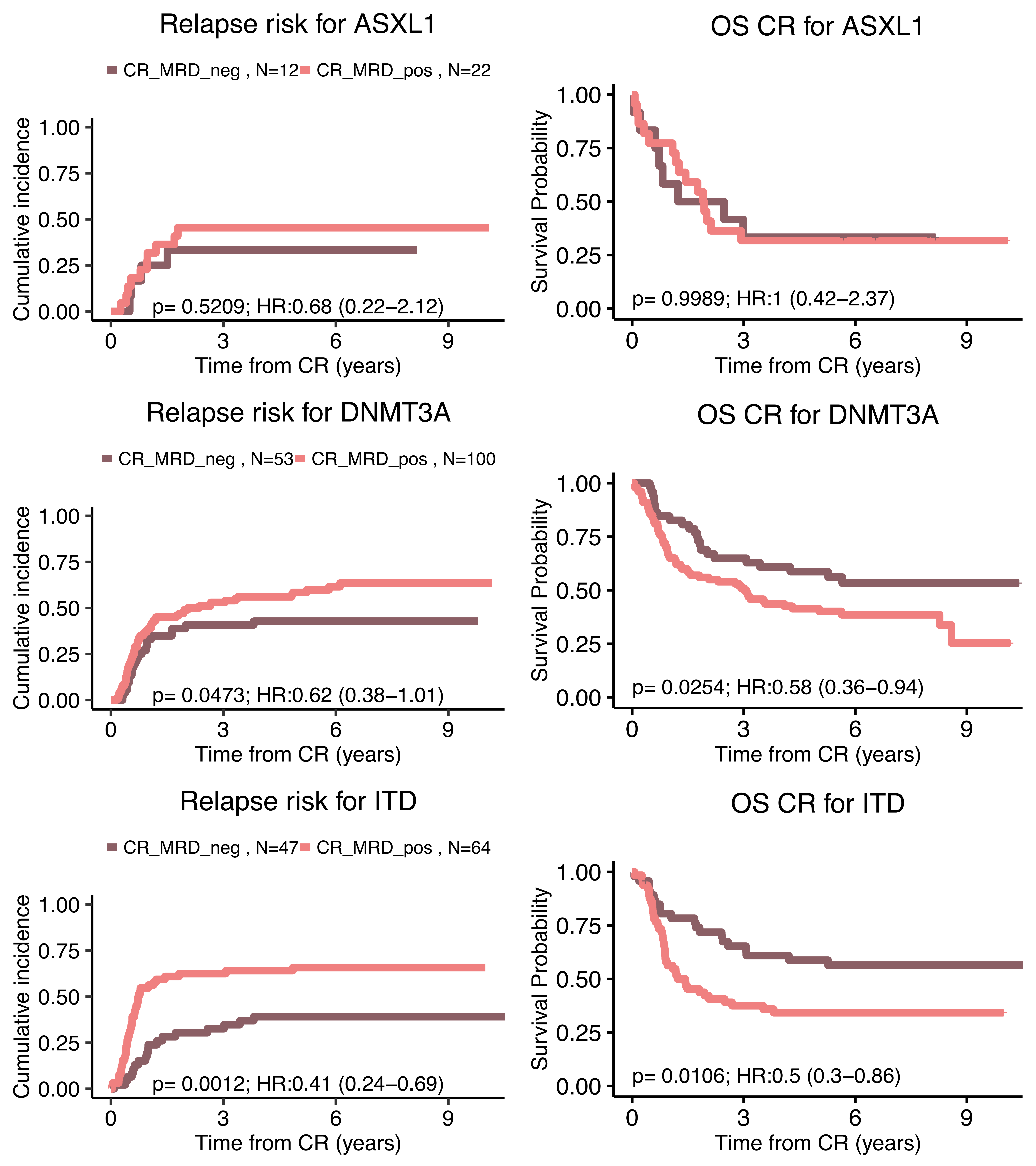
Relapse risk for ASXL1
CR_MRD_neg , N=12
CR_MRD_pos , N=22
Cumulative incidence
1.00
0.75
0.50
0.25
0.00
p= 0.5209; HR:0.68 (0.22–2.12)
0
3
6
9
Time from CR (years)
OS CR for ASXL1
Survival Probability
1.00
0.75
0.50
0.25
0.00
p= 0.9989; HR:1 (0.42–2.37)
0
3
6
9
Time from CR (years)
Relapse risk for DNMT3A
CR_MRD_neg , N=53
CR_MRD_pos , N=100
Cumulative incidence
1.00
0.75
0.50
0.25
0.00
p= 0.0473; HR:0.62 (0.38–1.01)
0
3
6
9
Time from CR (years)
OS CR for DNMT3A
Survival Probability
1.00
0.75
0.50
0.25
0.00
p= 0.0254; HR:0.58 (0.36–0.94)
0
3
6
9
Time from CR (years)
Relapse risk for ITD
CR_MRD_neg , N=47
CR_MRD_pos , N=64
Cumulative incidence
1.00
0.75
0.50
0.25
0.00
p= 0.0012; HR:0.41 (0.24–0.69)
0
3
6
9
Time from CR (years)
OS CR for ITD
Survival Probability
1.00
0.75
0.50
0.25
0.00
p= 0.0106; HR:0.5 (0.3–0.86)
0
3
6
9
Time from CR (years)

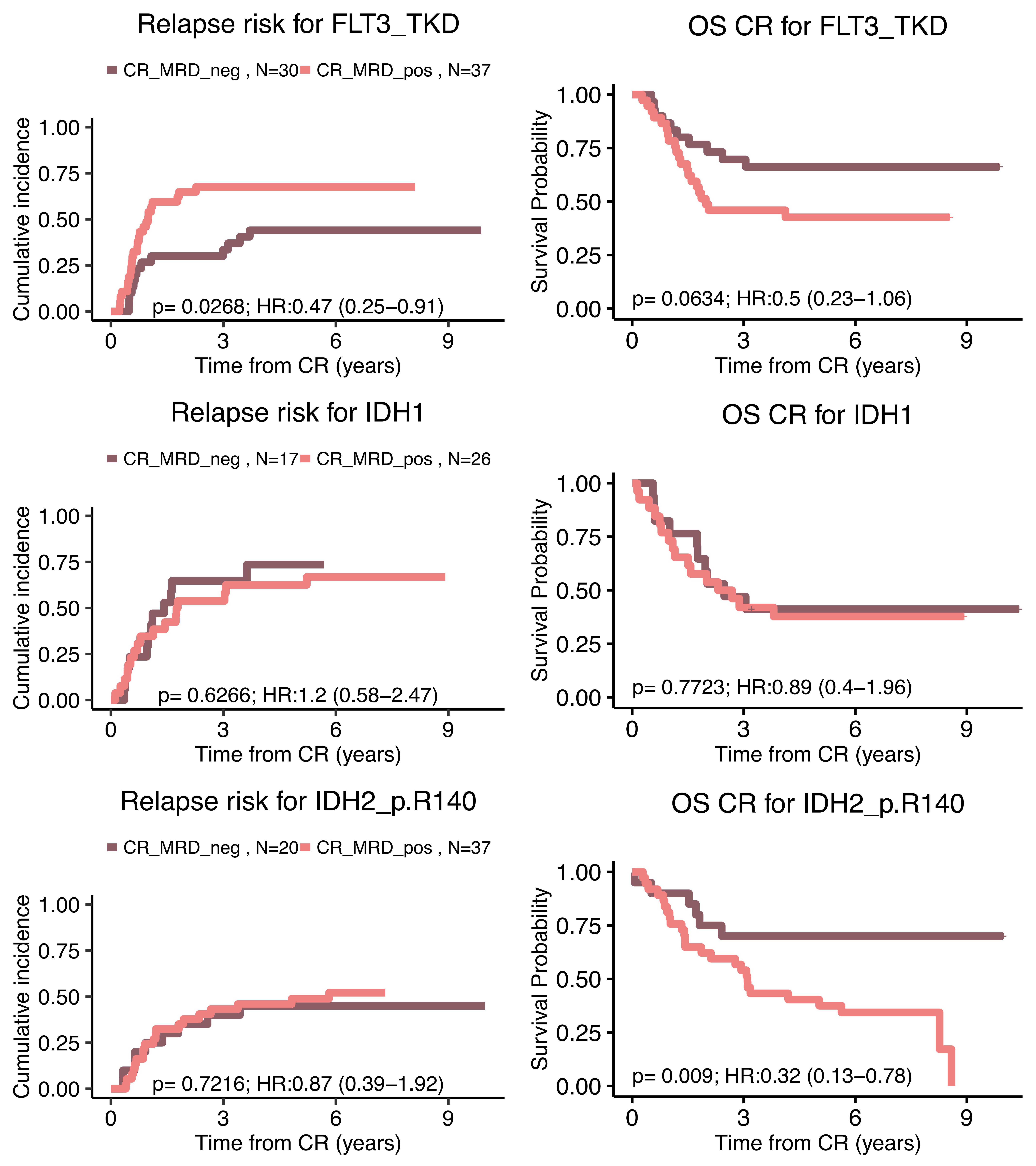
Relapse risk for FLT3_TKD
CR_MRD_neg , N=30
CR_MRD_pos , N=37
Cumulative incidence
Time from CR (years)
p= 0.0268; HR:0.47 (0.25–0.91)
OS CR for FLT3_TKD
Survival Probability
Time from CR (years)
p= 0.0634; HR:0.5 (0.23–1.06)
Relapse risk for IDH1
CR_MRD_neg , N=17
CR_MRD_pos , N=26
Cumulative incidence
Time from CR (years)
p= 0.6266; HR:1.2 (0.58–2.47)
OS CR for IDH1
Survival Probability
Time from CR (years)
p= 0.7723; HR:0.89 (0.4–1.96)
Relapse risk for IDH2_p.R140
CR_MRD_neg , N=20
CR_MRD_pos , N=37
Cumulative incidence
Time from CR (years)
p= 0.7216; HR:0.87 (0.39–1.92)
OS CR for IDH2_p.R140
Survival Probability
Time from CR (years)
p= 0.009; HR:0.32 (0.13–0.78)

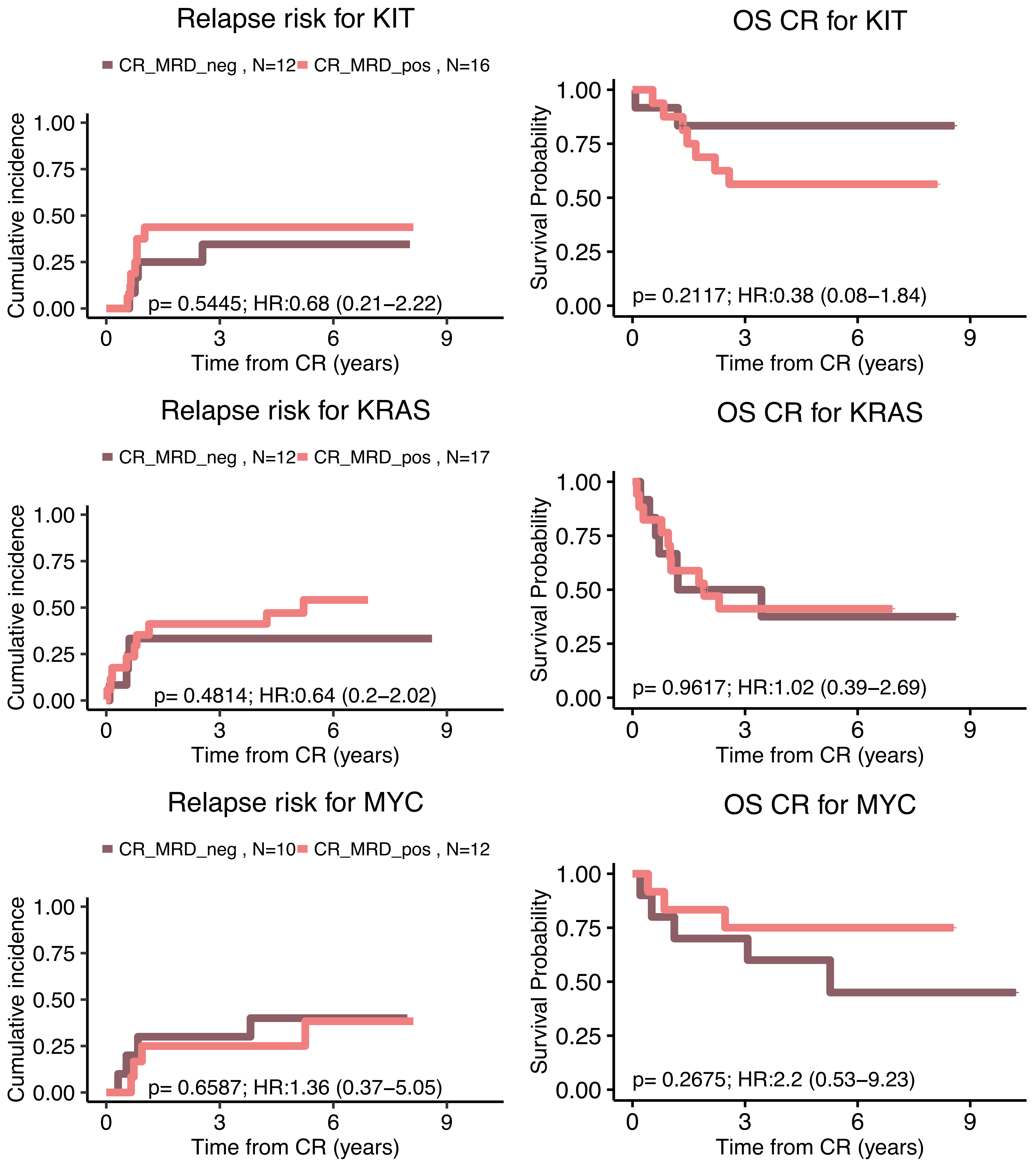
Relapse risk for KIT
CR_MRD_neg , N=12
CR_MRD_pos , N=16
Cumulative incidence
1.00
0.75
0.50
0.25
0.00
p= 0.5445; HR:0.68 (0.21–2.22)
0
3
6
9
Time from CR (years)
OS CR for KIT
Survival Probability
1.00
0.75
0.50
0.25
0.00
p= 0.2117; HR:0.38 (0.08–1.84)
0
3
6
9
Time from CR (years)
Relapse risk for KRAS
CR_MRD_neg , N=12
CR_MRD_pos , N=17
Cumulative incidence
1.00
0.75
0.50
0.25
0.00
p= 0.4814; HR:0.64 (0.2–2.02)
0
3
6
9
Time from CR (years)
OS CR for KRAS
Survival Probability
1.00
0.75
0.50
0.25
0.00
p= 0.9617; HR:1.02 (0.39–2.69)
0
3
6
9
Time from CR (years)
Relapse risk for MYC
CR_MRD_neg , N=10
CR_MRD_pos , N=12
Cumulative incidence
1.00
0.75
0.50
0.25
0.00
p= 0.6587; HR:1.36 (0.37–5.05)
0
3
6
9
Time from CR (years)
OS CR for MYC
Survival Probability
1.00
0.75
0.50
0.25
0.00
p= 0.2675; HR:2.2 (0.53–9.23)
0
3
6
9
Time from CR (years)

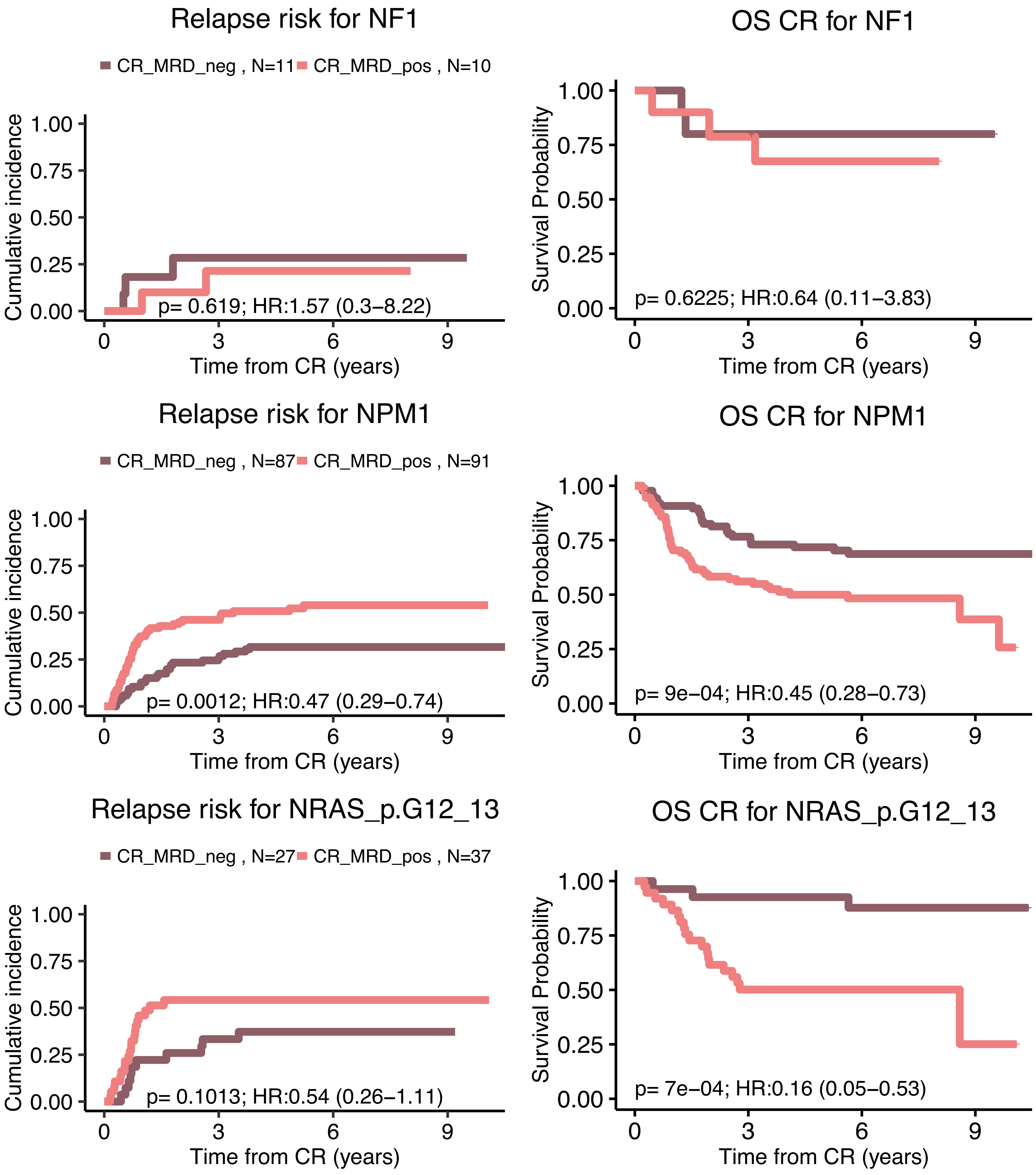
Relapse risk for NF1
CR_MRD_neg , N=11
CR_MRD_pos , N=10
Cumulative incidence
p= 0.619; HR:1.57 (0.3–8.22)
Time from CR (years)
OS CR for NF1
Survival Probability
p= 0.6225; HR:0.64 (0.11–3.83)
Time from CR (years)
Relapse risk for NPM1
CR_MRD_neg , N=87
CR_MRD_pos , N=91
Cumulative incidence
p= 0.0012; HR:0.47 (0.29–0.74)
Time from CR (years)
OS CR for NPM1
Survival Probability
p= 9e−04; HR:0.45 (0.28–0.73)
Time from CR (years)
Relapse risk for NRAS_p.G12_13
CR_MRD_neg , N=27
CR_MRD_pos , N=37
Cumulative incidence
p= 0.1013; HR:0.54 (0.26–1.11)
Time from CR (years)
OS CR for NRAS_p.G12_13
Survival Probability
p= 7e−04; HR:0.16 (0.05–0.53)
Time from CR (years)

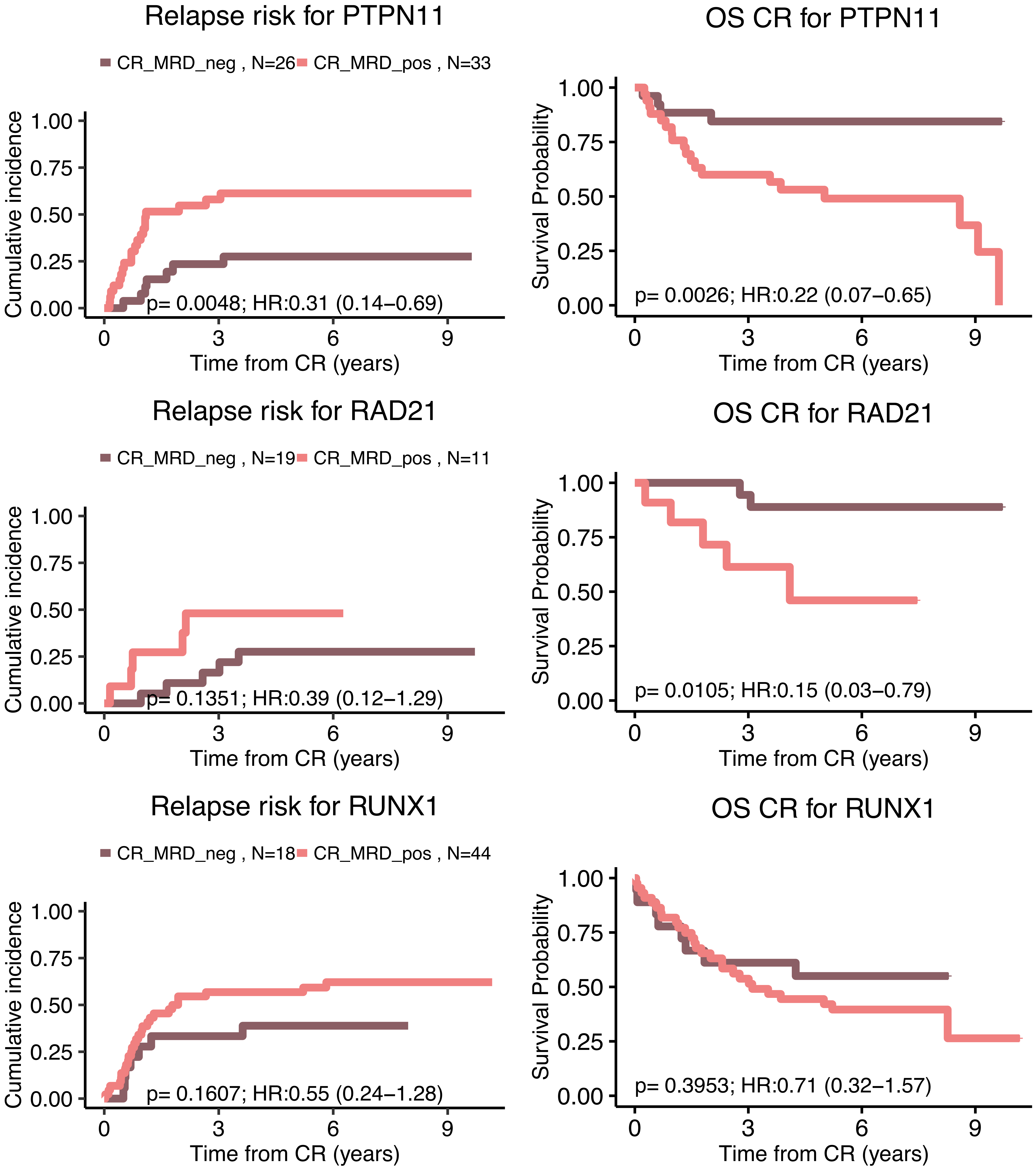
Relapse risk for PTPN11
CR_MRD_neg , N=26
CR_MRD_pos , N=33
Cumulative incidence
1.00
0.75
0.50
0.25
0.00
p= 0.0048; HR:0.31 (0.14–0.69)
0
3
6
9
Time from CR (years)
OS CR for PTPN11
Survival Probability
p= 0.0026; HR:0.22 (0.07–0.65)
Time from CR (years)
Relapse risk for RAD21
CR_MRD_neg , N=19
CR_MRD_pos , N=11
Cumulative incidence
p= 0.1351; HR:0.39 (0.12–1.29)
Time from CR (years)
OS CR for RAD21
Survival Probability
p= 0.0105; HR:0.15 (0.03–0.79)
Time from CR (years)
Relapse risk for RUNX1
CR_MRD_neg , N=18
CR_MRD_pos , N=44
Cumulative incidence
p= 0.1607; HR:0.55 (0.24–1.28)
Time from CR (years)
OS CR for RUNX1
Survival Probability
p= 0.3953; HR:0.71 (0.32–1.57)
Time from CR (years)

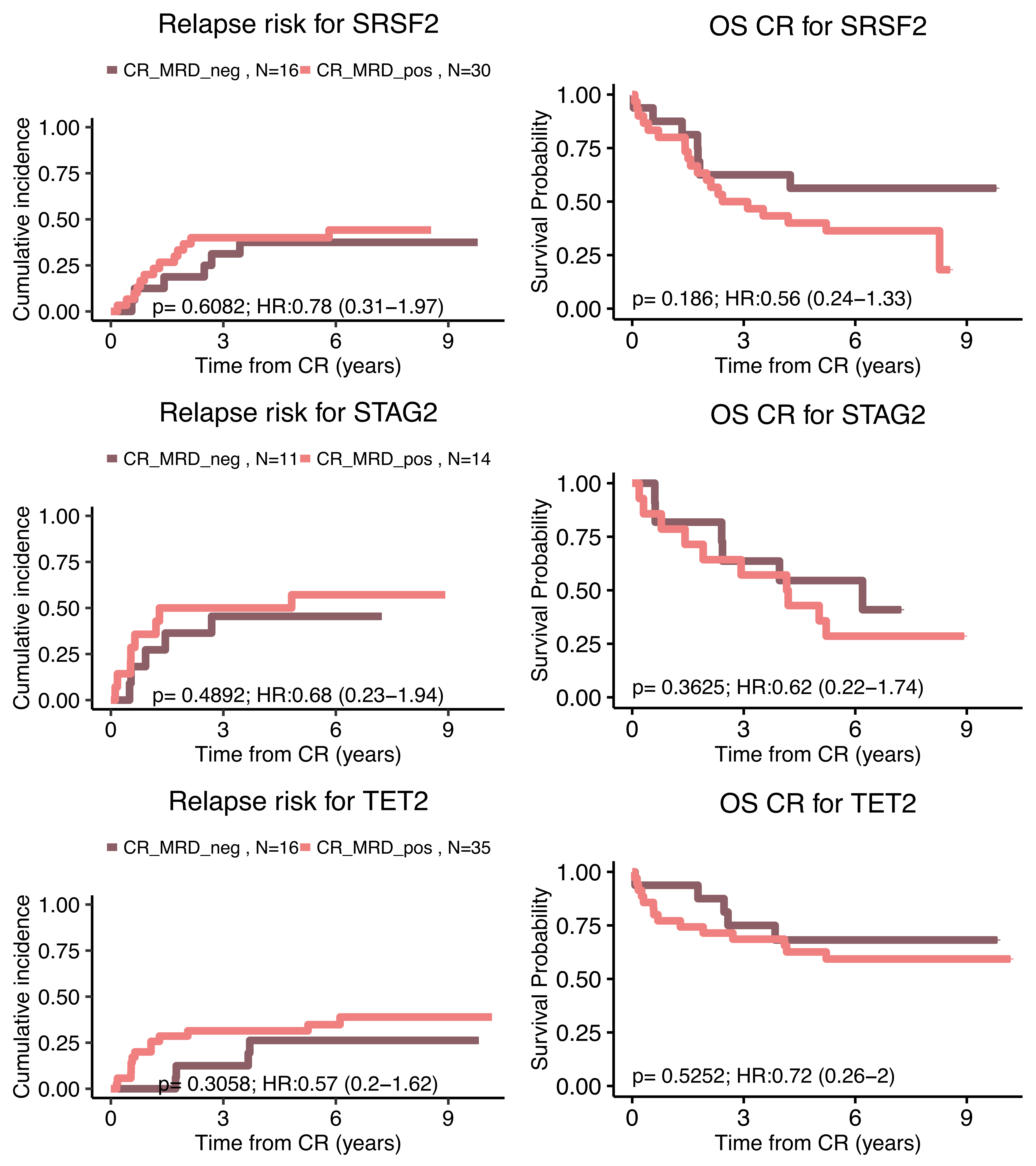

Relapse risk for SRSF2
CR_MRD_neg , N=16
CR_MRD_pos , N=30
Cumulative incidence
1.00
0.75
0.50
0.25
0.00
p= 0.6082; HR:0.78 (0.31−1.97)
0
3
6
9
Time from CR (years)
OS CR for SRSF2
Survival Probability
p= 0.186; HR:0.56 (0.24−1.33)
Relapse risk for STAG2
CR_MRD_neg , N=11
CR_MRD_pos , N=14
p= 0.4892; HR:0.68 (0.23−1.94)
OS CR for STAG2
p= 0.3625; HR:0.62 (0.22−1.74)
Relapse risk for TET2
CR_MRD_neg , N=16
CR_MRD_pos , N=35
p= 0.3058; HR:0.57 (0.2−1.62)
OS CR for TET2
p= 0.5252; HR:0.72 (0.26−2)

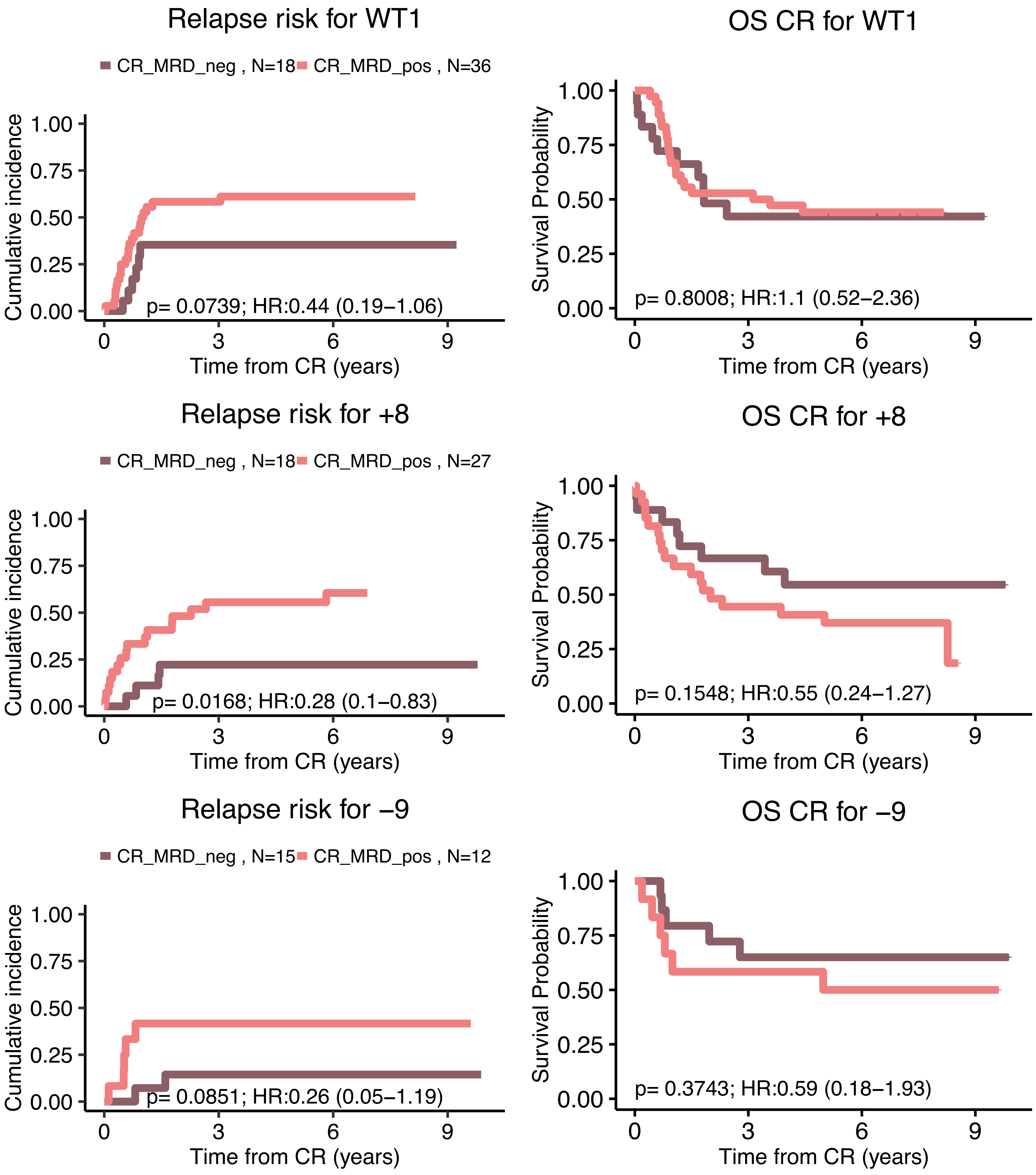

Relapse risk for WT1
CR_MRD_neg , N=18
CR_MRD_pos , N=36
Cumulative incidence
1.00
0.75
0.50
0.25
0.00
p= 0.0739; HR:0.44 (0.19–1.06)
0
3
6
9
Time from CR (years)
OS CR for WT1
Survival Probability
1.00
0.75
0.50
0.25
0.00
p= 0.8008; HR:1.1 (0.52–2.36)
0
3
6
9
Time from CR (years)
Relapse risk for +8
CR_MRD_neg , N=18
CR_MRD_pos , N=27
Cumulative incidence
1.00
0.75
0.50
0.25
0.00
p= 0.0168; HR:0.28 (0.1–0.83)
0
3
6
9
Time from CR (years)
OS CR for +8
Survival Probability
1.00
0.75
0.50
0.25
0.00
p= 0.1548; HR:0.55 (0.24–1.27)
0
3
6
9
Time from CR (years)
Relapse risk for −9
CR_MRD_neg , N=15
CR_MRD_pos , N=12
Cumulative incidence
1.00
0.75
0.50
0.25
0.00
p= 0.0851; HR:0.26 (0.05–1.19)
0
3
6
9
Time from CR (years)
OS CR for −9
Survival Probability
1.00
0.75
0.50
0.25
0.00
p= 0.3743; HR:0.59 (0.18–1.93)
0
3
6
9
Time from CR (years)

**S.Figure 36: Univariate and Multivariate Regression plots for model containing classes, ITD, clinical information as well as minimal residual disease (MRD) status for relapse (panel A) and survival post complete remission (panel B) endpoints in the AML 17 NCRI Trial Cohort with post course 1 MRD analysis (n=523).**

A. The first panel represents a univariate Cox model for all the covariates mentioned above for the relapse endpoint. The second panel represents a multivariate Cox model for all the covariates mentioned above for the relapse endpoint (β coefficients were averaged over 100 iterations).

B. The first panel represents a univariate Cox model for all the covariates mentioned above for the death from complete remission endpoint. The second panel represents a multivariate Cox model for all the covariates mentioned above for the survival post complete remission endpoint (β coefficients were averaged over 100 iterations).

The horizontal dotted curve corresponds to the pvalue threshold of 0.05 and the vertical one correspond to β=0 on the x axis. We highlighted the predictors that have a significant effect (pvalue greater than the threshold :0.05 here). The colors correspond to different types for the covariates: green for classes and ITD, pink for MRD status, red for clinical data.

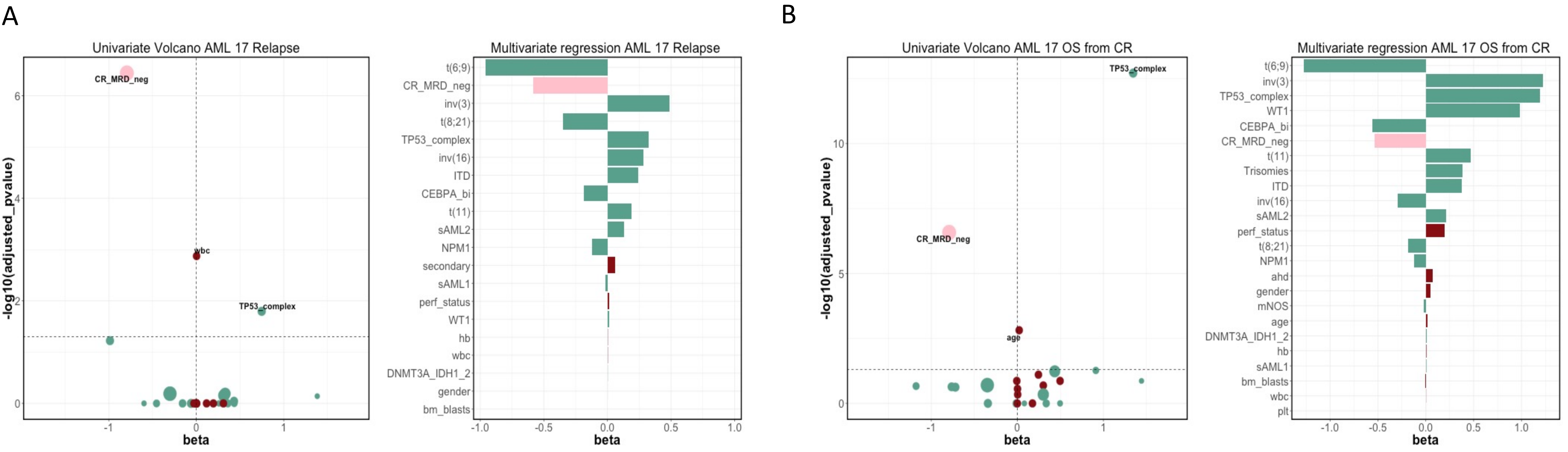

**S.Figure 37: Univariate and Multivariate Regression plots for the classes, ITD stratified by minimal residual disease (MRD) status on Relapse (panel A) and survival post complete remission (panel B) endpoints in the AML 17 NCRI Trial Cohort with post course 1 MRD analysis (n=523).**

A. The first panel represents a univariate Cox model for all the classes stratified by MRD status for the relapse endpoint. The second panel represents a multivariate Cox model for all the covariates mentioned above stratified by MRD status for the relapse endpoint (β coefficients were averaged over 100 iterations). We further stratified NPM1 class based on ITD status.

B. The first panel represents a univariate Cox model for all the classes stratified by MRD for the death from complete remission endpoint. The second panel represents a multivariate Cox model for all the covariates mentioned above stratified by MRD for the death from complete remission endpoint (β coefficients were averaged over 100 iterations). We further stratified NPM1 class based on ITD status.

The horizontal dotted curve corresponds to the pvalue threshold of 0.05 and the vertical one corresponds to β=0 on the x axis. We highlighted the predictors that have a significant effect (pvalue greater than the threshold :0.05 here). The two colors are defined in the legend corresponding to the stratification of each covariate based on MRD status.

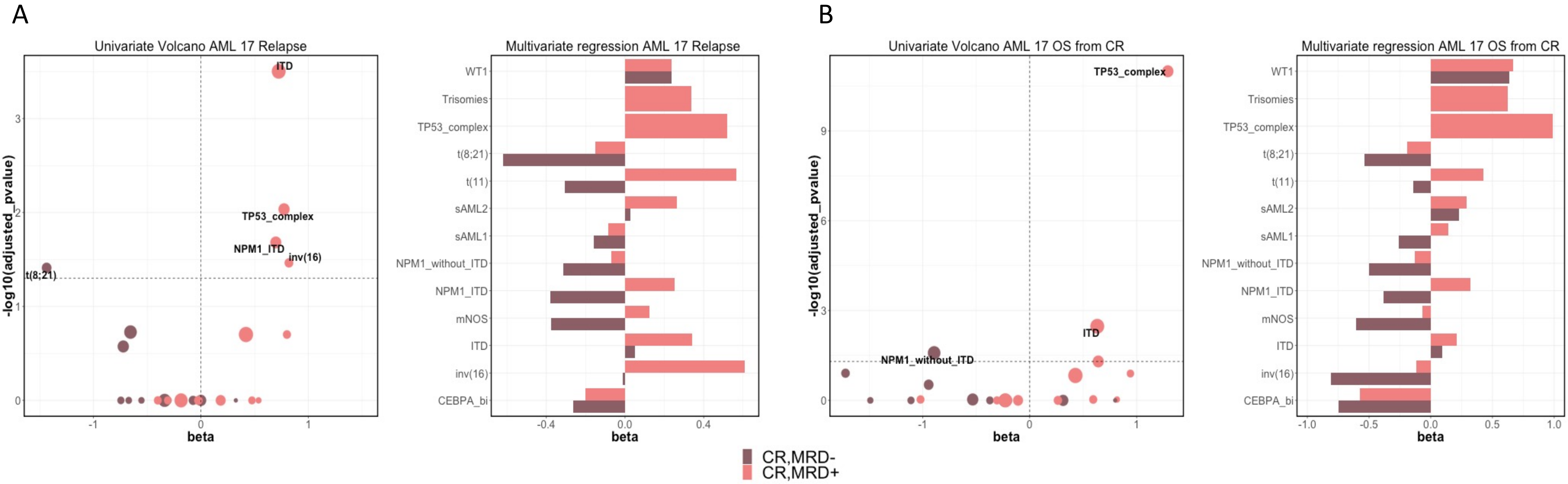

**S.Figure 38: Kaplan-Meier overall survival curves and associated risk tables** comparing each of the proposed risk strata (Favorable$^P$, Intermediate$^P$, Adverse$^P$) by the presence of NPM1 and FLT3$^{ITD}$ status for the Favorable$^P$ and by FLT3$^{ITD}$ status for the Intermediate$^P$ and Adverse$^P$ in the training AML NCRI cohort (n=2,113) and the validation AML SG cohort (n=1,540).

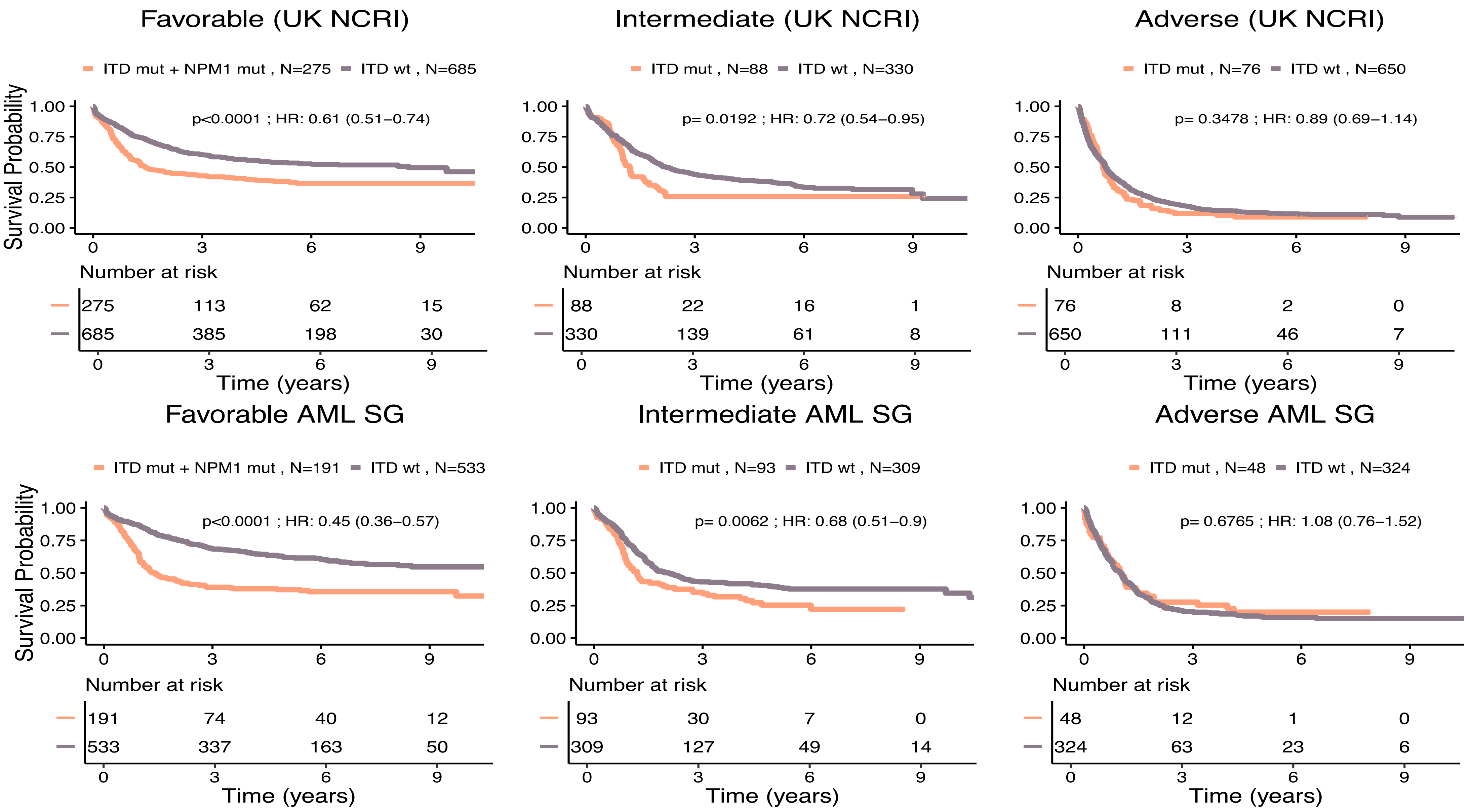

**S.Figure 39: Kaplan-Meier curves for overall survival and associated risk tables comparing ITD clinical ratio for all ITD mutated patients and for the subset of patients with both NPM1 and ITD mutations on the AML NCRI cohort (n=2,113).**

A. Kaplan-Meier curves for overall survival comparing ITD ratio (low is less than 50 and high is more than 50) for ITD mutated patients on the AML NCRI cohort (n=2,113). Dotted curves represent the ELN[2017] risk categories.

B. Kaplan-Meier curves for overall survival comparing ITD ratio (low is less than 50 and high is more than 50) for patients that have both NPM1 and ITD mutations on the AML NCRI cohort (n=2,113). Dotted curves represent the ELN[2017] risk categories.

P values were computed using log-rank test to compare ITD ratio survival differences.

A

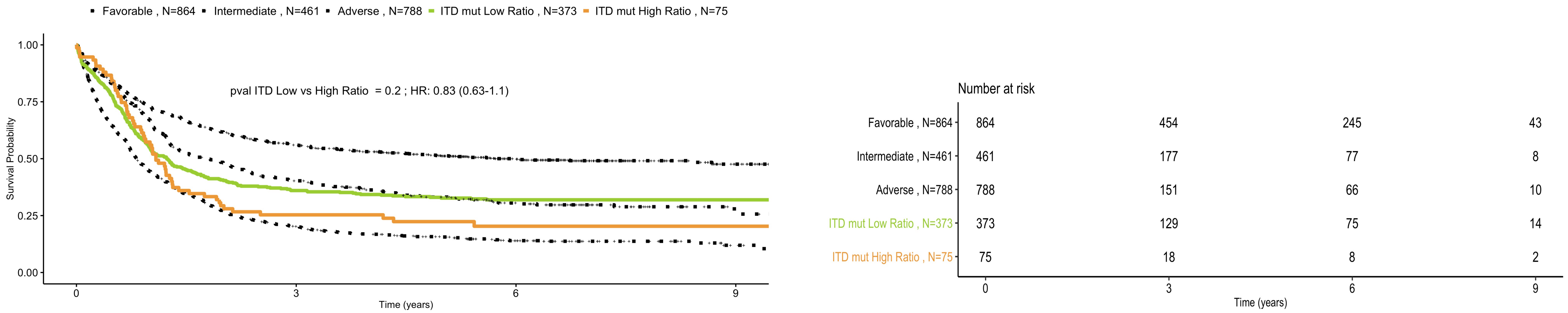

B

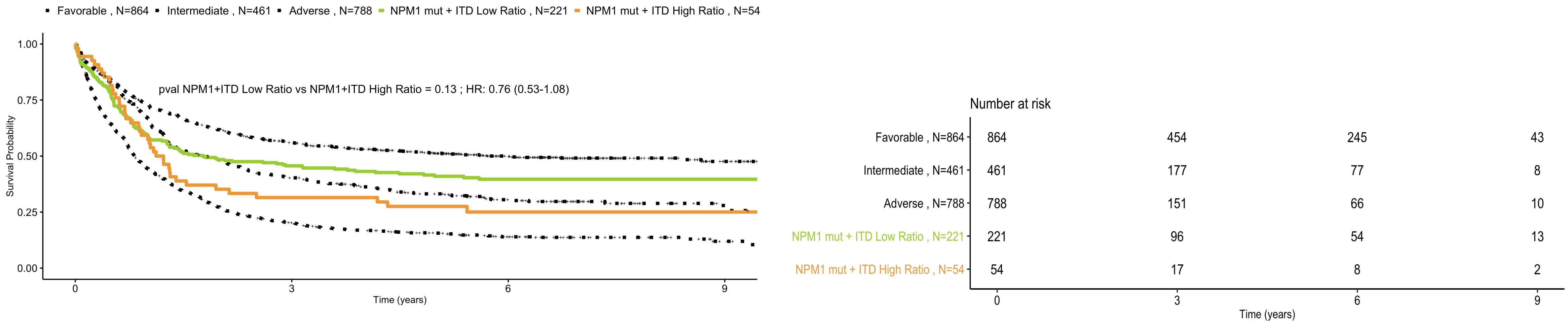

**S. Figure 40: Sankey plot comparing the proportion of the ELN2017 risk groups shifting in the proposed risk groups in the AML NCRI cohort (n=2,113).**

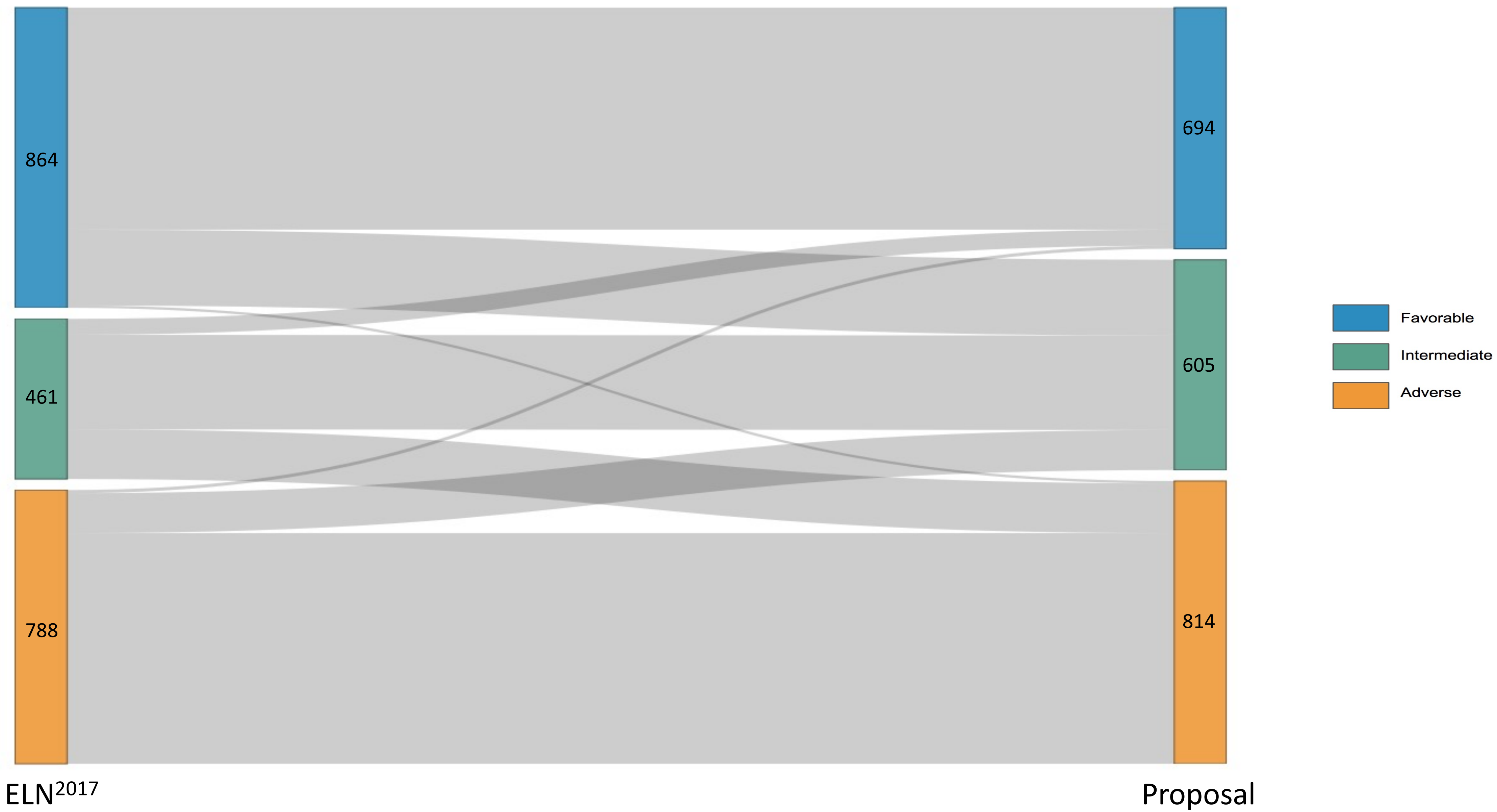

**S. Figure 41: New risk proposal based on the AML classes on the subset of intensively treated patients in the AML NCRI cohort (n=1,755).** A. Kaplan-Meier overall survival curves comparing each of the proposed risk strata ($Favorable^P$, $Intermediate^P$, $Adverse^P$) by $FLT3^{ITD}$ status for the training AML NCRI cohort for intensively treated patients (n=1,755) validate the rationale for the $FLT3^{ITD}$ shift in risk. P values were computed using the log-rank test. B. The estimated improvement in the concordance index (C-index) and pseudo-variance explained ($R^2$ ) for the two classifiers in the training AML NCRI Cohort (n=1,755). 95% confidence intervals were generated by bootstrap resampling for the C-index.

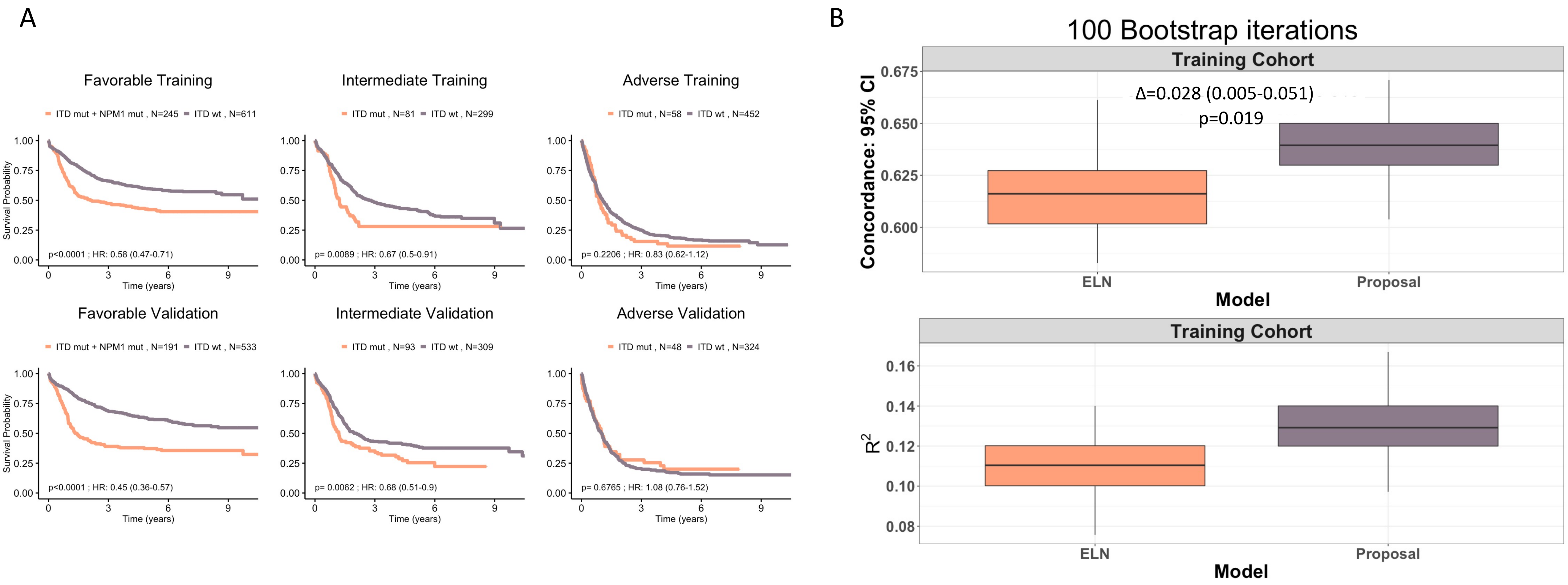

**S. Figure 42: Example presentation of personalized clinical decision support tool for relative contribution of the covariates in all possible transitions.**

The calculator is derived using the multi-state models that consider data from (n=3,201 total patients, UK NCRI and AMLSG) all intensively treated. Input parameters to include cytogenetic, genetic, clinical and demographic are considered to display each patient's $ELN^{2017}$ score alongside the proposed by this study molecular class and proposed risk group. Personalized estimates on the basis of age and gender are also enabled.

To contrast the contribution of the covariates, we also computed the relative contribution for a younger (fictif 40 year old) patient with the same covariates. The relative contributions correspond to the relative $\beta^*(Z-Z_{median})$ values where β is the coefficient derived from the multi-state Cox model for that particular transition for that specific covariate named Z and $Z_{median}$ is that same covariate for a median patient in the combined cohort. For more details, refer to S.Appendix. Precise age is replaced by 5-year range to remove identifying information.

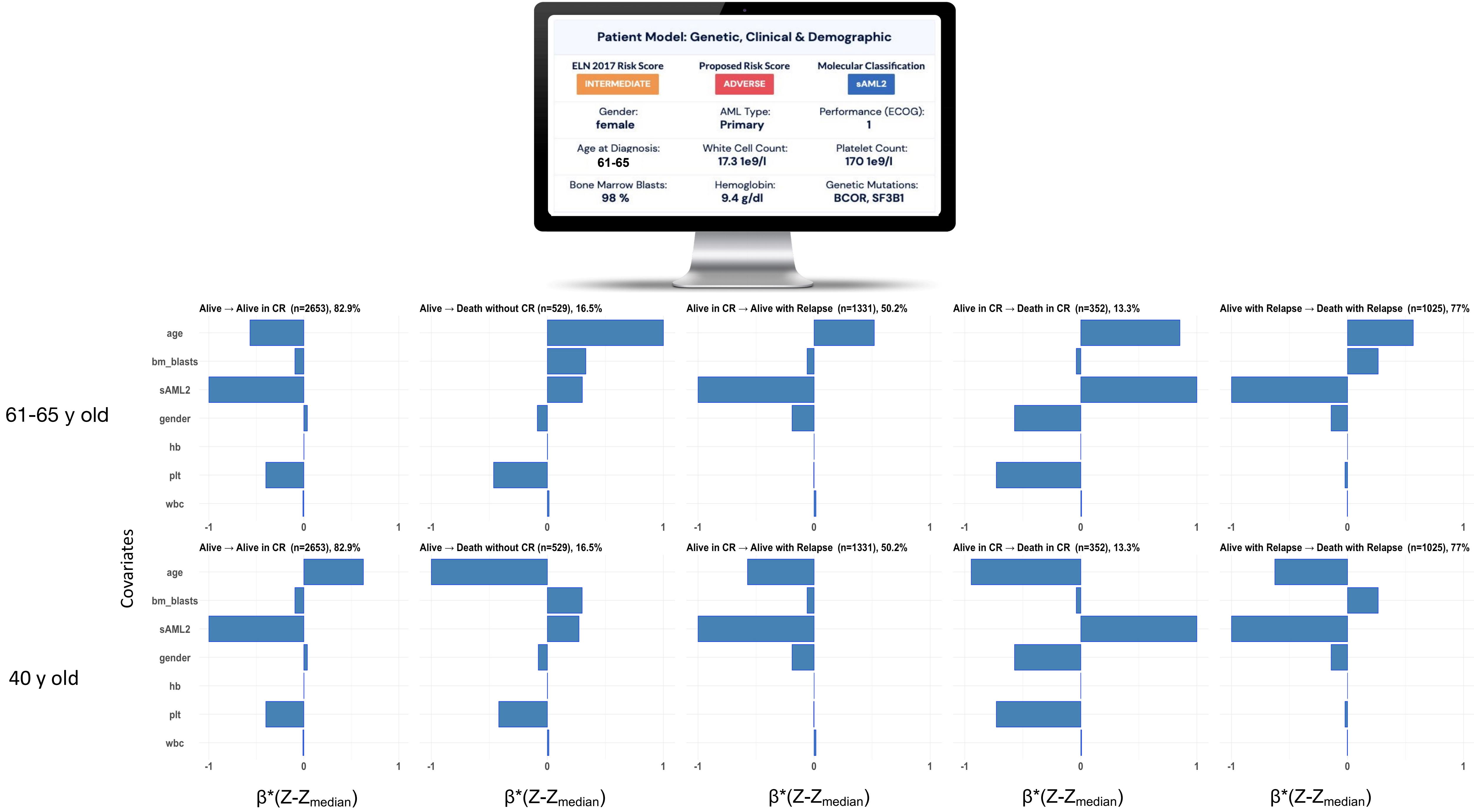
Patient Model: Genetic, Clinical & Demographic
ELN 2017 Risk Score
INTERMEDIATE
Proposed Risk Score
ADVERSE
Molecular Classification
sAML2
Gender:
female
AML Type:
Primary
Performance (ECOG):
1
Age at Diagnosis:
61-65
White Cell Count:
17.3 1e9/l
Platelet Count:
170 1e9/l
Bone Marrow Blasts:
98 %
Hemoglobin:
9.4 g/dl
Genetic Mutations:
BCOR, SF3B1
61-65 y old
40 y old
Covariates
Alive → Alive in CR (n=2653), 82.9%
Alive → Death without CR (n=529), 16.5%
Alive in CR → Alive with Relapse (n=1331), 50.2%
Alive in CR → Death in CR (n=352), 13.3%
Alive with Relapse → Death with Relapse (n=1025), 77%
age
bm_blasts
sAML2
gender
hb
plt
wbc
-1
0
1
β*(Z-Z_median)

**S. Figure 43: Comparison of different statical models C-index distributions on the top performing feature combination.**

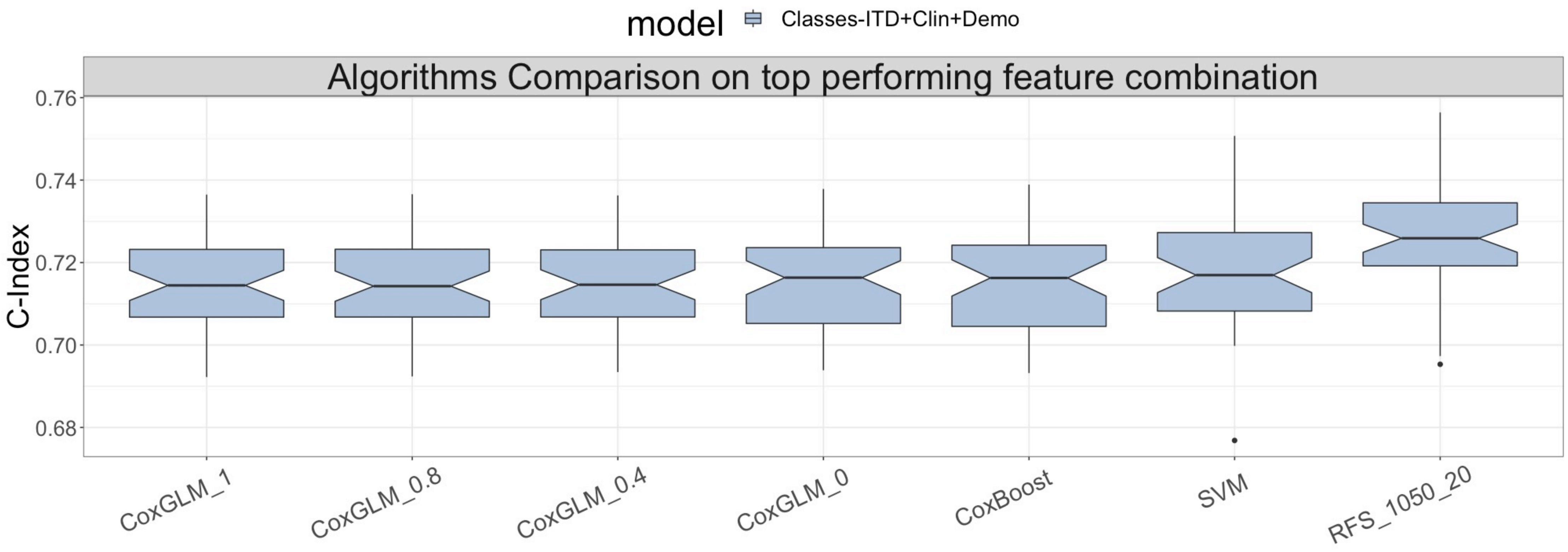

**S. Figure 44: Risk calculator web infrastructure**

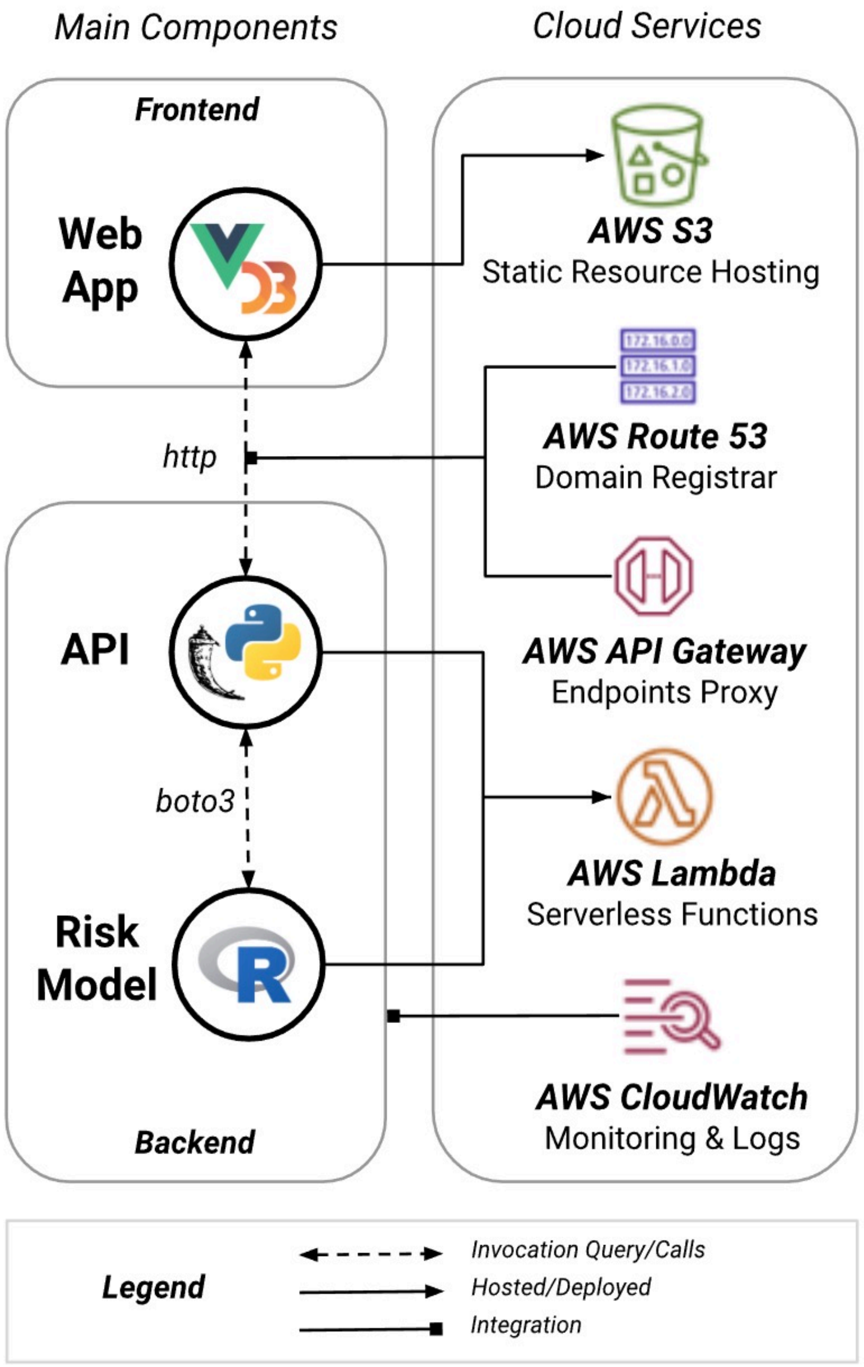

### Supplementary Tables

**S.Table 1: Baseline cohort Characteristics for training (AML NCRI) and validation (AML SG) cohorts**

| S.Table 1a | |
|---|---|
| **Training Cohort** | **AML NCRI** |
| **Variable** | **Distribution in the cohort** |
| **Sample size** | **n** |
| Total | 2113 |
| AML 17 | 1305 |
| AML 16 | 455 |
| Other trials* | 353 |
| **Follow up time** | **Days median (range)** |
| AML 17 | 2290 (36-3897) |
| AML 16 | 1468 (126-2664) |
| Others* | 2368 (625-4091) |
| **Type of AML** | **Total (AML17,AML 16, Other trials*)** |
| primary | 1748 (1144,334,270) |
| secondary | 260 (103,97,60) |
| other | 105 (58,24,23) |
| **Gender** | **Total (AML17,AML 16,others)** |
| Female | 929 (597,186,146) |
| Male | 1184 (708,269,207) |
| **Age** | **median (range)** |
| AML 17 | 52 (16-101) |
| AML 16 | 70 (54-91) |
| Other trials* | 69 (17-91) |
| **ELN 2017 risk group** | **Total (AML17,AML 16,others)** |
| Favorable | 864 (566,162,136) |
| Intermediate | 461 (323,71,67) |
| Adverse | 788 (416,222,150) |
| **WBC Count *(10e9/l)*** | **median (range)** |
| AML 17 | 14 (0.3-456) |
| AML 16 | 20 (0-398) |
| Other trials* | 22 (0.5-432) |
| **Platelet Count *(10e9/l)*** | **median (range)** |
| AML 17 | 60 (2-2013) |

| S.Table 1b | |
|---|---|
| **Validation Cohort** | **AML SG** |
| **Variable** | **Distribution in the cohort** |
| **Sample size** | **n** |
| Total | 1540 |
| HD98A | 627 |
| HD98B | 173 |
| 07-04 | 740 |
| **Follow up time** | **Days median(range)** |
| HD98A | 2693 (30-5384) |
| HD98B | 2709 (1774-5185) |
| 07-04 | 1781 (35-3080) |
| **Type of AML** | **Total (HD98A,HD98B,07-04)** |
| primary | 1376 (572,148,656) |
| secondary | 61 (22,9,30) |
| other | 103 (33,16,54) |
| **Gender** | **Total (HD98A,HD98B,07-04)** |
| Female | 723 (306,76,341) |
| Male | 817(321,97,399) |
| **Age** | **median(range)** |
| HD98A | 47 (18-65) |
| HD98B | 66 (58-84) |
| 07-04 | 49 (18-61) |
| **ELN 2017 risk group** | **Total (HD98A,HD98B,07-04)** |
| Favorable | 618 (291,48,279) |
| Intermediate | 387 (157,50,180) |
| Adverse | 535 (179,75,281) |
| **WBC Count *(10e9/l)*** | **median(range)** |
| HD98A | 14 (0.2-427) |
| HD98B | 8 (0.4-303) |
| 07-04 | 15 (0.2-533) |
| **Platelet Count *(10e9/l)*** | **median(range)** |
| HD98A | 50 (2-746) |

| | | | |
|---|---|---|---|
| AML 16 | 54 (5-706) | HD98B | 64 (6-445) |
| Other trials* | 51 (3-937) | 07-04 | 55 (5-916) |
| **Blasts *(%)*** | **median** | **Blasts *(%)*** | **median** |
| AML 17 | 60 | HD98A | 75 |
| AML 16 | 59 | HD98B | 75 |
| Other trials* | 59 | 07-04 | 75 |
| **Treatment** | **Total (AML17,AML 16,others)** | **Transplant type** | **Total (HD98A,HD98B,07-04)** |
| Intensive | 1755 (1305,289,161) | ALLO-HSCT-MRD | 302 (119,10,173) |
| Non-intensive | 358 (0,166,192) | AUTO | 95 (87,3,5) |
| | | HAPLO | 14 (0,0,14) |
| **Transplant type** | **AML17** | ALLO-HSCT-MUD | 409 (125,2,282) |
| ALLO | 215 | Other types | |
| AUTO | 2 | | |
| HAPLO | 2 | | |
| MUD/UD | 297 | | |
| Other types | 64 | | |

*Other trials include 5 patients from AML11 , 3 from AML12 , 21 from AML14 ,135 from AML15 and 189 from AML Li1

**S.Table 6: Repartition of age in each of the classes in the AML NCRI cohort**

| Classes | n | Median Age | Min Age | Max Age |
|---|---|---|---|---|
| **WT1** | 40 | 41.2 | 16.8 | 80.7 |
| **t(6;9)** | 16 | 41.6 | 16.9 | 68.1 |
| **biCEBPA** | 38 | 43.1 | 19 | 84.4 |
| **t(11)** | 75 | 43.7 | 15.9 | 100.7 |
| **inv(16)** | 92 | 44.3 | 20.6 | 78.5 |
| **t(8;21)** | 100 | 46.4 | 16 | 77.8 |
| **inv(3)** | 17 | 53 | 16.3 | 76.6 |
| **t(15;17)** | 19 | 53.2 | 29.7 | 70.6 |
| **mNOS** | 124 | 54 | 19.3 | 84.4 |
| **Trisomies** | 44 | 55.5 | 17 | 79.9 |
| **NPM1** | 674 | 58.8 | 16.1 | 86.4 |
| **sAML1** | 100 | 59.6 | 15.9 | 83.5 |
| **No events** | 46 | 60.3 | 22.8 | 76.5 |
| **DNMT3A-IDH** | 19 | 61.7 | 41.7 | 83.5 |
| **TP53-Complex** | 208 | 62.5 | 19.5 | 91.1 |
| **sAML2** | 501 | 66.4 | 23.1 | 90.8 |
| **TOTAL** | 2113 | 59.2 | 15.9 | 100.7 |

**S.Table 7: Detailed description of the features considered for each prognostic model in the study.**

| Models | Number of features | Features |
|---|---|---|
| **Model 1 : ELN 2017 Risk strata** | 3 | favorable,intermediate,adverse risk strata |
| **Model 2 : Genes** | 84 | ASXL1 , ASXL2 , ASXL3 , ATRX , BAGE3 , BCOR , BRAF , CBFB , CBL , CDKN2A , CEBPA_bi , CEBPA_mono , CNTN5 , CREBBP , CSF1R , CSF3R , CTCF , CUL2 , CUX1 , DNMT3A , EED , ETV6 , EZH2 , FBXW7 , ITD , FLT3_TKD , FLT3_other , GATA1 , GATA2 , GNAS , GNB1 , IDH1 , IDH2_p.R140 , IDH2_p.R172 , JAK2 , JAK3 , KANSL1 , KDM6A , KIT , KMT2C , KMT2D , KMT2E , KRAS , LUC7L2 , MED12 , MLL , MPL , MYC , NF1 , NFE2 , NOTCH1 , NPM1 , NRAS_other , NRAS_p.G12_13 , NRAS_p.Q61_62 , PDS5B , PHF6 , PPFIA2 , PRPF8 , PTEN , PTPN11 , PTPRF , PTPRT , RAD21 , RIT1 , RUNX1 , S100B , SETBP1 , SF1 , SF3B1 , SMC1A , SMC3 , SMG1 , SPP1 , SRSF2 , STAG2 , STAT5B , SUZ12 , TET2 , TP53 , U2AF1_p.S34 , U2AF1_p.Q157 , WT1 , ZRSR2 |
| **Model 3 : Classes** | 16 | NPM1 , t(11) , TP53_complex , sAML2 , sAML1 , CEBPA_bi , DNMT3A_IDH1_2 , inv(3) , mNOS , t(8;21), no_events , WT1 , inv(16) , Trisomies , t(6;9) , t(15;17) |
| **Model 4 : Classes+ITD** | 17 | NPM1 , t(11) , TP53_complex , sAML2 , sAML1 , CEBPA_bi , DNMT3A_IDH1_2 , inv(3) , mNOS , t(8;21), no_events , WT1 , inv(16) , Trisomies , t(6;9) , t(15;17) , ITD |
| **Model 5 : Genes+Cytogenetics** | 154 | ASXL1 , ASXL2 , ASXL3 , ATRX , BAGE3 , BCOR , BRAF , CBFB , CBL , CDKN2A , CEBPA_bi , CEBPA_mono , CNTN5 , CREBBP , CSF1R , CSF3R , CTCF , CUL2 , CUX1 , DNMT3A , EED , ETV6 , EZH2 , FBXW7 , ITD , FLT3_TKD , FLT3_other , GATA1 , GATA2 , GNAS , GNB1 , IDH1 , IDH2_p.R140 , IDH2_p.R172 , JAK2 , JAK3 , KANSL1 , KDM6A , KIT , KMT2C , KMT2D , KMT2E , KRAS , LUC7L2 , MED12 , MLL , MPL , MYC , NF1 , NFE2 , NOTCH1 , NPM1 , NRAS_other , NRAS_p.G12_13 , NRAS_p.Q61_62 , PDS5B , PHF6 , PPFIA2 , PRPF8 , PTEN , PTPN11 , PTPRF , PTPRT , RAD21 , RIT1 , RUNX1 , S100B , SETBP1 , SF1 , SF3B1 , SMC1A , SMC3 , SMG1 , SPP1 , SRSF2 , STAG2 , STAT5B , SUZ12 , TET2 , TP53 , U2AF1_p.S34 , U2AF1_p.Q157 , WT1 , ZRSR2 ,<br>+8 , +11 , +13 , +21 , +22 , -20 , -3 , -5 , -7 , -9 , -12 , -13 , -16 , -17 , -18 , minusy , t(v;11) , t(10;21) , t(12;13) , t(12;17) , t(12;22) , t(13;19) , t(15;16) , t(15;17) , t(16;17) , t(16;21) , t(17;19) , t(17;21) , t(1;12) , t(1;14) , t(1;16) , t(1;17) , t(1;19) , t(1;3) , t(1;4) , t(1;5) , t(1;6) , t(2;17) , t(2;3) , t(2;5) , t(2;7) , t(2;9) , t(3;16) , t(3;21) , t(3;5) , t(3;7) , t(3;9) , t(4;12) , t(4;21) , t(4;9) , t(5;12) , t(5;17) , t(5;9) , t(6;9) , t(7;16) , t(7;17) , t(7;8) , t(8;10) , t(8;13) , t(8;16) , t(8;17) , t(8;21) , t(9;11) , t(9;13) , t(9;17) , t(9_22) , complex , others_transloc , inv(3) , inv(16) |
| **Model 6 : Clinical+Demographic** | 9 | ahd , perf_status , bm_blasts , secondary , wbc , hb , plt , gender , age |
| **Model 7 : Genes+Cytogenetics+Clinical+Demographic** | 163 | ASXL1 , ASXL2 , ASXL3 , ATRX , BAGE3 , BCOR , BRAF , CBFB , CBL , CDKN2A , CEBPA_bi , CEBPA_mono , CNTN5 , CREBBP , CSF1R , CSF3R , CTCF , CUL2 , CUX1 , DNMT3A , EED , ETV6 , EZH2 , FBXW7 , ITD , FLT3_TKD , FLT3_other , GATA1 , GATA2 , GNAS , GNB1 , IDH1 , IDH2_p.R140 , IDH2_p.R172 , JAK2 , JAK3 , KANSL1 , KDM6A , KIT , KMT2C , KMT2D , KMT2E , KRAS , LUC7L2 , MED12 , MLL , MPL , MYC , NF1 , NFE2 , NOTCH1 , NPM1 , NRAS_other , NRAS_p.G12_13 , NRAS_p.Q61_62 , PDS5B , PHF6 , PPFIA2 , PRPF8 , PTEN , PTPN11 , PTPRF , PTPRT , RAD21 , RIT1 , RUNX1 , S100B , SETBP1 , SF1 , SF3B1 , SMC1A , SMC3 , SMG1 , SPP1 , SRSF2 , STAG2 , STAT5B , SUZ12 , TET2 , TP53 , U2AF1_p.S34 , U2AF1_p.Q157 , WT1 , ZRSR2 ,<br>+8 , +11 , +13 , +21 , +22 , -20 , -3 , -5 , -7 , -9 , -12 , -13 , -16 , -17 , -18 , minusy , t(v;11) , t(10;21) , t(12;13) , t(12;17) , t(12;22) , t(13;19) , t(15;16) , t(15;17) , t(16;17) , t(16;21) , t(17;19) , t(17;21) , t(1;12) , t(1;14) , t(1;16) , t(1;17) , t(1;19) , t(1;3) , t(1;4) , t(1;5) , t(1;6) , t(2;17) , t(2;3) , t(2;5) , t(2;7) , t(2;9) , t(3;16) , t(3;21) , t(3;5) , t(3;7) , t(3;9) , t(4;12) , t(4;21) , t(4;9) , t(5;12) , t(5;17) , t(5;9) , t(6;9) , t(7;16) , t(7;17) , t(7;8) , t(8;10) , t(8;13) , t(8;16) , t(8;17) , t(8;21) , t(9;11) , t(9;13) , t(9;17) , t(9_22) , complex , others_transloc , inv(3) , inv(16) ,<br>ahd , perf_status , bm_blasts , secondary , wbc , hb , plt , gender , age |
| **Model 8 : Classes+ITD+Clinical+Demographic** | 26 | NPM1 , t(11) , TP53_complex , sAML2 , sAML1 , CEBPA_bi , DNMT3A_IDH1_2 , inv(3) , mNOS , t(8;21), no_events , WT1 , inv(16) , Trisomies , t(6;9) , t(15;17) , ITD ,<br>ahd , perf_status , bm_blasts , secondary , wbc , hb , plt , gender , age |

**S.Table 8 : Comparison of coefficients for a Cox Proposal Hazard model and a Cox Proportional Hazard weighted model for the classes, ELN and the risk propoosal**

| | | COX PH | COX PH | COX PH WEIGHTED | COX PH WEIGHTED |
|---|---|---|---|---|---|
| **Model** | **Group** | **Hazard ratio [95%CI]** | **p-val** | **Hazard ratio [95%CI]** | **p-val** |
| **Risk Proposal** | **Favorable** | 0.62 [0.54,0.72] | <0.0001 | 0.63 [0.54,0.75] | <0.0001 |
| | **Adverse** | 2.00 [1.77,2.27] | <0.0001 | 1.98 [1.73,2.26] | <0.0001 |
| | **Intermediate** | Ref | Ref | Ref | Ref |
| **ELN** | **Favorable** | 0.63 [0.55,0.73] | <0.0001 | 0.68 [0.58,0.79] | <0.0001 |
| | **Adverse** | 1.74 [1.52,1.99] | <0.0001 | 1.79 [1.56,2.06] | <0.0001 |
| | **Intermediate** | Ref | Ref | Ref | Ref |
| **Classes** | **NPM1** | 1.29 [0.82,2.05] | 0.27 | 1.36 [0.83,2.21] | 0.22 |
| | **t(11)** | 1.52 [0.89,2.58] | 0.12 | 1.40 [0.80,2.43] | 0.23 |
| | **TP53-complex** | 5.3 [3.31,8.52] | <0.0001 | 5.23 [3.17,8.62] | <0.0001 |
| | **sAML2** | 2.64 [1.67,4.18] | <0.0001 | 2.60 [1.60,4.22] | <0.0001 |
| | **sAML1** | 1.48 [0.89,2.47] | 0.13 | 1.42 [0.83,2.43] | 0.2 |
| | **DNMT3A-IDH** | 2.41 [1.23,4.75] | 0.01 | 2.38 [1.18,4.79] | 0.02 |
| | **inv(3)** | 4.79 [2.49,9.23] | <0.0001 | 4.26 [2.37,7.65] | <0.0001 |
| | **mNOS** | 1.38 [0.84,2.27] | 0.21 | 1.38 [0.81,2.36] | 0.23 |
| | **t(8;21)** | 0.58 [0.33,1.02] | 0.06 | 0.54 [0.30,0.97] | 0.04 |
| | **no events** | 1.06 [0.59,1.91] | 0.85 | 0.89 [0.49,1.61] | 0.7 |
| | **WT1** | 2.32 [1.31,4.10] | <0.01 | 2.26 [1.24,4.11] | <0.01 |
| | **inv(16)** | 0.51 [0.29,0.92] | 0.03 | 0.45 [0.24,0.82] | <0.01 |
| | **Trisomies** | 1.46 [0.81,2.63] | 0.21 | 1.60 [0.85,3.02] | 0.14 |
| | **t(6;9)** | 1.87 [0.92,3.79] | 0.08 | 1.55 [0.84,2.88] | 0.16 |
| | **t(15;17)** | 0.35 [0.12,1.04] | 0.06 | 0.45 [0.13,1.50] | 0.19 |
| | **BiCEBPA** | Ref | Ref | Ref | Ref |

**S.Table 9: Multivariate Cox Transitions for both the training and validation cohorts**

#### Alive --> Alive in CR

| | AML MRC COHORT | | | | | | AML SG COHORT | | | | |
|---|---|---|---|---|---|---|---|---|---|---|---|
| | coef | exp(coef) | se(coef) | z | Pr(>\|z\|) | | coef | exp(coef) | se(coef) | z | Pr(>\|z\|) |
| NPM1 | 0.363545109 | 1.4384197 | 0.172951 | 2.10201251 | 3.56E-02 | | 0.331189826 | 1.3926241 | 0.1534609 | 2.15813792 | 0.0309171105 |
| t_11 | 0.149663529 | 1.1614434 | 0.2096688 | 0.71380928 | 4.75E-01 | | 0.009360986 | 1.0094049 | 0.2142866 | 0.04368442 | 0.9651559607 |
| TP53_complex | -0.806529696 | 0.4464045 | 0.2000283 | -4.03207698 | 5.53E-05 | | -0.553684213 | 0.5748281 | 0.1770615 | -3.12707361 | 0.0017655571 |
| sAML2 | -0.386232288 | 0.6796126 | 0.1787007 | -2.16133589 | 3.07E-02 | | -0.397600828 | 0.6719302 | 0.1673337 | -2.37609513 | 0.0174969514 |
| sAML1 | -0.037858398 | 0.9628493 | 0.2061717 | -0.18362556 | 8.54E-01 | | -0.32638424 | 0.7215279 | 0.1779433 | -1.83420325 | 0.0666238291 |
| CEBPA_bi | 0.556600269 | 1.7447308 | 0.2455286 | 2.26694715 | 2.34E-02 | | 0.657259686 | 1.9294977 | 0.1901869 | 3.45586239 | 0.0005485353 |
| DNMT3A_IDH1_[illegible] | -0.734598944 | 0.4796978 | 0.3446163 | -2.13164307 | 3.30E-02 | | -0.676559013 | 0.5083633 | 0.3230397 | -2.09435272 | 0.0362285713 |
| inv_3 | -1.083792333 | 0.3383101 | 0.4133067 | -2.62224712 | 8.74E-03 | | -1.39183155 | 0.2486195 | 0.3635661 | -3.82827693 | 0.0001290435 |
| mNOS | 0.011822345 | 1.0118925 | 0.1966622 | 0.06011499 | 9.52E-01 | | -0.325350421 | 0.7222742 | 0.1707892 | -1.90498266 | 0.0567823252 |
| t_8_21 | 0.356638014 | 1.4285187 | 0.1989563 | 1.79254438 | 7.30E-02 | | 0.549301303 | 1.7320424 | 0.1962405 | 2.79912358 | 0.0051241523 |
| WT1 | -0.338407255 | 0.7129049 | 0.260419 | -1.29947219 | 1.94E-01 | | -0.089552894 | 0.9143399 | 0.2663897 | -0.33617254 | 0.7367407566 |
| inv_16 | 0.684454216 | 1.9826894 | 0.2017593 | 3.39242931 | 6.93E-04 | | 0.674920524 | 1.9638769 | 0.1842009 | 3.66404618 | 0.000248262 |
| Trisomies | -0.008123888 | 0.991909 | 0.2391738 | -0.03396646 | 9.73E-01 | | 0.022237639 | 1.0224867 | 0.226323 | 0.09825618 | 0.9217288706 |
| t_6_9 | -0.030794186 | 0.9696751 | 0.3236556 | -0.09514493 | 9.24E-01 | | -0.393254297 | 0.6748571 | 0.3229184 | -1.21781337 | 0.2232949016 |
| t_15_17 | 0.242157103 | 1.2739943 | 0.3008008 | 0.80504136 | 4.21E-01 | | 0.683676082 | 1.9811472 | 0.1960861 | 3.48661153 | 0.0004891814 |
| no_events | NA | NA | 0 | NA | NA | | NA | NA | 0 | NA | NA |

#### Alive --> Death without CR

| | coef | exp(coef) | se(coef) | z | Pr(>\|z\|) | | coef | exp(coef) | se(coef) | z | Pr(>\|z\|) |
|---|---|---|---|---|---|---|---|---|---|---|---|
| NPM1 | 0.546124648 | 1.73E+00 | 0.468866 | 1.164777554 | 2.44E-01 | | 0.77270376 | 2.1656136 | 0.4106361 | 1.88172393 | 0.0598735095 |
| t_11 | 0.169770683 | 1.19E+00 | 0.5863516 | 0.289537318 | 7.72E-01 | | 0.42737398 | 1.5332259 | 0.5374543 | 0.79518193 | 0.4265076789 |
| TP53_complex | 1.183007838 | 3.26E+00 | 0.466357 | 2.536700214 | 1.12E-02 | | 1.48945895 | 4.4346955 | 0.4028636 | 3.69717936 | 0.0002180083 |
| sAML2 | 0.458136807 | 1.58E+00 | 0.4621503 | 0.991315685 | 3.22E-01 | | 0.40812708 | 1.5039983 | 0.4104064 | 0.99444615 | 0.3200057086 |
| sAML1 | 0.023849534 | 1.02E+00 | 0.5713274 | 0.041744077 | 9.67E-01 | | 0.41786731 | 1.5187191 | 0.4299467 | 0.97190501 | 0.3310978085 |
| CEBPA_bi | -14.62110419 | 4.47E-07 | 1291.145715 | -0.011324132 | 9.91E-01 | | -1.1148288 | 0.3279714 | 1.0723907 | -1.03957331 | 0.2985381827 |
| DNMT3A_IDH1_[illegible] | 0.301140336 | 1.35E+00 | 0.6073227 | 0.495848975 | 6.20E-01 | | 0.3142622 | 1.3692487 | 0.5877944 | 0.53464648 | 0.5928943377 |
| inv_3 | 1.045810681 | 2.85E+00 | 0.5723346 | 1.827271446 | 6.77E-02 | | 1.07199093 | 2.9211896 | 0.4720027 | 2.27115425 | 0.0231376416 |
| mNOS | 0.17942183 | 1.20E+00 | 0.5216425 | 0.343955554 | 7.31E-01 | | 0.10296784 | 1.1084558 | 0.4303067 | 0.23928944 | 0.8108811552 |
| t_8_21 | -0.53800339 | 5.84E-01 | 0.7323256 | -0.734650559 | 4.63E-01 | | 0.28078728 | 1.3241719 | 0.6317464 | 0.44446202 | 0.6567085845 |
| WT1 | 0.403400399 | 1.50E+00 | 0.6061774 | 0.665482404 | 5.06E-01 | | -0.66193516 | 0.5158521 | 1.0717344 | -0.61762983 | 0.5368193794 |
| inv_16 | 0.214438814 | 1.24E+00 | 0.6767911 | 0.316846393 | 7.51E-01 | | -0.05491071 | 0.9465697 | 0.6968227 | -0.07880156 | 0.9371904644 |
| Trisomies | 0.001408581 | 1.00E+00 | 0.6711855 | 0.002098647 | 9.98E-01 | | -0.41735251 | 0.6587887 | 0.8040158 | -0.51908498 | 0.6037014795 |
| t_6_9 | -0.814589862 | 4.43E-01 | 1.0960727 | -0.74318965 | 4.57E-01 | | 0.50632185 | 1.6591772 | 0.6931213 | 0.73049528 | 0.465087495 |
| t_15_17 | 1.246506093 | 3.48E+00 | 0.7342145 | 1.697741091 | 8.96E-02 | | 1.12317261 | 3.0745932 | 0.5441841 | 2.06395701 | 0.0390217925 |
| no_events | NA | NA | 0 | NA | NA | | NA | NA | 0 | NA | NA |

#### Alive in CR --> Alive with Relapse

| | coef | exp(coef) | se(coef) | z | Pr(>\|z\|) | | coef | exp(coef) | se(coef) | z | Pr(>\|z\|) |
|---|---|---|---|---|---|---|---|---|---|---|---|
| **NPM1** | -0.001408045 | 9.99E-01 | 0.2382362 | -0.005910291 | 9.95E-01 | | -0.1574659 | 0.8543059 | 0.2072815 | -0.7596719 | 0.4474507349 |
| **t_11** | 0.36317439 | 1.44E+00 | 0.2836302 | 1.280450552 | 2.00E-01 | | 0.6984768 | 2.0106876 | 0.2655002 | 2.6307956 | 0.0085185249 |
| **TP53_complex** | 0.952378488 | 2.59E+00 | 0.2677306 | 3.557226849 | 3.75E-04 | | 0.8064463 | 2.2399337 | 0.2298895 | 3.5079742 | 0.0004515329 |
| **sAML2** | 0.466167592 | 1.59E+00 | 0.2434997 | 1.914448086 | 5.56E-02 | | 0.3781954 | 1.4596482 | 0.2230743 | 1.6953788 | 0.090003583 |
| **sAML1** | 0.078586857 | 1.08E+00 | 0.2835985 | 0.277106043 | 7.82E-01 | | 0.1790979 | 1.1961378 | 0.239758 | 0.7469943 | 0.4550669915 |
| **CEBPA_bi** | -0.131989723 | 8.76E-01 | 0.3687845 | -0.357904744 | 7.20E-01 | | -0.372125 | 0.6892681 | 0.2775907 | -1.3405528 | 0.1800656832 |
| **NMT3A_IDH1_** | 0.220586679 | 1.25E+00 | 0.5026762 | 0.438824562 | 6.61E-01 | | -0.2321071 | 0.7928612 | 0.4869224 | -0.476682 | 0.6335885653 |
| **inv_3** | 1.339646629 | 3.82E+00 | 0.5514207 | 2.429445815 | 1.51E-02 | | 0.2954368 | 1.3437132 | 0.5361465 | 0.5510375 | 0.5816079897 |
| **mNOS** | 0.172107493 | 1.19E+00 | 0.2695309 | 0.638544488 | 5.23E-01 | | 0.0848482 | 1.0885518 | 0.2336573 | 0.363131 | 0.7165070574 |
| **t_8_21** | -0.830101537 | 4.36E-01 | 0.3100591 | -2.677236487 | 7.42E-03 | | -0.1492062 | 0.8613915 | 0.2748116 | -0.54294 | 0.5871710965 |
| **WT1** | 0.760047438 | 2.14E+00 | 0.3393984 | 2.239395979 | 2.51E-02 | | 0.543008 | 1.7211764 | 0.3376517 | 1.6081895 | 0.1077936799 |
| **inv_16** | 0.224198908 | 1.25E+00 | 0.2719277 | 0.824479784 | 4.10E-01 | | -0.3613845 | 0.6967111 | 0.2632792 | -1.3726284 | 0.1698679061 |
| **Trisomies** | -0.066307115 | 9.36E-01 | 0.3454097 | -0.191966542 | 8.48E-01 | | -0.1926607 | 0.8247618 | 0.3376038 | -0.5706711 | 0.5682226306 |
| **t_6_9** | 0.484166608 | 1.62E+00 | 0.4215767 | 1.148466115 | 2.51E-01 | | 0.7112296 | 2.0364939 | 0.4028591 | 1.765455 | 0.0774873206 |
| **t_15_17** | -1.324262083 | 2.66E-01 | 0.6213259 | -2.131348438 | 3.31E-02 | | -1.3819052 | 0.2510997 | 0.370351 | -3.7313387 | 0.0001904649 |
| **no_events** | NA | NA | 0 | NA | NA | | NA | NA | 0 | NA | NA |

#### Alive in CR --> Death in CR

| | coef | exp(coef) | se(coef) | z | Pr(>\|z\|) | | coef | exp(coef) | se(coef) | z | Pr(>\|z\|) |
|---|---|---|---|---|---|---|---|---|---|---|---|
| **NPM1** | 0.3725916 | 1.45E+00 | 0.5922308 | 0.62913242 | 5.29E-01 | | 0.1492311 | 1.1609413 | 0.4719056 | 0.31623089 | 0.751827267 |
| **t_11** | 0.9494903 | 2.58E+00 | 0.6584668 | 1.44197143 | 1.49E-01 | | 0.3479696 | 1.4161892 | 0.6714226 | 0.51825724 | 0.6042788 |
| **TP53_complex** | 2.0003495 | 7.39E+00 | 0.6149704 | 3.25275768 | 1.14E-03 | | 1.1326851 | 3.1039798 | 0.5097624 | 2.22198626 | 0.026284234 |
| **sAML2** | 0.95531 | 2.60E+00 | 0.5999773 | 1.59224349 | 1.11E-01 | | 1.1212662 | 3.0687374 | 0.483499 | 2.31906626 | 0.020391444 |
| **sAML1** | 0.466007 | 1.59E+00 | 0.6771023 | 0.68823719 | 4.91E-01 | | 0.7680416 | 2.1555408 | 0.5125149 | 1.49857425 | 0.133984116 |
| **CEBPA_bi** | 0.9362796 | 2.55E+00 | 0.7072092 | 1.32390755 | 1.86E-01 | | -0.1436245 | 0.8662129 | 0.6056226 | -0.23715184 | 0.812538997 |
| **NMT3A_IDH1_** | 1.1328158 | 3.10E+00 | 0.9130806 | 1.24065262 | 2.15E-01 | | -0.1067238 | 0.8987739 | 1.0955799 | -0.09741306 | 0.922398373 |
| **inv_3** | 2.9362387 | 1.88E+01 | 0.8212366 | 3.57538692 | 3.50E-04 | | 2.1473937 | 8.5625125 | 0.6722018 | 3.19456707 | 0.001400407 |
| **mNOS** | 0.7235722 | 2.06E+00 | 0.6362799 | 1.13719166 | 2.55E-01 | | 0.7338179 | 2.0830182 | 0.5001634 | 1.46715622 | 0.142333564 |
| **t_8_21** | -0.5734861 | 5.64E-01 | 0.7304544 | -0.78510874 | 4.32E-01 | | -0.1311045 | 0.8771261 | 0.6325719 | -0.20725635 | 0.835809661 |
| **WT1** | 1.1776352 | 3.25E+00 | 0.7639177 | 1.54157348 | 1.23E-01 | | 0.3047887 | 1.3563384 | 0.8368064 | 0.36422852 | 0.715687373 |
| **inv_16** | -1.7810675 | 1.68E-01 | 1.1547802 | -1.54234329 | 1.23E-01 | | -1.0796545 | 0.3397129 | 0.7316333 | -1.47567713 | 0.140030587 |
| **Trisomies** | 0.8609126 | 2.37E+00 | 0.707247 | 1.21727297 | 2.24E-01 | | 0.5495169 | 1.7324159 | 0.6325317 | 0.86875797 | 0.384979527 |
| **t_6_9** | 1.4298654 | 4.18E+00 | 0.8169583 | 1.75023055 | 8.01E-02 | | 1.0083249 | 2.7410056 | 0.8374157 | 1.20409112 | 0.228554361 |
| **t_15_17** | -15.1208762 | 2.71E-07 | 1370.214521 | -0.01103541 | 9.91E-01 | | -0.9642097 | 0.3812844 | 0.7306764 | -1.3196125 | 0.186964426 |
| **no_events** | NA | NA | 0 | NA | NA | | NA | NA | 0 | NA | NA |

#### Alive with Relapse --> Death with Relapse

| | coef | exp(coef) | se(coef) | z | Pr(>\|z\|) | | coef | exp(coef) | se(coef) | z | Pr(>\|z\|) |
|---|---|---|---|---|---|---|---|---|---|---|---|

| | | | | | | | | | | | |
|---|---|---|---|---|---|---|---|---|---|---|---|
| **NPM1** | 0.107627858 | 1.11E+00 | 0.2699735 | 0.398660851 | 6.90E-01 | | 0.140900895 | 1.1513105 | 0.2472142 | 0.56995475 | 0.568708389 |
| **t_11** | -0.001147971 | 9.99E-01 | 0.3233331 | -0.003550428 | 9.97E-01 | | 0.268198571 | 1.3076068 | 0.3066257 | 0.87467754 | 0.3817493825 |
| **TP53_complex** | 0.888020108 | 2.43E+00 | 0.29854 | 2.974542986 | 2.93E-03 | | 0.831365693 | 2.2964528 | 0.2690733 | 3.08973698 | 0.0020033382 |
| **sAML2** | 0.384437046 | 1.47E+00 | 0.2735949 | 1.405132316 | 1.60E-01 | | -0.003961932 | 0.9960459 | 0.2633272 | -0.01504566 | 0.9879957513 |
| **sAML1** | 0.105894454 | 1.11E+00 | 0.3167013 | 0.334366952 | 7.38E-01 | | 0.056839332 | 1.0584857 | 0.282308 | 0.20133798 | 0.840434308 |
| **CEBPA_bi** | -0.777441423 | 4.60E-01 | 0.4585837 | -1.695309653 | 9.00E-02 | | -1.093096424 | 0.335177 | 0.3699773 | -2.95449571 | 0.0031318051 |
| **NMT3A_IDH1_** | 0.885602773 | 2.42E+00 | 0.5192873 | 1.705419513 | 8.81E-02 | | -0.173941868 | 0.8403457 | 0.6227737 | -0.27930189 | 0.7800131598 |
| **inv_3** | 0.208538178 | 1.23E+00 | 0.5667685 | 0.367942425 | 7.13E-01 | | -0.131846516 | 0.8764755 | 0.6231251 | -0.21158916 | 0.8324275645 |
| **mNOS** | -0.213079589 | 8.08E-01 | 0.3097441 | -0.687921337 | 4.92E-01 | | -0.137536451 | 0.8715026 | 0.2762689 | -0.49783548 | 0.618600004 |
| **t_8_21** | 0.069147387 | 1.07E+00 | 0.3546821 | 0.194955938 | 8.45E-01 | | -0.174038292 | 0.8402647 | 0.3217107 | -0.54097769 | 0.5885229604 |
| **WT1** | 0.312308383 | 1.37E+00 | 0.3843168 | 0.812632615 | 4.16E-01 | | 0.20565609 | 1.2283307 | 0.3954188 | 0.52009687 | 0.6029960608 |
| **inv_16** | -1.276652582 | 2.79E-01 | 0.3504552 | -3.642841045 | 2.70E-04 | | -1.197986775 | 0.3018012 | 0.3604135 | -3.32392294 | 0.0008876072 |
| **Trisomies** | 0.141800796 | 1.15E+00 | 0.3893019 | 0.364243824 | 7.16E-01 | | 0.276752562 | 1.31884 | 0.3937008 | 0.70295148 | 0.4820859868 |
| **t_6_9** | 0.471263931 | 1.6020178 | 0.4390568 | 1.073355324 | 0.2831117214 | | -0.340032609 | 0.7117471 | 0.469871 | -0.72367232 | 0.4692669278 |
| **t_15_17** | -1.11841423 | 0.3267976 | 1.0333004 | -1.082370878 | 0.279087763 | | -0.460998285 | 0.6306538 | 0.4426269 | -1.04150525 | 0.2976411173 |
| **no_events** | NA | NA | 0 | NA | NA | | NA | NA | 0 | NA | NA |

| S.Table 10: Cohort characteristics for merged AML NCRI and AMLSG subsets that were used for HSCT analysis | |
|---|---|
| **Variable** | **Distribution in the cohort** |
| **Sample Size** | **n** |
| Total | 2244 |
| AML NCRI | 1095 |
| AML SG | 1149 |
| **Follow up time** | **Days median (range)** |
| Total | 2213 (35-5384) |
| AML NCRI | 2291 (36-3897) |
| AML SG | 2217 (35-5384) |
| **Type of AML (Total, AML NCR, AML SG)** | **Total** |
| primary | 2034, 985, 1049 |
| secondary | 98, 69, 29 |
| others | 112, 41, 71 |
| **Gender (Total, AML NCRI, AML SG)** | **Total** |
| Female | 1080, 514, 566 |
| Male | 1164, 581, 583 |
| **Age** | **median (range)** |
| Total | 50 (16-101) |
| AML NCRI | 51 (16-101) |
| AML SG | 49 (18-80) |
| **ELN 2017 risk group (Total, AML NCR, AML SG)** | **Total** |
| Favorable | 1070, 521, 549 |
| Intermediate | 556, 273, 283 |
| Adverse | 618, 301, 317 |
| ***WBC Count (10e9/l)*** | **median (range)** |

| | |
|---|---|
| Total | 14 (0.2-456) |
| AML NCRI | 14 (0.3-456) |
| AML SG | 14 (0.2-303) |
| ***Platelet Count (10e9/l)*** | **median (range)** |
| Total | 56 (2-2013) |
| AML NCRI | 60 (2-2013) |
| AML SG | 53 (5-746) |
| **Blasts *(%)*** | **median** |
| Total | 70 |
| AML NCRI | 60 |
| AML SG | 75 |

**S.Table 11: Detailed state by state distribution of transplanted patients that achieved complete remission in combined training and validation cohort (n=2,244)**

| | to | | | | | | | |
|---|---|---|---|---|---|---|---|---|
| from | CR1 | TPL in CR1 | TPL in CR2 | Death no TPL | Death from TPL in CR1 | Death from TPL in CR2 | no event | Total |
| CR1 | | n=759 | n=436 | n=507 | | | n=542 | n=2244 |
| TPL in CR1 | | | | | n=347 | | n=412 | n=759 |
| TPL in CR2 | | | | | | n=236 | n=200 | n=436 |
| Death no TPL | | | | | | | n=507 | n=507 |
| Death from TPL in CR1 | | | | | | | n=347 | n=347 |
| Death from TPL in CR2 | | | | | | | n=236 | n=236 |

S.Table 12: HSCT characteristics for AML NCRI and AMLSG subsets by ELN 2017 and proposed risk score

| | AML NCRI cohort N=1,095 ( 568 \| 321 \| 206 ) | AML SG cohort N=1,149 ( 481 \| 438 \| 230 ) |
|---|---|---|
| | **Total ( No transplant \| Transplant in CR1 \| Transplant in CR2 )** | **Total ( No transplant \| Transplant in CR1 \| Transplant in CR2 )** |
| **ELN Favorable** | 521 ( 326 \| 80 \| 115 ) | 549 ( 288 \| 149 \| 112 ) |
| **ELN Intermediate** | 273 ( 109 \| 113 \| 51 ) | 283 ( 97 \| 121 \| 65 ) |
| **ELN Adverse** | 301 ( 133 \| 128 \| 40 ) | 317 ( 96 \| 168 \| 53 ) |
| **Proposal Favorable** | 441 ( 279 \| 66 \| 96 ) | 517 ( 258 \| 142 \| 117 ) |
| **Proposal Intermediate** | 344 ( 147 \| 130 \| 67 ) | 375 ( 139 \| 163 \| 73 ) |
| **Proposal Adverse** | 310 ( 142 \| 125 \| 43 ) | 257 ( 84 \| 133 \| 40 ) |

**S.Table 13: Panel of 32 genes sufficient to deliver complete classification and risk stratification**

| Class defining genes | Independent genes |
|---|---|
| NPM1 | KIT |
| TP53 | KRAS |
| WT1 | NRAS |
| CEBPA | PTPN11 |
| DNMT3A | GATA2 |
| IDH1 | MYC |
| IDH2 | MPL |
| ZRSR2 | SMC3 |
| U2AF1 | **Total ,N=8** |
| SRSF2 | |
| SF3B1 | |
| ASXL1 | |
| STAG2 | |
| BCOR | |
| RUNX1 | |
| EZH2 | |
| MLL | |
| PHF6 | |
| SF1 | |
| NF1 | |
| CUX1 | |
| SETBP1 | |
| FLT3 | |
| TET2 | |
| **Total ,N=24** | |

**S.Table 14: Hyperparameter selection for BDP processes in study**

| Hyperparameter names | Hyperparameter search range | BDP1 | BDP2 |
|---|---|---|---|
| Burnin iterations | 3000 to 7000 | 7000 | 5000 |
| Number of chains | 3 to 7 | 3 | 3 |
| Number of iterations between collected samples | 20 | 20 | 20 |
| Cosine similarity threshold | 0.9 | 0.9 | 0.9 |
| Number of posterior samples | 150-1000 | 250 | 150 |
| Initial number of clusters | 3 to 30 | 17 | 5 |
| Base distribution | Uniform dirichlet, Gaussian, binomial | Gaussian | Gaussian |
| Concentration parameter $\alpha$A | 0.1 to 100 | 0.5 | 2 |
| Concentration parameter $\alpha$B | 0.1 to 100 | 1.5 | 6 |

**S.Table 15: Software and packages used for the regression modeling**

| Algorithms | Language / Library |
|---|---|
| Cox Proportional Hazards | R / survival<br>Python / sksurv |
| Cox Proportional Hazards penalized | R / glmnet |
| Cox Random Effects | R / CoxHD |
| Cox Boosting | R / CoxBoost |
| Support Vector Machine | Python / sksurv.svm.fastsurvivalSVM |
| Parallelization | R / parallel |
| Concordance Metric | R / SurvC1 |

### Supplementary Appendix

#### 1. Study Participants

##### 1.1 AML NCRI Cohort (Validation)

The following AML Study Group (AML 17 NCRI) institutions and investigators participated in this study:

Dominic Culligan, M.D, Aberdeen Royal Infirmary, UK; Kiran Tawana, M.D, Addenbrookes University Hospital, UK; Ranjit Dasgupta, M.D, Arrowe Park Hospital, UK; Andres Virchis, M.D, Barnet General Hospital, UK; Mary Mcmullin, Prof, Belfast City Hospital, UK; Manoj Raghavan, M.D, Birmingham Heartlands Hospital, UK; Paul Cahalin, M.D, Blackpool Victoria Hospital, UK; Sam Ackroyd, M.D, Bradford Royal Infirmary, UK; Priyanka Mehta, M.D, Bristol Haematology and Oncology Centre, UK; Adam Rye, M.D, Cheltenham General Hospital, UK; Emma Welch, M.D, Chesterfield Royal Hospital, UK; Salaheddin Tueger, M.D, Countess of Chester Hospital, UK; Hannah Hunter, M.D, Derriford Hospital, UK; Atchamamba Bobbili, M.D, Doncaster Royal Infirmary, UK; Roderick Neilson, M.D, Falkirk and District Royal Hospital, UK; Earnest Heartin, M.D, Glan Clwyd Hospital, UK; Adam Rye, M.D, Gloucestershire Royal Hospital, UK; Norbert Blesing, M.D, Great Western Hospital, UK; Kavita Raj, M.D, Guys & St Thomas Hospital, UK; Jirri Pavlu, M.D, Hammersmith Hospital, UK; Richard Kaczmarski, M.D, Hillingdon Hospital, UK; Debo Ademokun, M.D, Ipswich Hospital, UK; Angela Wood, M.D, James Cook University Hospital, UK; Cesar Gomez, M.D, James Paget Hospital, UK; Paresh Vyas, M.D, John Radcliffe Hospital, UK; Jindriska Lindsay, M.D, Kent and Canterbury Hospital, UK; Mark Kwan, M.D, Kettering General hospital, UK; Ghulam Mufti, Prof, Kings College Hospital, UK; Kate Hodgson, M.D, Leicester Royal Infirmary, UK; Charlotte Kallmeyer, M.D, Lincoln County Hospital, UK; Eleni Tholouli, M.D, Manchester Royal Infirmary, UK; Maadh Aldouri, M.D, Medway Maritime Hospital, UK; Moez Dungarwalla, M.D, Milton Keynes Hospital, UK; Simon Bolam, M.D, Musgrove Park Hospital, UK; Abraham Jacob, M.D, New Cross Hospital, UK; S Tauro, M.D, Ninewells Hospital and Medical Centre, UK; Matthew Lawes, M.D, Norfolk and Norwich University Hospital, UK; Nicki Panoskaltsis, M.D, Northwick Park Hospital, UK; Jenny Byrne, M.D, Nottingham University Hospital and Paediatrics, UK; Sateesh Nagumantry, M.D, Peterborough City Hospital, UK; Paul Moreton, M.D, Pinderfields Hospital, UK; Darshayani Furby, M.D, Poole Hospital, UK; Mansour Ceesay, M.D, Princess Royal Hospital, UK; Mary Ganczakowski, M.D, Queen Alexandra Hospital, Portsmouth, UK; Charles Craddock, Prof, Queen Elizabeth Hospital, Birmingham, UK; Peter Coates, M.D, Queen Elizabeth Hospital, Kings Lynn, UK; Sabia Rashid, M.D, Queen Elizabeth Hospital, Woolwich, UK; Paul Greaves, M.D, Queens Hospital, UK; Caroline Duncan, M.D, Raigmore Hospital, UK; Pratap Neelakantan, M.D, Royal Berkshire Hospital, UK; Rachel Hall, M.D, Royal Bournemouth Hospital, UK; Bryson Pottinger, M.D, Royal Cornwall Hospital, UK; Juanah Addada, M.D, Royal Derby Hospital, UK; Asim Khwaja, M.D, Royal Free Hospital, UK; Mohammad Mohsin, M.D, Royal Hallamshire Hospital, UK; Andrea Corcoz, M.D, Royal Shrewsbury, UK; Louise Hendry, M.D, Royal

Surrey Hospital, UK; Timothy Corbett, M.D, Royal Sussex County Hospital, UK; Josephine Crowe, M.D, Royal United Hospitals Bath NHS Foundation Trust, UK; Craig Taylor, M.D, Russells Hall Hospital, UK; Rowena Thomas-Dewing, M.D, Salford Royal Hospital, UK; Jonathan Cullis, M.D, Salisbury Hospital NHS Foundation, UK; Yasmin Hasan, M.D, Sandwell Hospital, UK; Unmesh Mohite, M.D, Singleton Hospital, UK; Deborah Richardson, M.D, Southampton General Hospital, UK; Gail Loudon, Southern General Hospital, New Victoria, UK; Jamie Cavenagh, St Bartholomew's Hospital, UK; Dr Richard Kelly, St James University Hospital, UK; Jamie Wilson, M.D, St Richard's Hospital, UK; Srinivas Pillai, M.D, Stafford Hospital, UK; Helen Eagleton, M.D, Stoke Mandeville Hospital, UK; Shikha Chattree, M.D, Sunderland Royal Hospital, UK; Marc Drummond, M.D, The Beatson WOS Cancer Centre, UK; Mike Dennis, M.D, The Christie NHS Foundation Trust, UK; Arpad Toth, M.D, The Royal Liverpool Hospital, UK; Mark Ethell, M.D, The Royal Marsden Hospital, UK; Gail Jones, M.D, The Royal Victoria Infirmary Freeman Hospital, UK; Deborah Turner, M.D, Torbay District General Hospital, UK; Asim Khwaja, Prof, University College London Hospitals, UK; Vikram Singh, M.D, University Hospital Aintree, UK; Julie Gillies, M.D, University Hospital Ayr, UK; Beth Harrison, M.D, University Hospital Coventry, Walsgrave, UK; William Gordon, M.D, University Hospital Crosshouse, UK; Naheed Mir, M.D, University Hospital Lewisham, UK; Srinivas Pillai, M.D, University Hospital of North Staffordshire, UK; Maria Szubert, M.D, University Hospital of North Tees and Hartlepool, UK; Jonathan Kell, M.D, University Hospital of Wales, UK; Kerri Davidson, M.D, Victoria Hospital, NHS Fife, UK; Toby Nicholson, M.D, Whiston Hospital, UK; Salim Shafeek, M.D, Worcestershire Royal Hospital, UK; Lee Bond, M.D, York Hospital, UK; Ernest Heartin, M.D, Ysbyty Gwynedd Hospital, UK; Ruth Spearing, M.D, Christchurch Hospital, New Zealand; John Carter, M.D, Wellington Hospital, New Zealand; Marianne Tong Severinsen, M.D, Aalborg University Hospital, Denmark; Ingolf Mølle, M.D, Aarhus University Hospital, Denmark; Ulrik Malthe Overgaard, M.D, Herlev Hospital, Denmark; Claus Werenberg Marcher, M.D, Odense Hospital, Denmark; Ulrik Malthe Overgaard, M.D, Rigshospitalet, Denmark

#### 1.2 AML SG Study Contributors

The following AML Study Group (AMLSG) institutions and investigators participated in this study:

Peter Brossart, M.D., Universitätsklinikum Bonn, Bonn Germany; Bernd Hertenstein M.D., Henrike Thomssen M.D., Klinikum Bremen-Mitte, Bremen, Germany; Rainer Haas M.D., Andrea Kuendgen M.D., Universitätsklinikum Düsseldorf, Düsseldorf, Germany; Peter Reimer M.D., Mohammed Wattad M.D., Kliniken Essen Süd, Ev. Krankenhaus Essen-Werden gGmbH, Essen, Germany; Carsten Schwaenen M.D., Klinikum Esslingen, Esslingen, Germany; Hans Günter Derigs M.D., Klinikum Frankfurt-Höchst GmbH, Frankfurt, Germany; Michael Lübbert M.D., Universitätsklinikum Freiburg, Freiburg, Germany; Alexander Burchardt M.D., Matthias Rummel M.D., Universitätsklinikum Gießen, Gießen, Germany; Volker Runde M.D., Wilhelm-Anton-Hospital, Goch, Germany; Gerald Wulf M.D., Lorenz Trümper M.D., Universitätsklinikum Göttingen, Göttingen, Germany; Walter Fiedler M.D., Universitätsklinikum Hamburg-Eppendorf, Hamburg, Germany; Hans Salwender M.D., Asklepios Klinik Altona, Hamburg, Germany; Elisabeth Lange M.D., Evangelisches Krankenhaus Hamm, Hamm,

Germany; Andrea Sendler M.D., Klinikum Hanau, Hanau, Germany; Arnold Ganser M.D., Jürgen Krauter, M.D., Brigitte Schlegelberger M.D., Medizinische Hochschule Hannover, Hannover, Germany; Hartmut Kirchner M.D. KRH Klinikum Siloah, Hannover, Germany; Uwe Martens M.D., SLK-Kliniken GmbH Heilbronn, Heilbronn, Germany; Michael Pfreundschuh M.D., Gerhard Held M.D., Universitätsklinikum des Saarlandes, Homburg, Germany; David Nachbaur M.D., Günter Gastl M.D., Universitätsklinikum Innsbruck, Innsbruck, Austria; Mark Ringhoffer M.D., Martin Bentz M.D., Städtisches Klinikum Karlsruhe gGmbH, 4 Karlsruhe, Germany; Heinz A. Horst M.D., Michael Kneba M.D., Universitätsklinikum Schleswig-Holstein–Campus Kiel, Kiel, Germany; Stephan Kremers M.D., Caritas-Krankenhaus Lebach, Lebach, Germany; Andreas Petzer M.D., Krankenhaus der Barmherzigen Schwestern Linz, Linz, Austria; Gerhard Heil M.D., Klinikum Lüdenscheid, Lüdenscheid, Germany; Thomas Kindler M.D., Matthias Theobald M.D., Universitätsklinikum Mainz, Mainz, Germany; Katharina Götze M.D., Christian Peschel M.D., Klinikum rechts der Isar der Technischen Universität München, München, Germany; Sabine Struve M.D. Klinikum Schwabing, München, Germany; Peter Schmidt M.D., Städtisches Klinikum Neunkirchen, Neunkirchen, Germany; Ali-Nuri Hünerlitürkoglu M.D., Lukaskrankenhaus GmbH Neuss, Neuss; Germany; Claus-Henning Köhne M.D., Klinikum Oldenburg, Oldenburg, Germany; Axel Matzdorff M.D., Caritas-Klinik St. Theresia, Saarbrücken, Germany; Richard Greil M.D., Gudrun Russ M.D., Universitätsklinikum der Paracelsus Medizinischen Universität Salzburg, Salzburg, Austria; Jochen Greiner M.D., Diakonie-Klinikum Stuttgart, Stuttgart, Germany; Heinz Kirchen M.D., Krankenhaus der Barmherzigen Brüder, Trier, Germany; Hans-Gernot Biedermann M.D., Kreisklinik Trostberg, Trostberg, Germany; Helmut Salih M.D., Lothar Kanz M.D., Universitätsklinikum Tübingen, Tübingen, Germany; Hartmut Döhner M.D., Konstanze Döhner M.D., Richard F. Schlenk M.D, Universitätsklinikum Ulm, Ulm, Germany; Wolfgang Brugger M.D., Schwarzwald-Baar Klinikum Villingen-Schwenningen GmbH, Villingen-Schwenningen, Germany; Elisabeth Koller M.D., Hanuschkrankenhaus, Wien, Austria; Aruna Raghavachar M.D., Helios-Klinikum Wuppertal, Wuppertal, Germany.
